# Federated Target Trial Emulation using Distributed Observational Data for Treatment Effect Estimation

**DOI:** 10.1101/2025.05.02.25326905

**Authors:** Haoyang Li, Chengxi Zang, Zhenxing Xu, Weishen Pan, Suraj Rajendran, Yong Chen, Fei Wang

## Abstract

Target trial emulation (TTE) aims to estimate treatment effects by simulating randomized controlled trials using real-world observational data. Applying TTE across distributed datasets shows great promise in improving generalizability and power but is always infeasible due to privacy and data-sharing constraints. Here we propose a Federated Learning-based TTE framework, FL-TTE, that enables TTE across multiple sites without sharing patient-level data. FL-TTE incorporates federated protocol design, federated inverse probability of treatment weighting, and a federated Cox proportional hazards model to estimate time-to-event outcomes across heterogeneous data. We validated FL-TTE by emulating Sepsis trials using eICU and MIMIC-IV data from 192 hospitals, and Alzheimer’s trials using INSIGHT Network across five New York City health systems. FL-TTE produced less biased estimates than traditional meta-analysis methods when compared to pooled results and is theoretically supported. Our FL-TTE enables federated treatment effect estimation across distributed and heterogeneous data in a privacy-preserved way.

## Introduction

Randomized Controlled Trials (RCTs) are the golden standard for estimating the efficacy of interventions. However, RCTs are expensive and time-consuming, and their stringent eligibility criteria exclude a large number of patients who could receive the treatment in the real world, which may lead to suboptimal estimation of the real-world effectiveness of the treatment. In the past decade, with the rapid development of computer hardware and software technologies, large amounts of patient health information have been collected and collected outside RCTs. These data also referred to as real-world data (RWD), including electronic health records (EHR), pharmaceutical and insurance claims, and others, contain insights into how medical devices and interventions work in usual care settings, and are thus instrumental for understanding healthcare effectiveness, safety, and patient effectiveness in real-world settings.^1,2^

Target trial emulation (TTE) is an approach in observational research that aims to mimic (or “emulate”) the design of an RCT using RWD.^3^ This method helps to make causal inferences about treatment effects by carefully designing the study to control biases common in observational settings. Compared with actual RCTs, TTE is more economic, and efficient, and the results derived from TTE are more representative of real-world patients. Several recent studies have demonstrated the promise of TTE in different disease contexts.^4–8^ Although treatment assignment in RWD is not randomized, TTE explicitly specifies experiment protocols to emulate randomization and mitigate potential biases with causal inference methods such as propensity score matching (PSM),^9,10^ inverse probability of treatment weighting (IPTW)^11–13^ and G-computation^14,15^. In order to achieve sufficient balance of confounding variables between treatment and control groups using these methods (e.g., measured by standard mean difference^16^), a descent sample size is required for both groups.^17,18^ Moreover, most of the TTE works were only conducted with a single institutional RWD warehouse,^19–23^ which may limit the generalization ability of the results due to the lack of diversity of the patient populations included.

With the reasons above, it is desirable to have a large RWD warehouse including diverse patient characteristics when performing TTE studies. This typically requires leveraging the patient data from multiple institutions. There have been efforts to build up large centralized repositories by aggregating the patient data from different institutions,^24–26^ but they are sporadic due to the sensitivity of patient health information, which makes them challenging to share outside the local institutions. Federated Learning (FL)^27,28^ is a promising paradigm that facilitates collaborative machine learning with data distributed across multiple local clients. FL does not require the data to be shared out of the local clients but only share model parameter updates with others, so that the data privacy is preserved.

With this appealing characteristic, FL has raised considerable attention from a broad set of applications,^29–31^ including healthcare and medicine, where FL has been applied in problems like disease diagnosis^32,33^ and clinical risk prediction.^34–36^ However, it is largely unknown how to leverage the TTE and FL frameworks to estimate real-world treatment effects using the distributed sites without sharing patient-level information.

In this paper, we propose a *Federated Learning-based Target Trial Emulation (FL-TTE)* framework to estimate real-world treatment effects using EHRs from distributed clinical institutions in a privacy-preserved way (Figure 1). The proposed FL-TTE effectively leverages siloed patient EHR data without sharing them, boosts sample size, balances confounders better, and achieves less-biased estimates compared to traditional meta-analysis methods, towards potentially more generalizable estimates for a bigger and more diverse population. Empirically, we systematically evaluated our FL-TTE framework with two different clinical research network datasets with applications for estimating repurposing treatment signals for two different diseases including Alzheimer’s disease (AD)^37^ and sepsis^38^ using longitudinal EHR data which were distributed across heterogeneous sites. Specifically, we leveraged the INSIGHT clinical research network (CRN), which includes 5,532,428 patients from the hospital systems of the greater New York City area, to estimate a range of repurposing agents for Alzheimer’s disease (AD). For the sepsis case, we used the eICU^26^ and MIMIC-IV^39^ datasets, comprising 274,040 patients from 192 sites, to investigate how corticosteroids might impact sepsis outcomes in the ICU settings. In both cases, our FL-TTE achieved less-biased treatment effect estimates than two typical meta-analysis methods^40,41^ when compared to the estimates from the pooled data (considered the gold standard but often infeasible to obtain due to privacy concerns of sharing patient data^42^), better global covariates balancing, dealing with sites’ heterogeneity well, and easily incorporated differential privacy component for better local data protection. Theoretically, we proved the less-biases of estimand from our FL-TTE by proving a better error bound than the meta-analysis methods. Our FL-TTE provides a unified framework to conduct TTE across heterogeneous datasets without exchanging patient data, and our empirical and theoretical investigations can facilitate potentially more generalizable and privacy-preserved treatment effect estimation from federated causal inference in observational studies.

**Figure 1:**
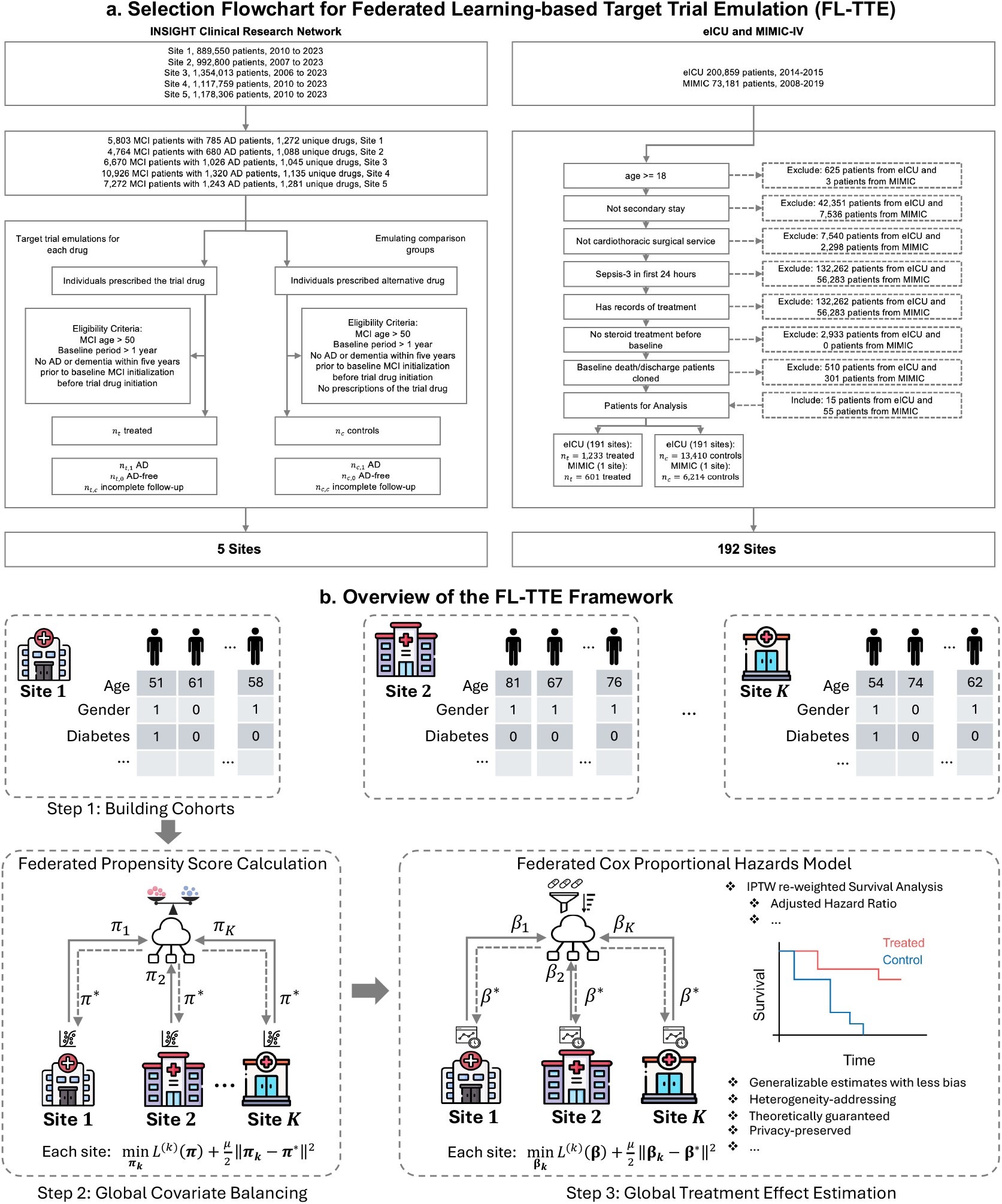
Federated Target Trial Emulation with Distributed Observational Data for Treatment Effect Estimation. (a) Selection Flowchart for Federated Learning-based Target Trial Emulation (FL-TTE). The study cohorts were from five sites within INSIGHT CRN and 192 sites in eICU and MIMIC-IV database, with applications of estimating different drug repurposing signals for Alzheimer’s disease and sepsis, respectively. (b) Overview of the FL-TTE Framework. Step 1: Cohorts were constructed from INSIGHT and eICU-MIMIC datasets, respectively. Step 2: Federated propensity score calculation adjusted for differences in patient covariates between treated and control groups with inverse probability of treatment weighting (IPTW) for achieving the global covariate balancing. Step 3: Federated Cox proportional hazards model estimated the treatment effects of target drugs for achieving less-biased global time-to-event outcome estimates. The optimizations are regularized by the proximal term which can ensure local updates align with the global model, limit the impact of over-large local updates that can induce overfit, and finally address the data heterogeneity among sites.

## Results

### Cohort Characteristics and Heterogeneity

Our study cohorts include the INSIGHT clinical research network,^43^ eICU and MIMIC-IV. For the INSIGHT cohort, there was a total of 35,435 eligible patients with at least one mild cognitive impairment (MCI) documented diagnosis between August 2006 and December 2023, which comprises of 5803, 4764, 6670, 10926, and 7,272 patients from each of five sites, respectively. The treated group includes individuals exposed to the target drug, while the control group contains the individuals treated by an alternative drug. The patient inclusion cascade and population characteristics are presented in Figure 1a and Supplementary Table 4. For the eICU-MIMIC cohort, there is a total of 200,859 patients from 191 sites from eICU time from 2014 to 2015 and 73,181 patients from the single site in MIMIC from 2008 to 2019. The cohort includes 1,233 treated patients and 13,410 controls from eICU, 601 treated patients, and 6,214 controls from MIMIC with the inclusion cascade shown in in Figure 1b.

We observed substantial heterogeneity in sample distributions across different sites (Figure 2). Specifically, Figure 2a illustrates the geographic locations of five sites from the INSIGHT in NYC. Patients in geographically different communities have different demographics as demonstrated in Figure 2c. For example, Site 4 has the highest proportion of self-reported White patients, and Site 2 has the largest proportion of self-reported Black or African American patients. Further, the disease progression characteristics across different patient cohorts are different. Figure 2b shows the Kaplan-Meier survival curve for patients progressing from MCI to AD across the 5 sites, where Site 1 exhibits the steepest decline in survival probability, while Site 4 demonstrates the slowest progression speed. Regarding the eICU-MIMIC cohort, Figure 2d illustrates the distribution of cohort sizes across 192 sites, which shows that although some sites include over 1,000 patients, there are 52 (27%) sites that have fewer than 10 patients.

**Figure 2:**
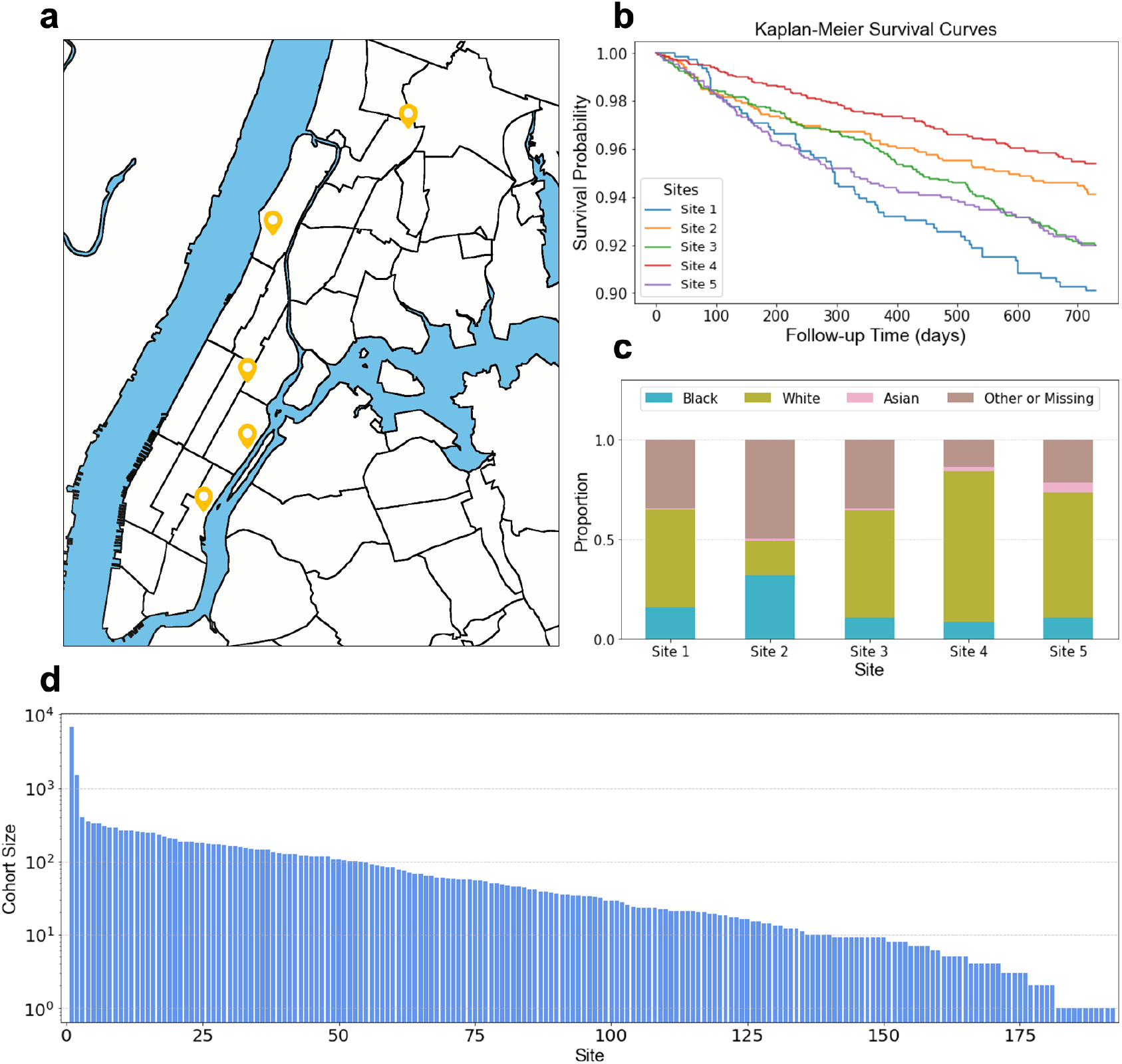
Data Heterogeneity Across INSIGHT and eICU-MIMIC Cohorts. (a) Geographic locations of the five INSIGHT sites in New York City. (b) Kaplan-Meier survival curves44 illustrated the data heterogeneity in survival probabilities across five INSIGHT sites. (c) Different race distributions varied among five INSIGHT sites. Site 4 has the highest proportion of self-reported White patients, and Site 2 has the largest proportion of self-reported Black or African American patients. (d) Logarithmic cohort size distribution across 192 sites in the eICU and MIMIC dataset exhibited a long-tailed pattern.

### FL-TTE Achieves Less Biased Estimates Than Local Analysis Methods

We evaluated the effectiveness of FL-TTE in the INSIGHT and eICU-MIMIC cohorts by emulating different target trials and comparing the results with the estimates from local data of each site and the global pooled data. We assume that the heterogeneity among multiple sites exists in baseline covariates but not the treatment effect,^45–47^ so that the pooled analysis can be a gold standard serving as an ideal benchmark for assessing the bias of the estimators.^48–50^

For the INSIGHT cohort, we emulated nine target trials investigating the effects of drugs with potential benefits for patients who are at risk for AD^8^ (see Methods for details). FL-TTE consistently produced less-biased estimates than the ones generated by local data analysis (see Figure 3). Specifically, With the results from pooled data analysis as references, FL-TTE typically had smaller Z-test statistics^51^, indicating greater similarity, and higher p-values, suggesting no significant differences, when compared with the results from local data analysis. The local analysis gave highly heterogeneous estimates across the five sites that did not align well with the pooled estimates, showing the large *I*^2^ statistics across the five sites in all trials on target drugs (0.942±0.008) using Cochran’s Q test^52^ (which can assess heterogeneity and “high heterogeneity” associates with *I*^2^ ≥ 0.5). For local analysis, in seven out of the nine target trials, we observed estimates with conflict directions across the five sites. For example, in the case of pantoprazole, at Sites 1, 3, and 5, the estimates suggested a decreased risk for AD onset, with aHRs of 0.85 (95% CI: 0.83–0.88), 0.79 (95% CI: 0.75–0.83), and 0.92 (95% CI: 0.89–0.95), respectively, while at Site 2 and Site 4, the estimates indicated an increased risk for AD onset, with aHRs of 1.09 (95% CI: 1.01–1.16) and 1.17 (95% CI: 1.15–1.18), respectively.

**Figure 3:**
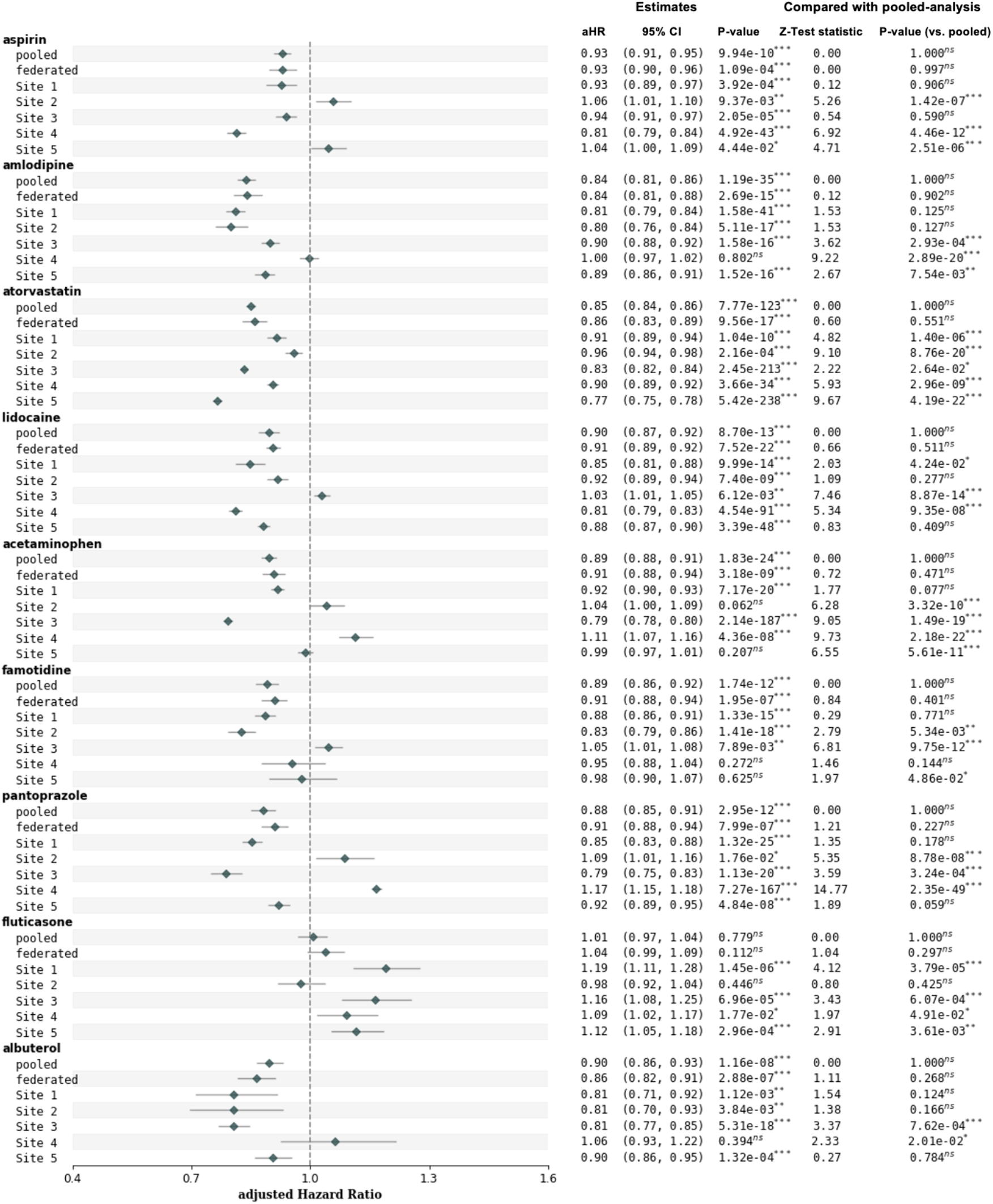
The estimated aHR and 95% CI on INSIGHT, comparing pooled analysis, our FL-TTE and local analysis. The third column in the right side is p-value and significance level of the Z-test on whether the estimated aHR is significantly different with 1.0 (reference value indicating the treatment does not alter the risk compared to no treatment). The fourth and fifth columns denote the test statistic and p-value of the Z-test on whether the estimated aHR is significantly different with the results of pooled analysis. Our FL-TTE addressed the poorly generalized single-site’s estimates induced by sites’ heterogeneity and achieved similar estimates with pooled-analysis. *p < 0.05; **p < 0.01; ***p < 0.001; not significant (“ns”) with p ≥ 0.05.

For the eICU-MIMIC cohort, we emulated a target trial aimed at evaluating the effects of corticosteroid treatment on sepsis. The aHR estimates with 95% confidence intervals (CIs) produced by FL-TTE were closer to the pooled results compared to the results from local analysis. Figure 4 shows the results from the five sites with the largest cohort sizes, which demonstrated larger bias (compared to the results from pooled analysis) quantified by Z-test^51^. For example, local analysis overestimated aHR on Site 3 with 1.32 (95% CI: 1.24-1.41, p < 0.001), 1.13 (95% CI: 1.06-1.20, p < 0.05), 1.24 (95% CI: 1.17-1.31, p < 0.001) in the three outcomes (28-day mortality, ICU discharge, and cessation of mechanical ventilation), showing significantly different estimates with the pooled results 1.10 (95% CI: 1.05-1.15), 1.03 (95% CI: 0.99-1.08), 1.03 (95% CI: 0.98-1.08) in these three outcomes. The estimates among these five sites also had high heterogeneity (*I*^2^ statistics=0.892±0.007 in all the three trials using Cochran’s Q test^52^), indicating the potential inconsistency of local analysis compared to pooled results.

**Figure 4:**
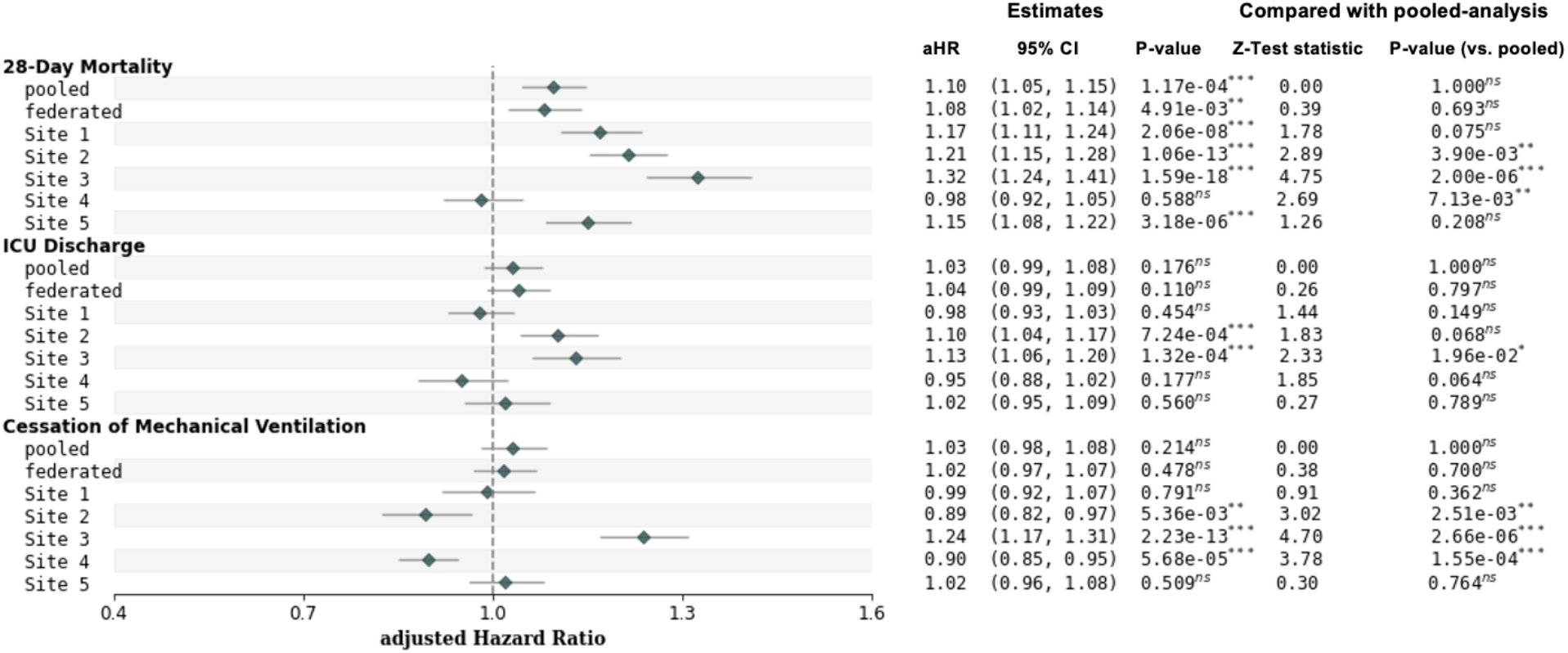
The estimated aHR and 95% CI on eICU-MIMIC, comparing pooled analysis, our FL-TTE and local analysis on the top 5 of 192 sites with the largest cohort sizes. The third column in the right side is p-value and significance level of the Z-test on whether the estimated aHR is significantly different with 1.0 (reference value indicating the treatment does not alter the risk compared to no treatment). The fourth and fifth columns denote the test statistic and p-value of the Z-test on whether the estimated aHR is significantly different with the results of pooled analysis. *p < 0.05; **p < 0.01; ***p < 0.001; not significant (“ns”) with p ≥ 0.05.

### FL-TTE Achieves Less Biased Estimates Than Meta-Analysis Methods

We tested the effectiveness of FL-TTE on both INSIGHT and eICU-MIMIC cohorts with different target trials through the comparison with the results from two representative meta-analysis methods^40^, including the fixed-effect model and random-effect model, as well as the the estimates derived from the pooled data.

For INSIGHT, we emulated nine target trials focusing on drugs that could be potentially repurposed to AD^8^ (see details in Methods). As shown in Figure 5, FL-TTE achieved aHR estimates with 95% confidence intervals (CIs) overlapping more with the pooled estimates compared to the meta-analysis methods, while at the same time with narrower confidence intervals. For the two meta analysis approaches, the fixed effect model tends to be more biased (i.e., with different estimation compared to the pooled results) with less variance (narrow CI), while meta analysis with random effect model tends to be less biased but much larger CI. We further quantified the difference using Z-test^51^. As shown in Figure 5, the Z-test statistics between FL-TTE and pooled results are typically smaller (indicating more similarity) with larger p-values compared to meta-analysis results. Interestingly, for pantoprazole, the two meta-analysis approaches gave estimates on different directions, where the fixed effects estimated an aHR of 1.09 (95% CI: 1.05–1.13, p < 0.001), while the random effects estimated an aHR of 0.95 (95% CI: 0.82–1.10, p = 0.304). This implies the potential instability of different meta analysis methods when facing with site heterogeneity.

**Figure 5:**
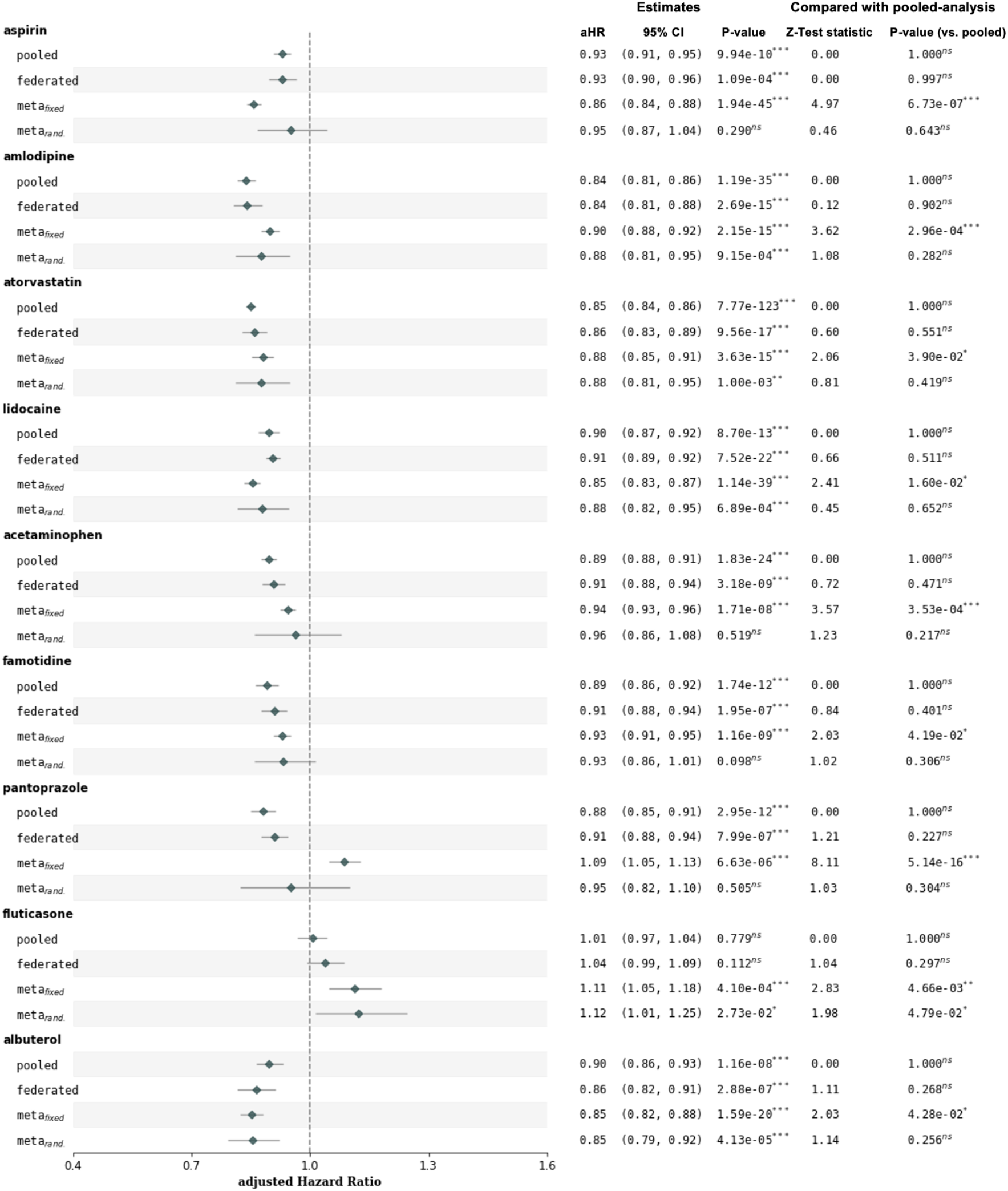
The estimated aHR and 95% CI on INSIGHT, compared with pooled analysis, our FL-TTE and meta-analysis with fixed effects and random effects. The third column on the right side is the p-value and significance level of the Z-test on whether the estimated aHR is significantly different with 1.0 (reference value indicating the treatment does not alter the risk compared to no treatment). The fourth and fifth columns denote the test statistic and p-value of the Z-test on whether the estimated aHR is significantly different from the results of pooled analysis. Our FL-TTE achieved less-biased treatment effect estimates than two typical meta-analysis methods when compared to the estimates from the pooled data. *p < 0.05; **p < 0.01; ***p < 0.001; not significant (“ns”) with p ≥ 0.05.

For eICU-MIMIC, we emulated the target trial of corticosteroid treatment on sepsis (see details in Methods). As shown in Figure 6, our FL-TTE consistently outperformed meta-analysis methods in estimating less-biased aHRs across three outcomes (28-day mortality, ICU discharge, and cessation of mechanical ventilation) compared to the pooled results. In particular, under 28-day mortality outcome, FL-TTE achieved an aHR of 1.08 (95% CI: 1.02–1.14), closely approximating the pooled aHR of 1.10 (95% CI: 1.05–1.15) with a non-significant z-test (0.39, p = 0.693). Meta-analysis with fixed effects overestimated the aHR of 1.16 (95% CI: 1.09–1.23), and random effects underestimated the aHR of 1.01 (95% CI: 0.94–1.07, p = 0.033), compared to the results obtained from pooled analysis.

**Figure 6:**
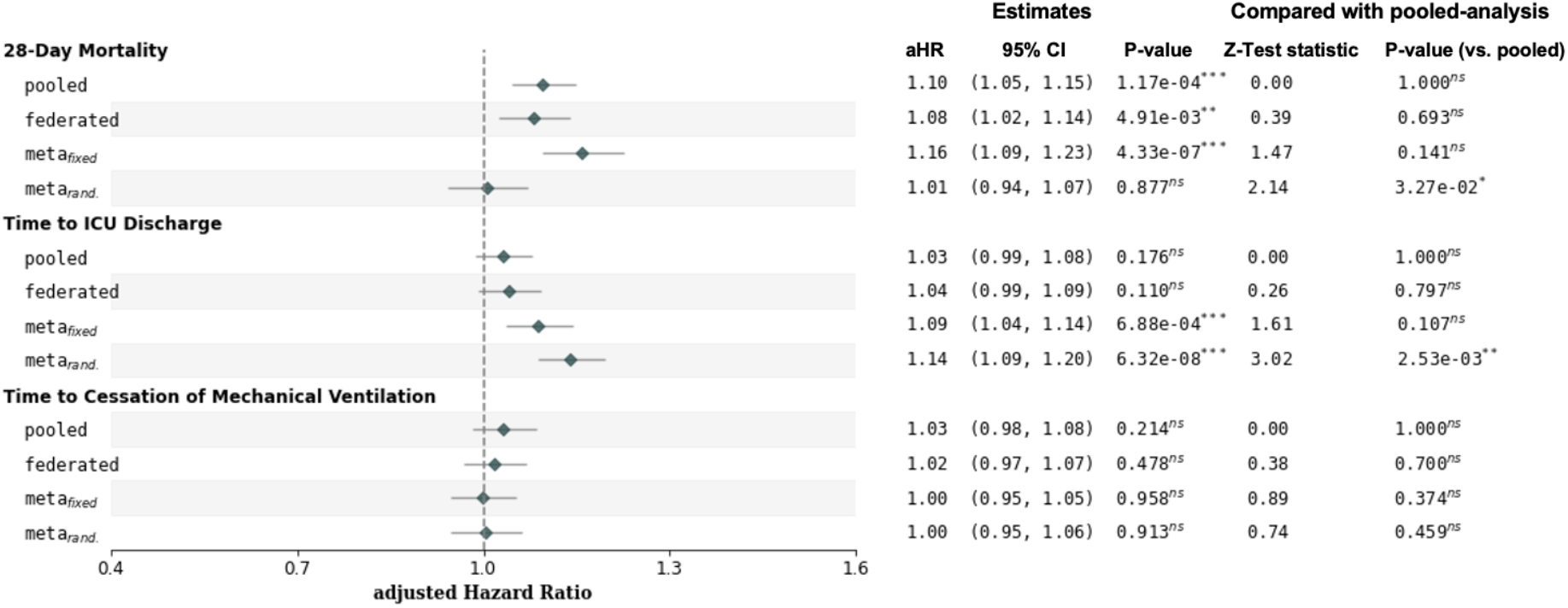
The estimated aHR and 95% CI on eICU and MIMIC, comparing pooled analysis, our FL-TTE and meta-analysis with fixed effects and random effects. The third column on the right side is the p-value and significance level of the Z-test on whether the estimated aHR is significantly different with 1.0 (reference value indicating the treatment does not alter the risk compared to no treatment). The fourth and fifth columns denote the test statistic and p-value of the Z-test on whether the estimated aHR is significantly different from the results of pooled analysis. Our FL-TTE had less-biased estimates than the meta-analysis in three types of outcomes (28-day mortality, Time to ICU discharge, and Time to cessation of mechanical ventilation). *p < 0.05; **p < 0.01; ***p < 0.001; not significant (“ns”) with p ≥ 0.05.

### FL-TTE Achieves Better Global Covariate Balance

FL-TTE also achieved higher success balancing ratios in adjusting for confounders on INSIGHT and eICU-MIMIC datasets than both the local analysis (see Figure 7 and Figure 8) and meta-analysis methods (see Figure 9 and Figure 10). For INSIGHT CRN (see Figure 9), the pooled-analysis achieved near-optimal covariate balancing ratios across all trials on target drugs (0.965±0.067), and FL-TTE closely approximated this performance (0.926±0.066). In contrast, meta-analysis methods demonstrated lower balancing ratios, particularly with fixed effects, where the ratios for drugs dropped to 0.767±0.055. The random-effects meta-analysis showed slightly better performance (0.772±0.062) but remained inferior to the federated method. As shown in Figure 7, the local analysis also did not achieve sufficient balance of confounding variables (e.g., 0.721±0.062 in Site 1, and 0.683±0.208 in site 5) under the smaller sample size of each site than pooled data. For eICU-MIMIC, FL-TTE achieved balancing ratios 0.985±0.014 across all outcomes. The pooled analysis consistently reached the optimal ratio of 1.000±0.000. However, neither meta-analysis methods nor local analysis cannot balance the covariates well. For example, the fixed-effects meta-analysis model achieved a balancing ratio of 0.667±0.000 under three outcomes, while the random-effect meta-analysis reported ratios 0.722±0.000.

**Figure 7:**
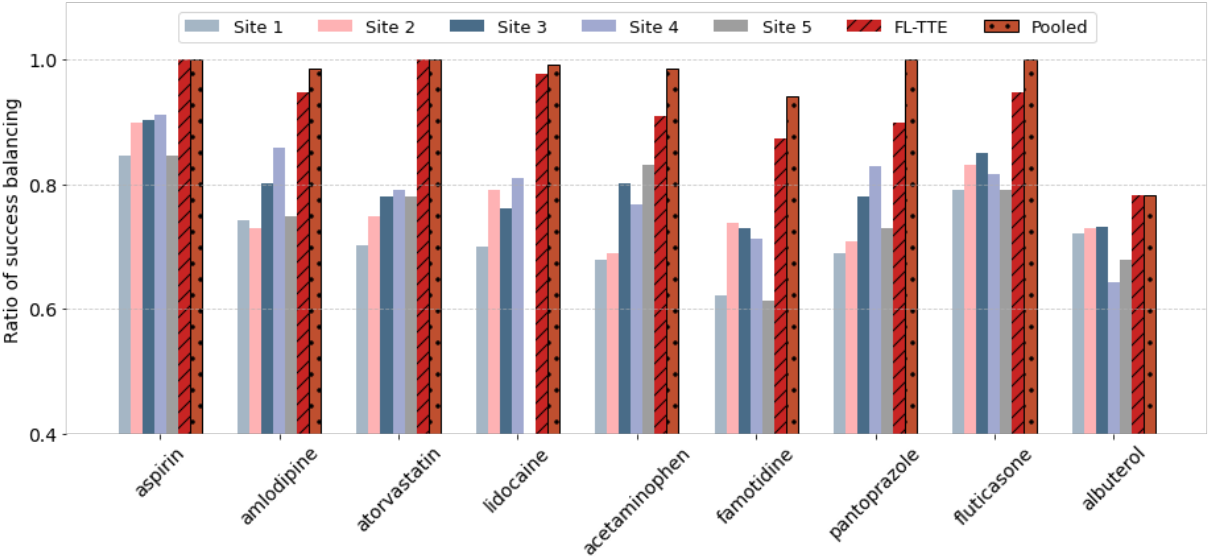
Ratio of success balancing in INSIGHT before and after reweighting with our FL-TTE, single-site analysis, and pool-analysis method. The sample size of each site is shown above each bar. For eICU-MIMIC in (b), we present the single-site results by selecting the top 5 of 192 sites with the largest cohort sizes.

**Figure 8:**
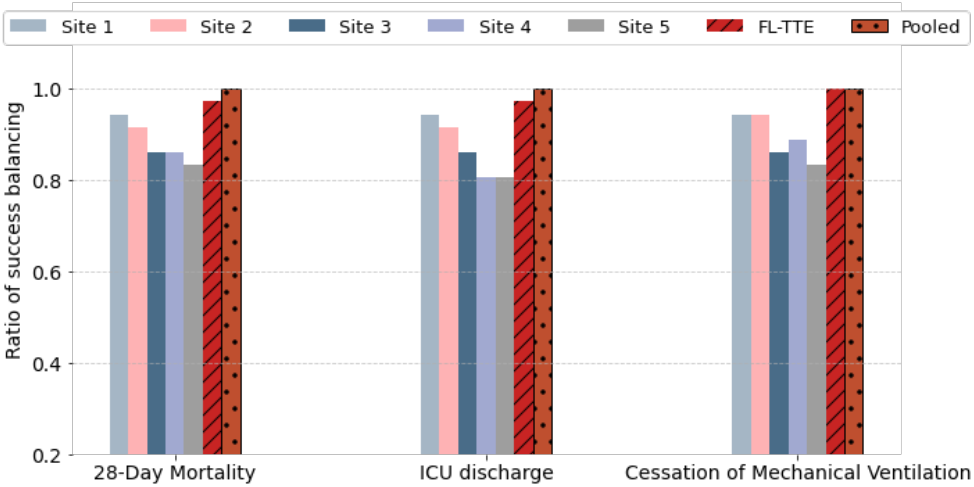
Ratio of success balancing in eICU-MIMIC before and after reweighting with our FL-TTE, single-site analysis, and pool-analysis method. The sample size of each site is shown above each bar. For eICU-MIMIC in (b), we present the single-site results by selecting the top 5 of 192 sites with the largest cohort sizes.

**Figure 9:**
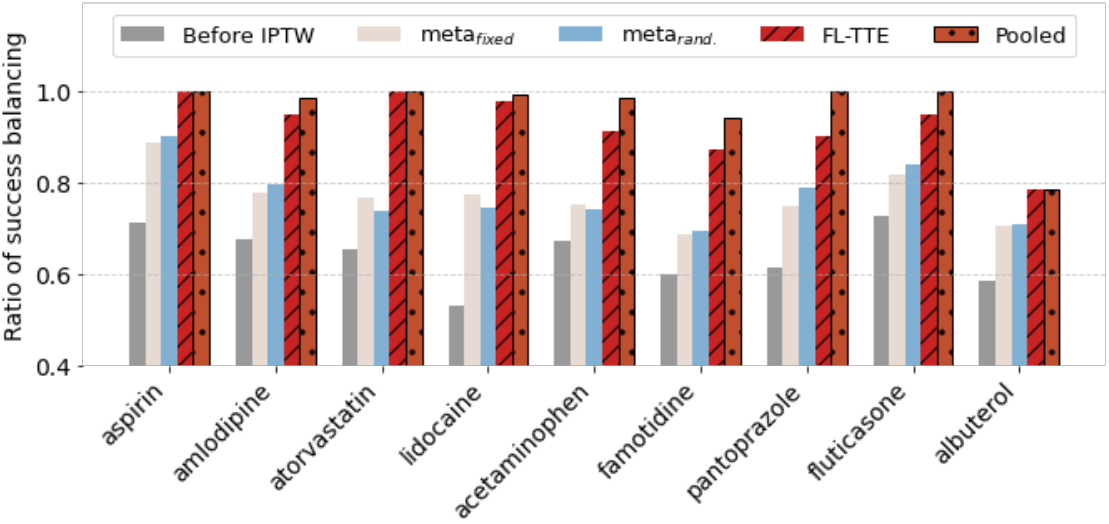
Ratio of success balancing in INSIGHT before and after reweighting with our FL-TTE, meta-analysis methods, and pool-analysis method. The FL-TTE achieved higher success balancing ratios in adjusting for covariates on both INSIGHT and eICU-MIMIC datasets than meta-analysis with fixed and random effects.

**Figure 10:**
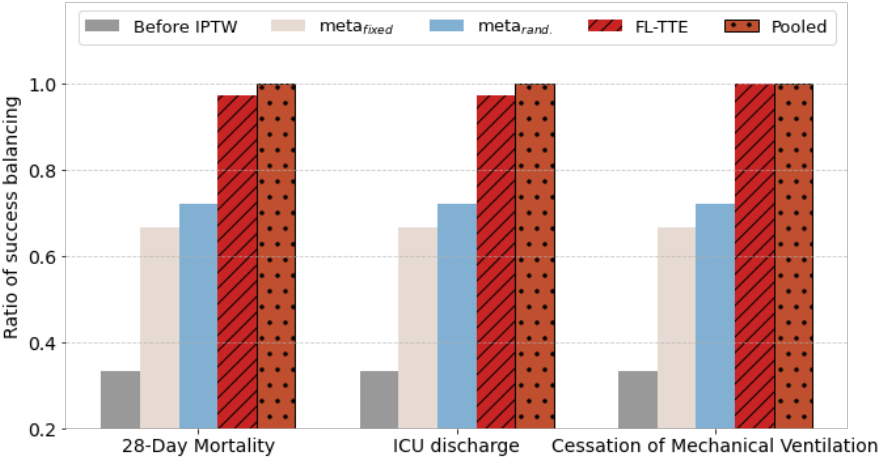
Ratio of success balancing in eICU-MIMIC before and after reweighting with our FL-TTE, meta-analysis methods, and pool-analysis method. The FL-TTE achieved higher success balancing ratios in adjusting for covariates on both INSIGHT and eICU-MIMIC datasets than meta-analysis with fixed and random effects.

### Theoretical Guarantee

In addition to empirical evaluation, we also proved theoretically that FL-TTE can achieve less biased estimations than meta-analysis methods. Theorem 1 in Box 2 establishes that under the assumptions of *C* -Lipschitz continuity, smoothness, and *λ* -strong convexity of the outcome model, the bias between the FL-TTE and pooled analysis ‖ log aHR _*FL*_ − log aHR _*pool*_ ‖ is upper bounded by 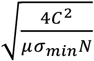. In contrast, the bias between meta-analysis and pooled analysis ‖log aHR_*meta*,_ − log aHR _*pool*_‖ is upper bounded by. 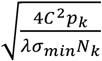 With proximal term coefficient *µ*, it is guaranteed that. 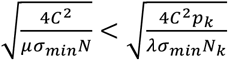, ensuring that the FL-TTE achieves a tighter bias bound compared to meta-analysis (see proof in Supplementary Note 1 and 2). Theorem 2 demonstrates the efficient convergence of our FL-TTE method, achieving a convergence rate of 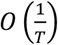, where *T* is the total number of iterations, indicating rapid approximation to the global optimum, meaning that the bias decreases significantly during the initial training rounds, bringing it close to the optimum, and continues to diminish steadily as the iterations progress. These theoretical results demonstrate the optimality and efficiency of the FL-TTE framework in achieving less-biased treatment effect estimations.

### Enhanced Privacy with Differential Privacy Techniques

While FL offers intrinsic privacy protections by retaining data within each site, model inversion and data reconstruction risks remain potential concerns.^53^ To further enhance privacy, we applied differential privacy techniques to mitigate the possibility of intercepting sensitive information from shared gradients during training. The techniques strengthen our FL-TTE framework by safeguarding against data leakage risks. Our framework still produced the less-biased aHR estimates than meta-analysis methods in 8 out of 9 trials on INSIGHT and all 9 trials on eICU-MIMIC (see Supplementary Figures 1 and 2).

### Sensitivity Analyses

To test the robustness of our framework, we conducted the following sensitivity analyses. First, we tested different regularizers when estimating the propensity scores and outcomes, including FedAvg^54^ which directly aggregates locally trained models on multiple sites, FedAvgM^55^ which introduces the momentum to address heterogeneity, and FedProx^56^ which encourages the consistency between local and global models (see Methods). We also compared it with Federated IPW-MLE introduced by Xiong et al.^57^ As shown in Supplementary Figures 3 and 4, the estimation results are not sensitive to the different choices of federated learning algorithms. No matter which FL algorithm is used, less-biased estimates can always be achieved than meta-analysis methods compared to pooled results. And our framework can also achieve less biased estimates than method of Xiong et al.^57^ Second, as shown in Supplementary Figure 5, we reported the results by adopting the clone-censor-weight approach.^58^ Specifically, all eligible patients were cloned into both treatment strategies at a unified time zero set to ICU admission. Patients were then censored at the time they deviated from their assigned strategy (e.g., initiated or failed to initiate treatment). To further account for potential bias introduced by non-random censoring, we applied inverse probability of censoring weights (IPCW) based on baseline covariates. This method ensures that both treatment and control groups have the same starting time point, thereby eliminating additional immortal time. We implemented this procedure and repeated the federated target trial emulation under the new time zero definition. The results are largely same as the primary results in Figure 6.

## Discussions

Although randomized controlled trials (RCTs) are still the golden standard of evaluating the effectiveness and satefy of interventions, they are expensive and time-consuming to conduct, and the recruited participants are usually not representative of real world patients due to the stringent eligibility criteria. Target trial emulation (TTE) is the process of simulating clinical trials using observational data. Compared with RCTs, TTE is economic, efficient and representative of real world patients. However, due to the non-randomized nature of observational data, effective control of the impact of potential confounding factors is critical, and a reasonable sample size for both treated and comparative groups plays a key role here to ensure unbiased estimation of treatment effects, which is usually a challenge in the real world due to the sensitivity of patient health information.

In this study, we developed a federated learning framework for target trial emulation (FL-TTE) to enable treatment effect estimation by leveraging the EHR from different institutions without sharing them. Our framework includes two main steps: federated propensity score calculation for covariate balancing and federated Cox proportional hazards model for outcome prediction. We proved theoretically the optimality of FL-TTE, which means it can approximate closely to the estimate obtained from the analysis of the data pooled together, as well as its efficiency, which means it can converge with a small number of iteration steps. Our results supported and extended recent findings^59,60^ that meta-analysis methods may suffer from bias under data heterogeneity. Building upon these insights, we theoretically and empirically demonstrate that our FL-TTE better recovers pooled ground-truth estimates across distributed EHR datasets, with lower bias than meta-analysis methods in time-to-event modeling. While prior federated causal methods focused on binary or continuous outcomes, our approach integrates trial emulation and survival analysis, offering practical value for real-world treatment effect estimation under privacy constraints.

We evaluated the effectiveness of FL-TTE on two different diseases. One is Alzheimer’s disease (AD), which is the most prelevant neurodegenerative disease and takes tens of years to progress. The INSIGHT database we used in this case is the EHRs from a general civilian population in New York city area spanning 17 years. The other is sepsis, which is a prevalent deadly condition in critical care. The eICU-MIMIC database we used for this case includes the EHRs from the ICUs in 192 hospitals across the US. Comparing the two case studies, the INSIGHT data include information of general patient visits, which are typically sparse and irregular, and it is more appropriate for studying chronic diseases such as AD. eICU-MIMIC mainly contains information of patients within ICU stays, which are much denser with higher frequency, and they are necessary for study acute conditions such as sepsis. Emulating target trials for these two distinct disease conditions using the EHRs with very different characteristics can effectively demonstrate the generalizability of FL-TTE.

On both case studies, we were able to demonstrate 1) FL-TTE can obtain estimates that are much closer to the pooled estimates compared with local estimates; 2) FL-TTE can better balance the covariates with the boosted sample size, while it is challenging for local sites to achieve good balancing performance, which makes their estimates not stable; 3) FL-TTE also outperformed meta analysis with regards to the quality of the estimates (closer to the pooled results with narrower confidence interval) and covariate balancing. These results validated the effectiveness of FL-TTE and its potential of enabling privacy-preserving multi-institutional collaborations on generating robust real world evidence for treatments.

The estimates derived from FL-TTE aligned well with the numbers reported from existing research. For instance, atorvastatin, a prescribed statin for managing high cholesterol and triglyceride levels, has been shown to be a potential repurposable candidate for treating AD. The study by Zang et al.^8^ reported an aHR of 0.74 (95% CI: 0.73–0.76) from the OneFlorida network^61^ and 0.92 (95% CI: 0.90– 0.94) from the MarketScan database.^62^ And the study by Zissimopoulos et al.^63^ reported an aHR 0.84 (95% CI: 0.78-0.89) among white women from Medicare beneficiaries.^64^ Similarly, using INSIGHT data,^43^ FL-TTE achieved estimates of aHR 0.86 (95% CI: 0.83–0.89). In addition, pantoprazole, a proton pump inhibitor (PPI) commonly used to treat gastroesophageal reflux disease, esophageal damage, and excessive stomach acid production caused by tumors and was also identified as a repurposing candidate for AD,^65,66^ was reported with an association with reduced risk of AD onset with aHR 0.81 (95% CI: 0.80–0.83) from the OneFlorida^61^ and aHR 0.94 (95% CI: 0.92–0.96) from MarketScan^62^, and FL-TTE estimated an aHR 0.91 (95% CI: 0.88–0.94). For the case of sepsis, corticosteroids was shown to be associated with an increased risk of 28-day mortality due to exacerbated immunosuppression and a higher incidence of acute kidney injury,^67,68^ with an aHR of 1.10 (95% CI: 1.04–1.16) as reported by Rajendran et al.^69^ In our analysis, FL-TTE also estimated an aHR of 1.08 (95% CI: 1.02–1.14) for 28-day mortality.

We further enhanced the privacy protection of FL-TTE with the differential privacy technique,^70,71^ where we perturbed the shared gradients when updating the model parameters by adding Gaussian noise. With our case study evaluations, FL-TTE demonstrated enhanced privacy preservation with retained model accuracy. Our investigation further improved the practicality of FL-TTE in terms of privacy-preservation.

Our study is not without limitations. First, our analyses estimated intention-to-treat (ITT) effects considering its simplicity, inclusiveness, and better reflecting real-world effectiveness than per-protocol effect. To develop federated learning framework for per-protocol effect estimation is a promising future direction. Second, we used pooled analysis as a gold standard for estimating treatment effect^48–50^. Although it is valid under heterogeneity in baseline covariates, its validity may be limited^72–74^ when treatment effects differ substantially across sites. Future work could explore alternative benchmarks beyond pooled analysis as the gold standard under treatment effect heterogeneity. Third, while the Cox model provides a useful summary of relative risk, hazard ratio estimates may be sensitive to violations of the proportional hazards assumption. Future work could consider alternative modeling strategies such as time-varying coefficients or flexible survival models to better capture time-dependent treatment effects. Fourth, in our study, we leveraged Electronic Health Records data in our primary analysis, and we did not consider insurance coverage. We acknowledged that this could serve as one of the contributing factors for site heterogeneity. We would also like to explore this in our future study, which will further justify the necessity of our proposed federated framework by handling heterogeneity across sites.

## Method

This study was approved by the Institutional Review Board of Weill Cornell Medicine with protocol number 21-07023759. It was conducted in accordance with the Declaration of Helsinki. All EHR used in this study were fully deidentified, ethics approval and informed consent were not required.

### Federated Learning-based Target Trial Emulation (FL-TTE) Framework

#### FL-TTE framework design

In this study, we performed an intention-to-treat (ITT) analysis to assess treatment effects for two different diseases including Alzheimer’s disease (AD)^37^ and sepsis^38^. For AD-repurposed drug trials with INSIGHT data, we evaluated the effect of initiating trial drugs for patients who were confirmed with mild cognitive impairment (MCI) on delaying AD onset over a five-year follow-up period. Two treatment strategies were compared: Strategy 0, alternative drug (a similar drug within the same therapeutic class) initiation at baseline, and Strategy 1, trial drug initiation at baseline (see Supplementary Table 1 for details). It follows an active comparator new user design,^75^ in which patients newly initiating the trial drug are compared with those newly initiating an alternative drug under the same drug class captured by the Anatomical Therapeutic Chemical (ATC) level 2.^76^ For sepsis with eICU-MIMIC data, we assessed the effects of corticosteroid treatment on outcomes such as 28-day mortality, ICU discharge timing, and the duration of mechanical ventilation among those patients who were admitted to intensive care units (ICU). Two treatment strategies were compared: Strategy 0, no corticosteroid initiation within 10 hours before to 24 hours after ICU admission, and Strategy 1, hydrocortisone initiation at a dose of at least 160 mg per day during the same window. We present the summary of the FL-TTE protocol and a comparison of the target trials on INSIGHT (Supplementary Table 1) and eICU-MIMIC (Supplementary Table 2).

To achieve balance across treatment (exposed) and control (non-exposed) groups, we introduced a federated propensity score calculation model designed to adjust baseline covariates. This global logistic regression (LR) model was trained with a federated learning paradigm. Specifically, treatment assignment served as the dependent variable, while baseline covariates acted as independent variables. The propensity scores from the global LR model were used to apply the inverse probability of treatment weighting (IPTW) for each individual. For survival analysis, we proposed a federated Cox proportional hazards model (CoxPH^77,78^) to calculate global adjusted hazard ratios (aHR) across sites, with 95% confidence intervals (CIs).

### Federated Propensity Score Calculation Model

This model is to adjust for differences in patient covariates between treated and control groups, which is achieved through the propensity score (PS) representing the probability that a patient receives a treatment given the baseline covariates. Specifically, for each patient *n*, the propensity score *e*(**z**_*n*_) is defined as:

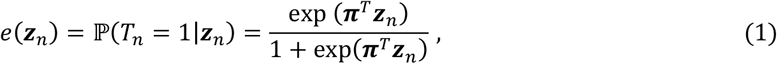

where *T*_*n*_ is a binary indicator of whether the patient received the treatment (*T*_*n*_ = 1) or not (*T*_*n*_ = 0). **z**_*n*_ represents the vector of baseline covariates for patient *n* (e.g., age, gender, medical history, etc.). **π** is the vector of PS calculation model parameters that are estimated through logistic regression (LR). The equation (1) models the likelihood of treatment assignment based on patient covariates. Next, Inverse Probability of Treatment Weighting (IPTW) is applied to balance the covariates between the treated and control groups. The weights *w*_*n*_ for each patient *n* are computed as follows:

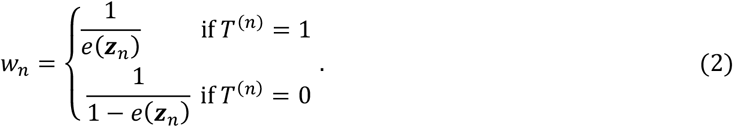

These weights help reweight the data so that the treated and control groups are balanced, which is crucial for the next-step treatment effect estimation.

In our FL-TTE framework, each site *k* computes the partial log-likelihood for the LR model:

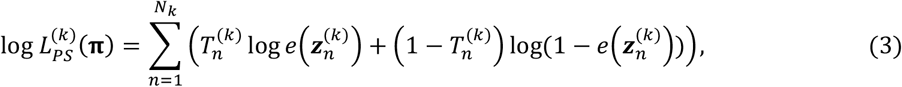

where *N*_*k*_ is the number of patients at site 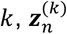 represents the covariates for patient *n* at site *k*.

After each site optimized its local model, the central server aggregates the updates to update the global PS calculation model in an interative process. Finally, the federated partial log-likelihood for all sites is:

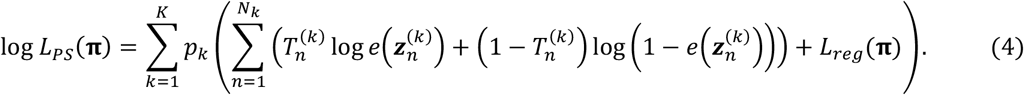

Here *K* is the total number of sites. 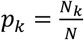 represents the proportion of the total data *n*_*k*_ located at site *k*, where *L*_*reg*_(**π**) represents the regularization term for helping FL address data heterogeneity problem. Our framework is compatible with several types of regularizers or different federated algorithms for aggregating local models. For example, (1) the regularizer can be instantiated as *L*_*reg*_(**π**) = 0, i.e. using FedAvg^54^ algorithm and no explicit regularizer for data heterogeneity issue. (2) It can alse be instantiated as 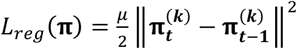, i.e. using FedAvgM^55^ algorithm that maintains smooth local model updates between two consecutive iterations *t* and *t* − 1. (3) Besides, it can be instantiated as 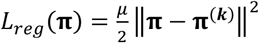, i.e. using FedProx^56^ algorithm that ensures the consistency between local model **π**^(***k***)^ and global model **π**. Here *µ*/2 is the coefficient of the regularizer. It means that each local objective equation (3) includes a proximal regularization term. The updated local parameters are then aggregated by the central server to form the global PS calculation model parameters for the next round of each local site (Box 1).

### Federate Cox Proportional Hazards Model

Once the covariates are successfully balanced, we estimate the treatment effects using a CoxPH model. The hazard function for patient *n*, given their covariates **z**_*n*_, is:

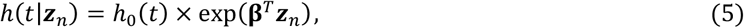

where *h*(*t*|**z**_*n*_) is the hazard rate at time *t* for a patient with covariates **z**_*n*_. *h*_0_(*t*) is the baseline hazard function (the hazard when all covariates are zero). **β** is the vector of model parameters that describes the effect of the covariates on the hazard.

Generally, at each site *k*, the partial likelihood for the Cox model is computed locally as:

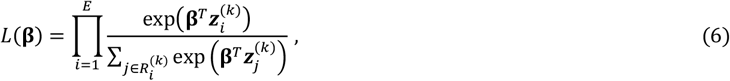

where *E* is the number of distinct event times (e.g., the times at which patients develop the outcome), 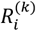 is the risk set at time *t*_*i*_, i.e., the set of patients still at risk for the event at time 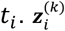 represents the covariates for patient *i* at site *k*.

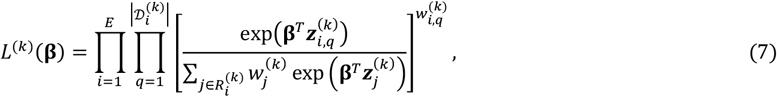

In our FL-TTE framework, we employ IPTW-adjusted Cox regression, where the partial likelihood is adjusted with the IPTW weights for each site *k*:

Where 𝒟_B_ is the set of patients with tied events at time 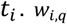 is the IPTW weight for patient *i*_C_, calculated based on the PS.

Furthermore, the federated partial likelihood aggregates these updates across all sites:

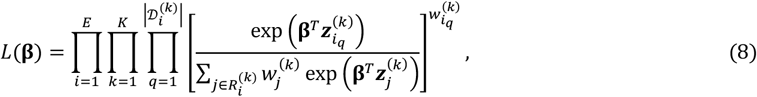

Finally, the partial log-likelihood of our federated CoxPH model is shown in equation (9):

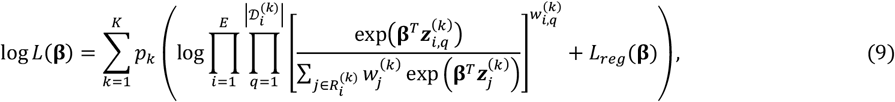

where 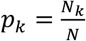 represents the proportion of the total data *N*_*k*_ located at site *k*. We add the regularization term for **β** (which can also adopt different instantiations) to address data heterogeneity during the optimization process of federated CoxPH model. Similar with equation (4), each local objective equation (8) includes a proximal regularization term. And the updated local parameters of CoxPH model optimized with equation (9) are then aggregated by the central server to form the global CoxPH model parameters for the next round of each local site.

The overall training pipeline of our FL method is summarized in Box 1.

#### Box 1

**The algorithm of our FL-TTE.**

**Input:** Given *K* sites where each site holds a local dataset *S*_*k*_, the whole dataset is 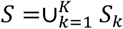. The number of epochs for federated learning is *T*.

**Parameters: *θ*** = (***π*, β**) denotes the parameter, where ***π*** is the parameter of the propensity score calculation model and **β** is the parameter of the CoxPH model.

**Output:** The estimated treatment effect adjusted hazard ratio (aHR).

1. for *t* = 1, …, *T* do
2. Server sends ***θ***^(.)^ = (***π***^(.)^, **β**^(.)^) to all local sites.
3. for each site *S*_*k*_ do
4. Calculate the objective of the federated propensity score model with equation (4).
5. Obtain the local updated parameter 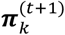.
6. Calculate the objective of the federated CoxPH model with equation (9).
7. Obtain the local updated parameter 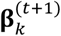.
8. Send back the updated 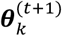 to server.
9. end for
10. Server aggregates ***π*** as 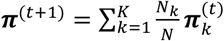.
11. Server aggregates **β** as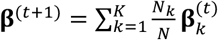.
12. Obtain the global updated parameter ***θ***^(t+1)^.
13. end for
14. Obtain the optimized parameters ***θ***^(*T*)^ = (***π***^(*T*)^, **β**^(*T*)^).
15. Estimate treatment effect using the optimized CoxPH model with **β**^(*T*)^.

### Adding Differential Privacy to FL-TTE

FL allows the participating sites to collaborate on model optimization without directly sharing sensitive patient data. However, despite this advantage, there are still inherent risks associated with the potential ‘inversion’ of the model^53^, which means it could potentially reconstruct original training data from the model’s gradients.^79^ To address the concerns, we further incorporated differential privacy techniques aimed at reducing the possibility of data reconstructions during communication between the central server and participating sites. Specifically, we explored (*ϵ, δ*)-differential privacy techniques^70,71^ that prevent the interception of sensitive data transmitted during the training process, strengthening the overall FL-TTE framework, where the privacy budget *ϵ* = 1.0 and the failure probability *δ* = 1/*N*, where *N* is the number of patients in a trial.

### Theoretical Guarantee

We present a theoretical analysis (Box 2) showing that our FL-TTE framework yields a tighter bias bound than meta-analysis, compared with the pooled results (**Theorem 1**). It highlights the strong generalization capabilities of our FL-TTE framework. Additionally, our method demonstrates a good convergence rate, significantly reducing communication costs during training, which enhances its practicality for real-world applications (**Theorem 2**). This efficiency makes it particularly well-suited for deployment in distributed healthcare systems, where bandwidth and latency constraints are often limiting factors.

#### Box 2

**Theoretical analysis of our FL-TTE framework.**

##### Theorem 1

Assuming the *C*-Lipschitz continuity^80^ and smoothness of the outcome model and its loss function is *λ*-strong convex^81^ with the parameters, the bias between our FL model and pool analysis ‖log aHR _*FL*_ − log aHR _*pool*_‖ is upper bounded with. 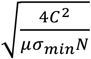, while the bias between meta-analysis and pool analysis ‖log aHR _*meta*_ − log aHR _*pool*_ ‖^2^ is upper bounded with 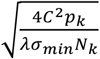. By choosing a proper proximal term coefficient *µ*, we can always have. 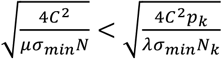, where σ _*min*_ is the minimum eigenvalue of the Hessian Matrix in the optimizations of the CoxPH model.

##### Theorem 2

Our FL method has good convergence with a convergence rate of 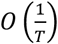 to an approximation of the global optimum, where *T* is the total number of iterations.

The proofs are shown in Supplementary Materials (Supplementary Note 1 and 2).

Several existing studies have addressed federated treatment effect estimation across data from multiple sites.^36,41,82^ Most of these works assume a homogeneous setting,^83–85^ where the covariate distributions are identical across sites. More recently, research has begun to explore federated treatment effect estimation under heterogeneous covariates.^56,57,59,86^ For example, Xiong et al.^57^ proposed federated estimation of average treatment effects (ATEs) across multiple sites by aggregating summary statistics based on propensity scores and outcome models. Their approach emphasizes asymptotic guarantees for estimators under heterogeneous data. Khellaf et al.^59^ studied federated causal inference under randomized controlled trial (RCT) settings, comparing meta-analysis, one-shot, and gradient-based federated estimators of the ATE from the theoretical aspect. However, these works have primarily focused on binary or continuous outcomes and can not be directly applied to time-to-event settings. In contrast, our work investigates federated treatment effect estimation for survival outcomes, a relatively underexplored area, and provides theoretical guarantees demonstrating that our estimator yields less biased results compared to both local-analysis and meta-analysis approaches. Besides, we propose a comprehensive federated target trial emulation framework to estimate real-world treatment effects using EHRs. This includes specification of eligibility criteria, treatment strategies, time zero, follow-up windows, and outcome definitions, which is also underexplored in the literature.

## Data

### INSIGHT

In this study, we selected patients diagnosed with mild cognitive impairment (MCI) between 2006 and 2023 from the INSIGHT network. Eligible patients were required to meet several criteria: they had to be at least 50 years of age at the time of MCI diagnosis, have no history of Alzheimer’s disease (AD) or related dementias in the five years preceding the index date, and have a baseline observation period of at least one year prior to treatment initiation, with no upper limit imposed on this baseline period. The index date was defined as the date of initiation for the study drug, with all inclusion criteria being confirmed by this point. We constructed nine target trials (aspirin, amlodipine, atorvastatin, lidocaine, acetaminophen, famotidine pantoprazole, fluticasone, albuterol).

Treatment initiation was determined as the date of the first prescription of the drug of interest, with at least two consecutive prescriptions within a 30-day window required to confirm valid initiation. Based on baseline eligibility and treatment strategies, patients were assigned to either treatment or comparison groups. We assumed baseline comparability between both groups by adjusting for key covariates, including age, gender, comorbidities, prior medication use, and the time elapsed between MCI diagnosis and treatment initiation. Baseline comorbidities were drawn from the Chronic Conditions Data Warehouse^87^ and other expert-determined risk factors for AD^88,89^, with a total of 64 covariates considered (Supplementary Table 8). These covariates were defined using ICD-9/10 codes, and medication history was constructed from the 200 most frequently prescribed drugs. In total, 267 covariates were adjusted for, including continuous variables such as age and time from MCI diagnosis to treatment initiation, as well as binary variables for gender, comorbidities, and medication use.

Patients were followed from baseline until the earliest of the following events: first AD diagnosis, loss to follow-up, five years after baseline, or the database’s end date. The primary outcome of interest was a newly recorded AD diagnosis during the follow-up period, classified as a positive event. If no AD diagnosis was recorded and the last documented prescription or diagnosis date occurred after the follow-up period ended, the event was classified as negative. Conversely, cases where no AD diagnosis was recorded, but the last prescription or diagnosis date fell before the end of follow-up, were classified as censoring events. The timing of these events was calculated as follows: for positive events, the time was measured from baseline (the initiation date of the drug) to the first AD diagnosis. For negative events, the time corresponded to the total follow-up duration. For censoring events, time was calculated as the interval between baseline and the last recorded prescription or diagnosis date, whichever occurred later. We identified clinical phenotypes relevant to the study based on a set of expert-selected diagnostic codes (Supplementary Table 7). These phenotypes helped refine event classifications and enabled precise tracking of patient outcomes across different trial emulations. This careful differentiation of event types allowed for comprehensive time-to-event analysis across the cohort, ensuring consistency in handling positive, negative, and censoring events.

### eICU-MIMIC^26,39^

We identified suspected infection by the concurrent administration of antibiotics and collection of a body fluid culture. We used a simplified definition of sepsis, classifying any patient with a Sequential Organ Failure Assessment (SOFA) score of 2 or more as having an infectious critical illness, deviating from the Sepsis-3 criterion^90^ of a 2-point increase in SOFA score from baseline. Enrollment for this cohort was defined as the first 24 hours after ICU admission, with patients required to be at least 18 years old and diagnosed with sepsis according to our infectious critical illness definition. Patients with a history of infection or corticosteroid use prior to ICU admission were excluded. See Supplementary Table 5 and 6 for more details on patient characteristics.

We adjusted for a broad array of baseline covariates in the analysis, including vital signs, laboratory measurements, and demographic characteristics, all routinely monitored in ICU settings. These covariates included heart rate, mean arterial pressure, respiratory rate, oxygen saturation, systolic arterial blood pressure, body temperature, and key biochemical, hematological, and physiological markers. Demographic data, such as age, sex, and body mass index (BMI), were also considered, with BMI categorized according to WHO guidelines. We applied the Elixhauser Comorbidity Index^91^ to account for patients’ past medical histories. Data preprocessing involved removing outliers beyond the 99th percentile and imputing missing values using median imputation. The missingness of covariates is shown in Supplementary Table 3. When multiple measurements were available during the 24-hour enrollment window, the worst values were selected to reflect the most severe clinical condition of the patient.

The study’s primary outcome was 28-day mortality, with secondary outcomes including time to ICU discharge and time to cessation of mechanical ventilation. Mechanical ventilation cessation was defined as a 24-hour period without ventilatory support. Competing risk analyses were performed for the secondary outcomes, with death treated as a competing risk^92^. Patients were followed from ICU admission until the first of death, discharge, or loss to follow-up.

## Data Availability

The INSIGHT data can be requested through https://insightcrn.org/. The de-identified data utilized in this study for the development cohort (eICU and MIMIC-IV) can be accessed upon the approval of a formal proposal and the execution of a Data Access Agreement via Physio Net (https://physionet.org/).

## Code Availability

The primary repository is hosted on https://github.com/lihy96/FederatedTrialEmulations. The experiments were conducted using Python 3.10, with survival analysis performed via the lifelines package (version 0.29). All implementation details, including preprocessing scripts, model training, and hyperparameter configurations, are documented within the repository.

## Acknowledgements

F.W. would like to acknowledge the support from NIH awards RF1AG072449, RF1AG084178, R01AG080991, R01AG080624, R01AG076448, R01AG076234, as well as NSF award 1750326 and 2212175.

## Author Contributions

F.W. conceived the initial idea. H.L., C.Z. and W.P. conceived the method and designed the algorithmic techniques. H.L. wrote the codes and performed the computational analysis. C.Z., Z.X. and S.R. preprocessed the INSIGHT and eICU-MIMIC datasets and contributed to the analysis. H.L. drafted the initial manuscript, with critical revisions by F.W. and C.Z. Y.C. reviewed the manuscript and provided suggestions. F.W. supervised the project. All authors reviewed, provided feedback, and approved the final manuscript.

