## Supplementary Materials for "Federated Target Trial Emulation using Distributed Observational Data for Treatment Effect Estimation"

**Table of Content**


- **Supplementary Note 1:** Proof of Theorem 1.
- **Supplementary Note 2:** Proof of Theorem 2 (Convergence Property).
- **Supplementary Table 1:** Federated Target trial emulation  for drug repurposing in treating AD on INSIGHT.
- **Supplementary Table 2**: Federated Target trial emulation for drug repurposing in treating sepsis on eICU-MIMIC.
- **Supplementary Table 3**: Missingness of covariates with the percentage of missing covariates indicated in parentheses of eICU-MIMIC dataset.
- **Supplementary Table 4**: Population characteristics of the five sites in the New York metropolitan area within INSIGHT Clinical Research Networks.
- **Supplementary Table 5**: Patient characteristics of the eICU-MIMIC dataset.
- **Supplementary Table 6**: Patient characteristics comparisons between RI and RW in the eICU-MIMIC cohort.
- **Supplementary Table 7**: Selected ICD-9/10 diagnosis codes for Mild Cognitive Impairment (MCI) and Alzheimer's Disease (AD).
- **Supplementary Table 8**: Baseline comorbidity ICD codes.
- **Supplementary Figure 1**: The estimated aHR and 95% CI on INSIGHT under differential privacy.
- **Supplementary Figure 2**: The estimated aHR and 95% CI on eICU-MIMIC under differential privacy.
- **Supplementary Figure 3**: Sensitivity analysis on different federated learning algorithms on INSIGHT dataset.
- **Supplementary Figure 4**: Sensitivity analysis on different federated learning algorithms on eICU-MIMIC dataset.
- **Supplementary Figure 5**: The estimated aHR and 95% CI on eICU-MIMIC using clone-censor-weight approach to limit immortal time bias.

### Supplementary Notes

#### Supplementary Note 1: Proof of Theorem 1

In federated learning, we have $K$ clients, where each client $k$ holds a local dataset $S_k$, and the total dataset is $S = \bigcup_{k=1}^{K} S_k$. Our goal is to train a global model with parameters $\boldsymbol{\beta}$, minimizing the expected risk over the overall data distribution $D$:

$$R(w) = \mathbb{E}_{z\sim D}[L(\boldsymbol{\beta}; z)].$$

where $L(\boldsymbol{\beta}; z)$ is the loss function.

We consider algorithm stability as the key tool for analyzing the generalization error of our FL model and meta-analysis model. The less sensitive an algorithm is to small changes in the training data, the better its stability.

**Algorithm Stability**: An algorithm $A$ is $\gamma$-stable with respect to a dataset $S$ if for any two datasets $S$ and $S'$ that differ by only one sample, we have:

$$\forall\, z, |\mathbb{E}[L(A(S); z)] - \mathbb{E}[L(A(S'); z)]| \leq \gamma$$

**Generalization Error Bound**: An algorithm $A$ is $\gamma$-stable,then the generalization error bound between the expected risk $R(A(S))$ and empirical risk $\hat{R}\big(A(S)\big)$ and :

$$\mathbb{E}_S\big[(R(A(S))) - \big(\hat{R}(A(S))\big)\big] \leq \gamma.$$

In the following proofs, we would like to first show a tighter generalization error bound of our method compared with meta-analysis by deriving a tighter stability parameter $\gamma$ for our method.

We first analyze the generalization error for our method. Our method introduces a proximal term in the local optimization problem of each client to handle data heterogeneity. Each client $k$ solves the following optimization problem:

$$\min_{\boldsymbol{\beta}} f_k(\boldsymbol{\beta}) + \frac{\mu}{2}\left\|\boldsymbol{\beta} - \boldsymbol{\beta}^{(t)}\right\|^2,$$

where $f_k(\boldsymbol{\beta}) = \frac{1}{N_k}\sum_{z\in S_k} L(\boldsymbol{\beta}; z)$ is the local empirical risk, $\boldsymbol{\beta}^{(t)}$ is the global model parameter (at iteration $t$), and $\mu > 0$ is the coefficient of the proximal term.

For the derivation, we need the following assumptions:

**Assumption 1 (Lipschitz Continuity and Smoothness)**:

$L(\boldsymbol{\beta}; z)$ is $C$-Lipschitz continuous with respect to $\boldsymbol{\beta}$, i.e.,

$$\|L(\boldsymbol{\beta}; z) - L(\boldsymbol{\beta}'; z)\| \leq C\|\boldsymbol{\beta} - \boldsymbol{\beta}'\|.$$

And $L(\boldsymbol{\beta}; z)$ is $\kappa$-smooth with respect to $\boldsymbol{\beta}$, i.e., the second derivative is bounded.

**Assumption 2 ($\lambda$-strongly convex)**: The local optimization $f_k(\boldsymbol{\beta})$ is $\lambda$-strongly convex with respect to $\boldsymbol{\beta}$:

$$\|\boldsymbol{\beta} - \boldsymbol{\beta}'\| \leq \frac{1}{\lambda} \|\nabla f(\boldsymbol{\beta}) - \nabla f(\boldsymbol{\beta}')\|$$

Assume we have two datasets $S$ and $S'$ which differ by only one sample, and this sample belongs to site $k$. We aim to analyze the effect of this change on the global model $\boldsymbol{\beta}$. For client $k$, the local model parameter trained on $S_k$ is $\boldsymbol{\beta}_k$, and the local model parameter trained on $S'_k$ (differing by one sample) is $\boldsymbol{\beta}'_k$. For other clients $j \neq k$, the local model parameters remain unchanged.

Since the loss function is $C$-Lipschitz, the gradient of $L(\boldsymbol{\beta}; z)$ is bounded by $C$. For $S_k$ and $S'_k$ differ by only one sample, the gradient difference is:

$$\|\nabla f_k(\boldsymbol{\beta}_k) - \nabla f_k(\boldsymbol{\beta}'_k)\| \leq \frac{2C}{N_k}$$

Using the strong convexity assumption, we have:

$$\|\boldsymbol{\beta}_k - \boldsymbol{\beta}'_k\| \leq \frac{1}{\lambda} \|\nabla f_k(\boldsymbol{\beta}_k) - \nabla f_k(\boldsymbol{\beta}'_k)\| \leq \frac{2C}{\lambda N_k}$$

The global model is the weighted average of local models:

$$\boldsymbol{\beta} = \textstyle\sum_{k=1}^{K} p_k \boldsymbol{\beta}_k.$$

Thus, the change in the global model is:

$$\|\boldsymbol{\beta} - \boldsymbol{\beta}'\| = p_k \|\boldsymbol{\beta}_k - \boldsymbol{\beta}'_k\| \leq \frac{2Cp_k}{\lambda N_k}.$$

For any sample $z$, we have:

$$\|L(\boldsymbol{\beta}; z) - L(\boldsymbol{\beta}'; z)\| \leq C\|\boldsymbol{\beta} - \boldsymbol{\beta}'\| \leq \frac{2C^2 p_k}{\lambda N_k}.$$

Thus, the stability parameter for meta-analysis is:

$$\gamma_{meta} = \frac{2C^2 p_k}{\lambda N_k}.$$

Since we have the proximal term $\frac{\mu}{2}\left\|\boldsymbol{\beta} - \boldsymbol{\beta}^{(t)}\right\|^2$ for optimization, the local optimization $f_k(\boldsymbol{\beta})$ is $\mu$-strongly convex with respect to $\boldsymbol{\beta}$. Similarly, we have the stability parameter for our method is:

$$\gamma_{FL} = \frac{2C^2 p_k}{\mu N_k} = \frac{2C^2}{\mu N}$$

Typically, we have $\gamma_{FL} < \gamma_{meta}$, due to the following reasons. (1) On the one hand, the strong convexity of our federated learning method comes from the proximal term, while the strong convexity of the meta-analysis method depends on the inherent convexity of the loss function so it can easily achieve $\mu > \lambda$ if $\mu$ is chosen accordingly. On the other hand, data heterogeneity might lead to a small strong convexity parameter $\lambda$ but our method ensures strong convexity through the proximal term with $\mu$. (2) The stability parameter of our method is inversely proportional to the total number of samples, while that of meta-analysis is inversely proportional to the sample size of each client. (3) The more significant of data heterogeneity among sites for meta-analysis, the larger $p_k$. Overall, we have $\gamma_{FL} < \gamma_{meta}$.

Under the CoxPH model and assuming a sufficiently large sample size, we consider a ground-truth pooled model with true parameter vector $\boldsymbol{\beta}^*$. We perform a second-order Taylor expansion of the risk function $L(\boldsymbol{\beta})$ around $\boldsymbol{\beta}^*$:

$$L(\boldsymbol{\beta}) \approx L(\boldsymbol{\beta}^*) + \nabla L(\boldsymbol{\beta}^*)^{\mathrm{T}}(\boldsymbol{\beta} - \boldsymbol{\beta}^*) + \frac{1}{2}(\boldsymbol{\beta} - \boldsymbol{\beta}^*)^{\mathrm{T}}\nabla^2 L(\boldsymbol{\beta}^*)(\boldsymbol{\beta} - \boldsymbol{\beta}^*).$$

Since $\nabla L(\boldsymbol{\beta}^*) = 0$, the expansion simplifies to:

$$L(\boldsymbol{\beta}) \approx L(\boldsymbol{\beta}^*) + \frac{1}{2}(\boldsymbol{\beta} - \boldsymbol{\beta}^*)^{\mathrm{T}}\nabla^2 L(\boldsymbol{\beta}^*)(\boldsymbol{\beta} - \boldsymbol{\beta}^*).$$

According to **Assumption 2**, the Hessian Matrix $\nabla^2 L(\boldsymbol{\beta}^*)$ is positive-definite, so

$$L(\boldsymbol{\beta}) - L(\boldsymbol{\beta}^*) \geq \frac{\sigma_{min}}{2}\|\boldsymbol{\beta} - \boldsymbol{\beta}^*\|^2.$$

where $\sigma_{min}$ is the minimum eigenvalue of the Hessian Matrix $\nabla^2 L(\boldsymbol{\beta}^*)$. the bias of the parameter can be upper bounded by the generalization error:

$$\|\boldsymbol{\beta} - \boldsymbol{\beta}^*\|^2 \leq \frac{2}{\sigma_{min}}(L(\boldsymbol{\beta}) - L(\boldsymbol{\beta}^*)) \leq \frac{2\gamma}{\sigma_{min}}.$$

Finally, the bias of our FL is upper bounded with $\frac{4C^2}{\mu\sigma_{min}N}$, while the bias of meta-analysis is upper bounded with $\frac{4C^2 p_k}{\lambda\sigma_{min}N_k}$, and given a proper $\mu$ we have $\frac{4C^2}{\mu\sigma_{min}N} < \frac{4C^2 p_k}{\lambda\sigma_{min}N_k}$.

Overall, since adjust hazard ratio is calculated $\mathrm{aHR} = \exp(\boldsymbol{\beta})$, we can derive the upper bound of bias of our federeated learning method $\|\mathrm{aHR}_{FL} - \mathrm{aHR}_{pool}\|^2$ is strictly smaller than that of meta-anaysis method $\|\mathrm{aHR}_{meta} - \mathrm{aHR}_{pool}\|^2$. We conclude that our method has a smaller generalization error bound, indicating the effectiveness of our method to estimate the treatment effect with less bias.

**Supplementary Note 2: Proof of Theorem 2 (Convergence Property)**

For convergence analysis of our algorithm, we follow the common **Assumption 1** and **Assumption 2.** Since the loss function is $C$-Lipschitz and convex, the gradient is bounded: $\|\nabla f_k(\boldsymbol{\beta})\| \leq G$. On each site $k$, the local update rule is:

$$\boldsymbol{\beta}_k^{t+1} = \boldsymbol{\beta}_k^t - \eta(\nabla f_k(\boldsymbol{\beta}_k^t) + \mu(\boldsymbol{\beta}_k^t - \boldsymbol{\beta}^t))$$

First, by performing a Taylor expansion of the global objective, according to $\kappa$**-smoothness**, we have

$$f(\boldsymbol{\beta}^{t+1}) \leq f(\boldsymbol{\beta}^{t}) + \nabla f_m(\boldsymbol{\beta}^{t})^T(\boldsymbol{\beta}^{t+1} - \boldsymbol{\beta}^{t}) + \frac{\kappa}{2}||\boldsymbol{\beta}^{t+1} - \boldsymbol{\beta}^{t}||^2$$

Since the local updates in our method contain a regularization term that controls the deviation from the global model, and using the smoothness condition, we can further bound the error term. From the local update rule and the bounded gradient assumption, we obtain:

$$||\boldsymbol{\beta}^{t+1} - \boldsymbol{\beta}^{t}||^2 \leq \eta^2 G^2$$

Considering decrease in global objective function: In each round of federated learning, the expected global objective function $F(\boldsymbol{\beta})$ is supposed to decrease (Here we use $F(\boldsymbol{\beta})$ to denote the log-likelihood function expressed by Eq. (7)). By averaging over all site objective functions $F_k(\boldsymbol{\beta})$ weighted by their data proportions, we have:

$$f(\boldsymbol{\beta}^{t+1}) - f(\boldsymbol{\beta}^{t}) \leq -\eta_t \sum_{k=1}^{K} p_k ||\nabla f_k(\boldsymbol{\beta}^{t})||^2 + \frac{\kappa\eta^2}{2} \sum_{k=1}^{K} p_k ||\nabla f_k(\boldsymbol{\beta}^{t})||^2$$

To ensure convergence, the learning rate $\eta$ needs to be appropriately selected. A common choice is $\eta = \frac{1}{\sqrt{T}}$. If the conditions mentioned are satisfied, our algorithm can converge at a rate of $O(\frac{1}{T})$to an approximation of the global optimum, where $T$ is the total number of iterations.

$$f(\boldsymbol{\beta}^{T}) - f(\boldsymbol{\beta}^{*}) \leq \frac{1}{T}(f(\boldsymbol{\beta}^{0}) - f(\boldsymbol{\beta}^{*}) + \frac{\kappa G^2 T \eta^2}{2})$$

So,

$$f(\boldsymbol{\beta}^{T}) - f(\boldsymbol{\beta}^{*}) \leq O\left(\frac{1}{T}\right).$$

Overall, by introducing a consistency penalty between local and global models, our algorithm effectively addresses the issue of data heterogeneity across sites and ensures convergence under certain conditions with a promising convergence rate.

### Supplementary Tables

**Supplementary Table 1: Federated Target trial emulation for drug repurposing in treating AD on INSIGHT**

| Protocol component | Target trial specification | Federated Target trial emulation |
|---|---|---|
| **Eligibility criteria** | Patients with MCI.<br>Age ≥ 50 at MCI diagnosis.<br>No history of AD or dementia before baseline.<br>No trial drug prescription before baseline.<br>The baseline is the date when all eligibility criteria are met. | Same as for the target trial.<br>We define MCI diagnosis according to the selected ICD-9/10 codes between January 2010 and November 2023 in Site 1, Febrary 2007 and December 2023 in Site 2, August 2006 to November 2023 in Site 3, May 2010 and November 2023 in Site 4, January 2010 and June 2020 in Site 5.<br>We require at least one year of data prior to the index date, with no upper limit, for each individual.<br>We require no AD or related dementia five years before the index date.<br>The first MCI diagnosis is required to occur before the initiation of the trial drug.<br>The index date is defined as the date of the first trial drug prescription, at which point all eligibility criteria had to be fulfilled. |
| **Treatment strategies** | Strategy 0: Initiation of an alternative drug at baseline. The alternative drug means a similar drug within the same therapeutic class (e.g., the second-level Anatomical Therapeutic Chemical classification[1]).<br><br>Strategy 1: Initiation of the trial drug at baseline. | Same as for the target trial.<br>The drug initiation date is defined as the first recorded prescription of the trial drug. A valid initiation required at least two prescriptions, with a minimum interval of one month between the initiation date and the second prescription. |
| **Treatment assignment** | At baseline, patients are randomly assigned to one of the treatment strategies, with full awareness of the strategy they have been assigned to. | Patients are classified into different arms based on their baseline eligibility criteria and assigned treatment strategy.<br>We emulate randomization by applying inverse probability of treatment weighting (IPTW) based on baseline covariates, including age, gender, comorbidities, medications, and the time lag between MCI diagnosis and the index date, to ensure conditional exchangeability between the treated and control groups. |
| **Outcomes** | AD onset | Same as for the target trial.<br>We define the outcome of incident AD based on selected ICD-9/10 diagnosis codes during the follow-up period. |
| **Follow-up** | Time zero is the date of the first trial drug prescription, at which point all eligibility criteria had to be fulfilled.<br><br>Each patient was followed from their baseline date until the | Same as for the target trial. |

| | | |
|---|---|---|
| | earliest of the following events: their first AD diagnosis, loss to follow-up, or five years after baseline. | |
| **Causal contrasts** | Intention-to-treat effect | Observational analog of intention-to-treat effect. |
| **Statistical analysis** | Intention-to-treat analysis is conducted as a time-to-first event approach, applying inverse probability of treatment weighting (IPTW) to adjust for baseline covariates.<br><br>We use the Cox proportional hazards model to report adjusted hazard ratios. | Same as for the target trial.<br>To protect the privacy, we only share summary-level machine learning model information among hospitals without aggregating patient-level data. |

**Supplementary Table 2: Federated Target trial emulation for drug repurposing in treating sepsis on eICU-MIMIC with 28-day mortality outcome.**

| Protocol component | Target trial specification | Federated Target trial emulation |
|---|---|---|
| **Eligibility criteria** | Age >=18 at enrollment window<br>Patient with sepsis at enrollment window<br>No sepsis history before enrollment window<br>No corticosteroid prescription more than 10 hrs before enrollment window<br>Sepsis is defined based on Sepsis-3 criteria<br>The enrollment window is defined as 24 hours after the ICU admission [0h, 24h]<br>On mechanical ventilation at enrollment window (only for analyzing cessation of mechanical ventilation outcome) | Age >=18 at enrollment window<br>We require the first ICU stay if he/she has multiple ICU stays in the database<br>No sepsis history before enrollment window<br>We define sepsis based on Sepsis-3 with the record at his/her enrollment window<br>The enrollment window is defined as the first day of the patient's ICU admission<br>On mechanical ventilation at enrollment window (only for analyzing cessation of mechanical ventilation outcome) |
| **Treatment strategies** | Strategy 0: No initiation of any corticosteroids drug at enrollment window<br><br>Strategy 1: Initiation of hydrocortisone at a dose of 160 mg at enrollment window | We compute cumulative milligram dosing of hydrocortisone at enrollment window, and if a patient received more than 160 mg per day hydrocortisone equivalent, they are denoted as having hydrocortisone exposure.<br><br>We consider corticosteroids including Prednisolone, Prednisone, Hydrocortisone, Dexamethasone, and Methylprednisolone.<br><br>**Conversions:**<br>1 mg Prednisolone = 4 mg Hydrocortisone<br>1 mg Prednisone = 4 mg Hydrocortisone<br>1 mg Dexamethasone = 25 mg Hydrocortisone |

| | | |
|---|---|---|
| | | 1 mg Methylprednisolone = 5 mg Hydrocortisone |
| **Treatment assignment** | Patients are randomly assigned to either treatment strategy on the first day after ICU admission and are aware of the strategy they are assigned to. | We classified individuals according to the strategy that their data were compatible with at the enrollment window and attempted to emulate randomization by adjusting for baseline confounders. |
| **Outcomes** | Primary outcome:<br>28-day mortality from time of randomization<br><br>Secondary outcomes:<br>Time to ICU Discharge from time of randomization<br><br>Time to ventilation cessation from time of randomization, ventilation cessation is defined as 24 hours of no ventilation support | Same as for the target trial. |
| **Follow-up** | Time zero is defined as the time of treatment initiation (receiving ≥ 160 mg/day hydrocortisone equivalent during the enrollment window) for treated patients, and as the time of ICU admission for untreated patients.<br><br>We follow each patient from his/her baseline until the day of his/her death, loss to follow-up, or discharge, whichever occurs first. | Same as for the target trial. |
| **Causal contrasts** | Intention-to-treat effect | Observational analog of intention-to-treat effect |
| **Statistical analysis** | Intention-to-treat analysis is conducted as a time-to-first event approach, applying inverse probability of treatment weighting (IPTW) to adjust for baseline covariates.<br><br>We use the Cox proportional hazards model to report adjusted hazard ratios. | Same as for the target trial.<br><br>Patients whose transfer location are unknown are censored at time transfer<br><br>To protect the privacy, we only share summary-level machine learning model information among hospitals without aggregating patient-level data. |

**Supplementary Table 3: Missingness of covariates with the percentage of missing covariates indicated in parentheses of eICU-MIMIC dataset.**

| Covariates | eICU | MIMIC-IV |
|---|---|---|
| **ALT** | 2913.0 (0.20) | 2957.0 (0.48) |
| **AST** | 2894.0 (0.20) | 2970.0 (0.48) |
| **Albumin** | 2722.0 (0.19) | 3509.0 (0.57) |
| **BMI** | 0.0 (0.00) | 0.0 (0.00) |
| **BUN** | 325.0 (0.02) | 111.0 (0.02) |
| **Bands** | 10962.0 (0.76) | 4962.0 (0.81) |
| **Bilirubin** | 3008.0 (0.21) | 2997.0 (0.49) |
| **CRP** | 13712.0 (0.95) | Not Available |
| **Chloride** | 391.0 (0.03) | 109.0 (0.02) |
| **Creatinine** | 328.0 (0.02) | 82.0 (0.01) |

| | | |
|---|---|---|
| **FiO2** | 10959.0 (0.76) | 1079.0 (0.18) |
| **GCS** | 3787.0 (0.26) | 2.0 (0.00) |
| **Glucose** | 419.0 (0.03) | 152.0 (0.02) |
| **Heart rate** | 1170.0 (0.08) | 122.0 (0.02) |
| **Hemoglobin** | 442.0 (0.03) | 120.0 (0.02) |
| **INR** | 4607.0 (0.32) | 316.0 (0.05) |
| **Lactate** | 4103.0 (0.29) | 1076.0 (0.18) |
| **Lymphocyte percent** | 3179.0 (0.22) | Not Available |
| **MAP** | 13891.0 (0.97) | 121.0 (0.02) |
| **Pa02** | 4596.0 (0.32) | 1298.0 (0.21) |
| **Platelet** | 476.0 (0.03) | 126.0 (0.02) |
| **Respiratory rate** | 1297.0 (0.09) | 106.0 (0.02) |
| **SO2** | 2538.0 (0.18) | 62.0 (0.01) |
| **Sodium** | 308.0 (0.02) | 116.0 (0.02) |
| **Systolic ABP** | 1308.0 (0.09) | 1918.0 (0.31) |
| **Temperature** | 610.0 (0.04) | 4647.0 (0.76) |
| **Troponin I** | 8055.0 (0.56) | Not Available |
| **Troponin T** | 13684.0 (0.95) | 4143.0 (0.68) |
| **Urine** | 4323.0 (0.30) | 200.0 (0.03) |
| **WBC** | 443.0 (0.03) | 125.0 (0.02) |
| **Age** | 118.0 (0.01) | 232.0 (0.04) |

**Supplementary Table 4: Population characteristics of the five sites in the New York metropolitan area within five INSIGHT sites.**

| **Site 1, January 2010 to November 2023, 889,550 patients** | | | | |
|---|---|---|---|---|
| | **MCI** | **AD** | **MCI \ AD** | **P-value**[a] |
| No. of patients | 5803 (100%) | 785 (13.53%) | 5018 (86.47%) | – |
| MCI age, median (IQR)[c] | 71 (53, 80) | 77 (72, 83) | 68 (49, 79) | 0.000 |
| Sex-female | 3069 (52.89%) | 476 (60.64%) | 2593 (51.67%) | 0.000[b] |
| Sex-male | 2734 (47.11%) | 309 (39.36%) | 2425 (48.33%) | – |
| Antidiabetic medication | 2196 (37.84%) | 347 (44.20%) | 1849 (36.85%) | 0.0001 |
| Antihypertensives medication | 869 (14.98%) | 95 (12.10%) | 774 (15.42%) | 0.0177 |
| Alcohol Use Disorders | 427 (7.36%) | 48 (6.11%) | 379 (7.55%) | 0.1733 |
| Anxiety Disorders | 2190 (37.74%) | 318 (40.51%) | 1872 (37.31%) | 0.0925 |
| Depression | 2103 (36.24%) | 337 (42.93%) | 1766 (35.19%) | 0.0000 |
| Diabetes | 1901 (32.76%) | 296 (37.71%) | 1605 (31.98%) | 0.0017 |
| Heart Failure | 1292 (22.26%) | 184 (23.44%) | 1108 (22.08%) | 0.4208 |
| Hyperlipidemia | 3245 (55.92%) | 557 (70.96%) | 2688 (53.57%) | 0.0000 |
| Hypertension | 3735 (64.36%) | 588 (74.90%) | 3147 (62.71%) | 0.0000 |
| Ischemic Heart Disease | 1951 (33.62%) | 322 (41.02%) | 1629 (32.46%) | 0.0000 |
| Obesity | 1260 (21.71%) | 155 (19.75%) | 1105 (22.02%) | 0.1641 |
| Stroke/Transient Ischemic Attack | 1477 (25.45%) | 225 (28.66%) | 1252 (24.95%) | 0.0295 |
| Tobacco Use | 665 (11.46%) | 61 (7.77%) | 604 (12.04%) | 0.0006 |
| Traumatic Brain Injury | 217 (3.74%) | 21 (2.68%) | 196 (3.91%) | 0.1121 |
| Sleep disorders | 1845 (31.79%) | 272 (34.65%) | 1573 (31.35%) | 0.0708 |
| Periodontitis | 209 (3.60%) | 32 (4.08%) | 177 (3.53%) | 0.5062 |
| Menopause | 17 (0.29%) | 3 (0.38%) | 14 (0.28%) | 0.8869 |
| **Site 2, Febrary 2007 to December 2023, 992,800 patients** | | | | |
| | **MCI** | **AD** | **MCI \ AD** | **P-value**[a] |
| No. of patients | 4764 (100%) | 680 (14.27%) | 4084 (85.73%) | – |
| MCI age, median (IQR)[c] | 73 (61, 80) | 78 (72, 84) | 71 (58, 79) | 0.000 |
| Sex-female | 2933 (61.57%) | 473 (69.56%) | 2460 (60.24%) | 0.000[b] |
| Sex-male | 1831 (38.43%) | 207 (30.44%) | 1624 (39.76%) | – |
| Antidiabetic medication | 2915 (61.19%) | 497 (73.09%) | 2418 (59.21%) | 0.0000 |

| | | | | |
|---|---|---|---|---|
| Antihypertensives medication | 1357 (28.48%) | 234 (34.41%) | 1123 (27.50%) | 0.0003 |
| Alcohol Use Disorders | 603 (12.66%) | 70 (10.29%) | 533 (13.05%) | 0.0524 |
| Anxiety Disorders | 1914 (40.18%) | 286 (42.06%) | 1628 (39.86%) | 0.2987 |
| Depression | 2184 (45.84%) | 389 (57.21%) | 1795 (43.95%) | 0.0000 |
| Diabetes | 2505 (52.58%) | 425 (62.50%) | 2080 (50.93%) | 0.0000 |
| Heart Failure | 1354 (28.42%) | 221 (32.50%) | 1133 (27.74%) | 0.0124 |
| Hyperlipidemia | 3480 (73.05%) | 594 (87.35%) | 2886 (70.67%) | 0.0000 |
| Hypertension | 3996 (83.88%) | 653 (96.03%) | 3343 (81.86%) | 0.0000 |
| Ischemic Heart Disease | 2175 (45.65%) | 386 (56.76%) | 1789 (43.81%) | 0.0000 |
| Obesity | 1915 (40.20%) | 268 (39.41%) | 1647 (40.33%) | 0.6825 |
| Stroke/Transient Ischemic Attack | 1224 (25.69%) | 212 (31.18%) | 1012 (24.78%) | 0.0005 |
| Tobacco Use | 920 (19.31%) | 97 (14.26%) | 823 (20.15%) | 0.0004 |
| Traumatic Brain Injury | 122 (2.56%) | 17 (2.50%) | 105 (2.57%) | 1.0000 |
| Sleep disorders | 1744 (36.61%) | 281 (41.32%) | 1463 (35.82%) | 0.0066 |
| Periodontitis | 39 (0.82%) | 3 (0.44%) | 36 (0.88%) | 0.3421 |
| Menopause | 61 (1.28%) | 10 (1.47%) | 51 (1.25%) | 0.7702 |
| **Site 3, August 2006 to November 2023, 1,354,013 patients** | | | | |
| | **MCI** | **AD** | **MCI \ AD** | **P-value**[a] |
| No. of patients | 4764 (100%) | 680 (14.27%) | 4084 (85.73%) | – |
| MCI age, median (IQR)[c] | 73 (61, 80) | 78 (72, 84) | 71 (58, 79) | 0.000 |
| Sex-female | 2933 (61.57%) | 473 (69.56%) | 2460 (60.24%) | 0.000[b] |
| Sex-male | 1831 (38.43%) | 207 (30.44%) | 1624 (39.76%) | – |
| Antidiabetic medication | 914 (19.19%) | 112 (16.47%) | 802 (19.64%) | 0.0000 |
| Antihypertensives medication | 1974 (41.44%) | 312 (45.88%) | 1662 (40.70%) | 0.0003 |
| Alcohol Use Disorders | 1451 (30.46%) | 197 (28.97%) | 1254 (30.71%) | 0.0524 |
| Anxiety Disorders | 86 (1.81%) | 12 (1.76%) | 74 (1.81%) | 0.2987 |
| Depression | 2915 (61.19%) | 497 (73.09%) | 2418 (59.21%) | 0.0000 |
| Diabetes | 1357 (28.48%) | 234 (34.41%) | 1123 (27.50%) | 0.0000 |
| Heart Failure | 603 (12.66%) | 70 (10.29%) | 533 (13.05%) | 0.0124 |
| Hyperlipidemia | 1914 (40.18%) | 286 (42.06%) | 1628 (39.86%) | 0.0000 |
| Hypertension | 2184 (45.84%) | 389 (57.21%) | 1795 (43.95%) | 0.0000 |
| Ischemic Heart Disease | 2505 (52.58%) | 425 (62.50%) | 2080 (50.93%) | 0.0000 |
| Obesity | 1354 (28.42%) | 221 (32.50%) | 1133 (27.74%) | 0.6825 |
| Stroke/Transient Ischemic Attack | 3480 (73.05%) | 594 (87.35%) | 2886 (70.67%) | 0.0005 |
| Tobacco Use | 3996 (83.88%) | 653 (96.03%) | 3343 (81.86%) | 0.0004 |
| Traumatic Brain Injury | 2175 (45.65%) | 386 (56.76%) | 1789 (43.81%) | 1.0000 |
| Sleep disorders | 1915 (40.20%) | 268 (39.41%) | 1647 (40.33%) | 0.0066 |
| Periodontitis | 1224 (25.69%) | 212 (31.18%) | 1012 (24.78%) | 0.3421 |
| Menopause | 920 (19.31%) | 97 (14.26%) | 823 (20.15%) | 0.7702 |
| **Site 4, May 2010 to November 2023, 1,117,759 patients** | | | | |
| | **MCI** | **AD** | **MCI \ AD** | **P-value**[a] |
| No. of patients | 10926 (100%) | 1320 (12.08%) | 9606 (87.92%) | – |
| MCI age, median (IQR)[c] | 72 (58, 80) | 79 (73, 84) | 70 (56, 79) | 0.000 |
| Sex-female | 5942 (54.38%) | 771 (58.41%) | 5171 (53.83%) | 0.000[b] |
| Sex-male | 4984 (45.62%) | 549 (41.59%) | 4435 (46.17%) | – |
| Antidiabetic medication | 7602 (69.58%) | 1025 (77.65%) | 6577 (68.47%) | 0.3947 |
| Antihypertensives medication | 1646 (15.06%) | 150 (11.36%) | 1496 (15.57%) | 0.0009 |
| Alcohol Use Disorders | 1153 (10.55%) | 107 (8.11%) | 1046 (10.89%) | 0.0033 |
| Anxiety Disorders | 483 (4.42%) | 36 (2.73%) | 447 (4.65%) | 0.4723 |
| Depression | 4858 (44.46%) | 572 (43.33%) | 4286 (44.62%) | 0.0068 |
| Diabetes | 2622 (24.00%) | 268 (20.30%) | 2354 (24.51%) | 0.0006 |
| Heart Failure | 559 (5.12%) | 45 (3.41%) | 514 (5.35%) | 0.0000 |

| | | | | |
|---|---|---|---|---|
| Hyperlipidemia | 4856 (44.44%) | 574 (43.48%) | 4282 (44.58%) | 0.0000 |
| Hypertension | 4512 (41.30%) | 591 (44.77%) | 3921 (40.82%) | 0.0000 |
| Ischemic Heart Disease | 3526 (32.27%) | 481 (36.44%) | 3045 (31.70%) | 0.0000 |
| Obesity | 2275 (20.82%) | 334 (25.30%) | 1941 (20.21%) | 0.0000 |
| Stroke/Transient Ischemic Attack | 7983 (73.06%) | 1144 (86.67%) | 6839 (71.20%) | 0.0000 |
| Tobacco Use | 7833 (71.69%) | 1115 (84.47%) | 6718 (69.94%) | 0.0000 |
| Traumatic Brain Injury | 5426 (49.66%) | 780 (59.09%) | 4646 (48.37%) | 0.0000 |
| Sleep disorders | 3316 (30.35%) | 326 (24.70%) | 2990 (31.13%) | 0.0882 |
| Periodontitis | 3438 (31.47%) | 503 (38.11%) | 2935 (30.55%) | 0.6691 |
| Menopause | 1238 (11.33%) | 101 (7.65%) | 1137 (11.84%) | 0.0832 |
| **Site 5, January 2010 to June 2020, 1,178,306 patients** | | | | |
| | **MCI** | **AD** | **MCI \ AD** | **P-value**[a] |
| No. of patients | 7272 (100%) | 1243 (17.09%) | 6029 (82.91%) | – |
| MCI age, median (IQR)[c] | 74 (63, 82) | 79 (74, 84) | 73 (60, 71) | 0.000 |
| Sex-female | 3987 (54.83%) | 721 (58.00%) | 3266 (54.17%) | 0.000[b] |
| Sex-male | 3285 (45.17%) | 522 (42.00%) | 2763 (45.83%) | – |
| Antidiabetic medication | 4166 (57.29%) | 829 (66.69%) | 3337 (55.35%) | 0.4432 |
| Antihypertensives medication | 1837 (25.26%) | 269 (21.64%) | 1568 (26.01%) | 0.0107 |
| Alcohol Use Disorders | 932 (12.82%) | 96 (7.72%) | 836 (13.87%) | 0.2200 |
| Anxiety Disorders | 328 (4.51%) | 47 (3.78%) | 281 (4.66%) | 0.6264 |
| Depression | 2234 (30.72%) | 370 (29.77%) | 1864 (30.92%) | 0.4388 |
| Diabetes | 885 (12.17%) | 124 (9.98%) | 761 (12.62%) | 0.7367 |
| Heart Failure | 438 (6.02%) | 65 (5.23%) | 373 (6.19%) | 0.0702 |
| Hyperlipidemia | 2932 (40.32%) | 493 (39.66%) | 2439 (40.45%) | 0.0000 |
| Hypertension | 2601 (35.77%) | 457 (36.77%) | 2144 (35.56%) | 0.0000 |
| Ischemic Heart Disease | 2098 (28.85%) | 364 (29.28%) | 1734 (28.76%) | 0.0000 |
| Obesity | 1379 (18.96%) | 259 (20.84%) | 1120 (18.58%) | 0.0000 |
| Stroke/Transient Ischemic Attack | 5052 (69.47%) | 977 (78.60%) | 4075 (67.59%) | 0.0000 |
| Tobacco Use | 4867 (66.93%) | 896 (72.08%) | 3971 (65.86%) | 0.0000 |
| Traumatic Brain Injury | 2599 (35.74%) | 518 (41.67%) | 2081 (34.52%) | 0.0567 |
| Sleep disorders | 1302 (17.90%) | 153 (12.31%) | 1149 (19.06%) | 0.7166 |
| Periodontitis | 2034 (27.97%) | 418 (33.63%) | 1616 (26.80%) | 0.0642 |
| Menopause | 634 (8.72%) | 68 (5.47%) | 566 (9.39%) | 0.5848 |

[a] Two-sided T-test for the null hypothesis that two independent samples (population with AD diagnosis v.s. population without any AD diagnosis) have identical average values, except for sex [b] Chi-square test of independence of the observed male and female frequencies.
[c] MCI age is the sample median with inter-quartile range (IQR, 25th to 75th percentile).

**Supplementary Table 5: Patient characteristics of the eICU-MIMIC dataset.**

| **Characteristics** | **Total** | **Treated N** | **Untreated** | **p-value** |
|---|---|---|---|---|
| No. of patients | 21458 | 1834 | 19624 | |
| Age, median (IQR) | 67 (55, 77) | 66 (55, 76) | 67 (55, 77) | 0.064 |
| Sex, No. (%) | | | | <0.001 |
| Male | 12068 (56.2%) | 937 (51.1%) | 11131 (56.7%) | |
| Female | 9388 (43.8%) | 897 (48.9%) | 8491 (43.3%) | |
| Ethnicity, No. (%) | | | 0.004 | |
| White | 15651 (72.9%) | 1394 (76.0%) | 14257 (72.7%) | |
| Black | 1791 (8.3%) | 125 (6.8%) | 1666 (8.5%) | |
| Other | 4016 (18.7%) | 315 (17.2%) | 3701 (18.9%) | |
| Elixhauser index, median (IQR) | 5.0 [0.0 - 13.0] | 6.0 [0.0 - 15.0] | 5.0 [0.0 - 12.0] | <0.001 |
| Length stay, median (IQR) | 5.7 [2.8 - 10.6] | 5.7 [2.9 - 10.0] | 5.7 [2.7 - 10.7] | 0.08 |
| Mechanical ventilation at admission, No. (%) | 14356 (66.9%) | 1366 (74.5%) | 12990 (66.2%) | <0.001 |
| Baseline SOFA, mean (SD) | 7.2 (3.8) | 7.3 (4.1) | 7.3 (3.8) | 0.335 |

| | | | | |
|---|---|---|---|---|
| Infection, No. (%) | | | | |
| Central nervous system | 130 (0.6%) | 13 (0.7%) | 117 (0.6%) | 0.662 |
| Intra-abdominal | 1870 (8.7%) | 91 (5.0%) | 1779 (9.1%) | <0.001 |
| Pneumonia | 6495 (30.3%) | 851 (46.4%) | 5644 (28.8%) | <0.001 |
| Septicemia bacteremia | 9778 (45.6%) | 798 (43.5%) | 8980 (45.8%) | 0.069 |
| Skin soft tissue | 1174 (5.5%) | 77 (4.2%) | 1097 (5.6%) | 0.014 |
| Urinary tract | 3760 (17.5%) | 194 (10.6%) | 3566 (18.2%) | <0.001 |

**Supplementary Table 6: Patient characteristics comparisons between RI and RW cohorts from eICU+MIMIC.** Median and IQR is shown for select covariates. P-value(s) are determined by student's t-test.

| Covariate, Median (IQR) | Rapidly Improving, n = 7208 | Rapidly Worsening, n = 7359 | p-value |
|---|---|---|---|
| **Albumin** | 3.0, [2.9, 3.0] | 2.9, [2.5, 3.3] | <0.001 |
| **ALT** | 49.0, [25.0, 64.54] | 45.0, [23.0, 100.45] | <0.001 |
| **AST** | 68.0, [35.0, 103.57] | 67.0, [29.0, 166.29] | <0.001 |
| **Bilirubin** | 0.9, [0.5, 0.94] | 1.3, [0.6, 1.6] | <0.001 |
| **BUN** | 21.0, [15.0, 37.0] | 34.0, [22.0, 50.0] | <0.001 |
| **Chloride** | 109.0, [106.0, 112.0] | 105.6, [101.0, 110.0] | <0.001 |
| **Creatinine** | 1.1, [0.8, 1.82] | 1.77, [1.12, 2.65] | <0.001 |
| **FiO2** | 65.45, [65.45, 100.0] | 68.58, [68.58, 68.58] | <0.001 |
| **Glucose** | 142.0, [117.0, 180.81] | 166.0, [130.0, 217.0] | <0.001 |
| **Heart Rate** | 100.0, [89.0, 111.0] | 121.22, [108.0, 133.5] | <0.001 |
| **Hemoglobin** | 9.8, [8.5, 11.2] | 10.14, [8.6, 11.6] | <0.001 |
| **INR** | 1.3, [1.2, 1.4] | 1.88, [1.3, 1.88] | <0.001 |
| **MAP** | 58.19, [56.0, 59.0] | 54.23, [54.23, 54.23] | <0.001 |
| **Lactate** | 2.19, [1.6, 2.8] | 4.11, [2.3, 4.71 | <0.001 |
| **GCS** | 4.0, [3.0, 8.01 | 13.0, [12.95, 15.0] | <0.001 |
| **Pa02** | 97.49, [76.0, 117.0] | 78.27, [64.0, 78.27] | <0.001 |
| **Platelet** | 171.0, [127.0, 226.0] | 156.0, [104.0, 205.0] | <0.001 |
| **Respiratory Rate** | 26.0, [22.0, 29.0] | 34.0, [28.0, 38.0] | <0.001 |
| **SO2** | 93.01, [92.0, 96.0] | 87.0, [86.0, 92.0] | <0.001 |
| **Sodium** | 140.0, [138.0, 142.0] | 139.0, [136.0, 142.0] | <0.001 |
| **Systolic ABP** | 88.92, [82.0, 95.01 | 83.0, [73.0, 91.0] | <0.001 |
| **Temperature** | 37.61, [37.5, 37.71 | 37.5, [37.05, 38.11] | 0.006 |
| **Troponin T** | 0.23, [0.16, 0.66] | 0.2, [0.2, 0.2] | <0.001 |
| **Urine** | 1765.0, [1300.0, 2599.25] | 1303.78, [650.0, 1521.0] | <0.001 |
| **White Blood Cell Count** | 15.1, [11.5, 19.1] | 15.0, [10.1, 20.0] | 0.629 |
| **Comorbidity Score** | 0.0, [0.0, 10.0] | 9.0, [0.0, 18.0] | <0.001 |

**Supplementary Table 7: Selected ICD-9/10 diagnosis codes for Mild Cognitive Impairment (MCI) and Alzheimer's Disease (AD).**

| | |
|---|---|
| **MCI** | **Usage**: The definition of MCI in real-world healthcare data for the selection of the targeted population.<br><br>**ICD-9 codes**:<br>331.83 Mild cognitive impairment, so stated<br>294.9 Unspecified persistent mental disorders due to conditions classified elsewhere<br>**ICD-10 codes**:<br>G31.84 Mild cognitive impairment, so stated<br>F09 Unspecified mental disorder due to known physiological condition<br>To select patients with any of the above codes in the database |

| | |
|---|---|
| **AD** | **Usage**: The definition of AD in real-world healthcare data for the selection of eligible individuals before baseline and identification of outcome in follow-up.<br><br>**ICD-9 codes**:<br>331.0 Alzheimer's disease<br>**ICD-10 codes**:<br>G30 Alzheimer's disease<br>G30.0 Alzheimer's disease with early onset<br>G30.1 Alzheimer's disease with late onset<br>G30.8 Other Alzheimer's disease<br>G30.9 Alzheimer's disease, unspecified<br>To select patients with any of the above codes in the database |
| **AD-related dementias** | **Usage**: The definition of AD-related dementias in real-world healthcare data for selection of eligible individuals before baseline.<br><br>**ICD-9 codes**:<br>294.10 Dementia in conditions classified elsewhere without behavioral disturbance<br>294.11 Dementia in conditions classified elsewhere with behavioral disturbance<br>294.20 Dementia, unspecified, without behavioral disturbance.<br>294.21 Dementia, unspecified, with behavioral<br>disturbance 290.* Dementias<br>**ICD-10 codes**:<br>F01.* Vascular dementia<br>F02.* Dementia in other diseases classified<br>elsewhere F03.* Unspecified dementia<br>To select patients with any of the above codes in the database |

MCI, mild cognitive impairment; AD, Alzheimer's disease; ICD-9/10, the International Classification of Diseases 9th or 10th Revision

**Supplementary Table 8: Baseline comorbidity ICD codes**

| Comorbidities | ICD codes |
|---|---|
| ADHD, Conduct Disorders, and Hyperkinetic Syndrome | 31200, 31201, 31202, 31203, 31210, 31211, 31212, 31213, 31220, 31221, 31222, 31223, 31230, 31231, 31232, 31233, 31234, 31235, 31239, 3124, 31281, 31282, 31289, 3129, 31400, 31401, 3141, 3142, 3148, 3149, F630, F631, F632, F633, F6381, F6389, F639, F900, F901, F902, F908, F909, F910, F911, F912, F913, F918, F919 |
| Acquired Hypothyroidism | 2440, 2441, 2442, 2443, 2448, 2449, E018, E02, E032, E033, E038, E039, E890 |
| Acute Myocardial Infarction | 41001, 41011, 41021, 41031, 41041, 41051, 41061, 41071, 41081, 41091, I2101, I2102, I2109, I2111, I2119, I2121, I2129, I213, I214, I219, I21A1, I21A9, I220, I221, I222, I228, I229 |
| Alcohol Use Disorders | 2910, 2911, 2912, 2913, 2914, 2915, 2918, 29181, 29182, 29189, 2919, 30300, 30301, 30302, 30390, 30391, 30392, 30500, 30501, 30502, 3575, 4255, 53530, 53531, 5710, 5711, 5712, 5713, 76071, 9800, E8600, F1010, F10120, F10121, F10129, F1014, F10150, F10151, F10159, F10180, F10181, F10182, F10188, F1019, F1020, F10220, F10221, F10229, F10230, F10231, F10232, F10239, F1024, F10250, F10251, F10259, F1026, F1027, F10280, F10281, F10282, F10288, F1029, F10920, F10921, F10929, F1094, F10950, F10951, F10959, F1096, F1097, F10980, F10981, F10982, F10988, F1099, G621, I426, K2920, K2921, K700, K7010, K7011, K702, K7030, K7031, K7040, K7041, K709, P043, Q860, T510X1A, T510X2A, T510X3A, T510X4A, V6542, V791, Z7141, Z7142 |

| | |
|---|---|
| Anemia | 2800, 2801, 2808, 2809, 2810, 2811, 2812, 2813, 2814, 2818, 2819, 2820, 2821, 2822, 2823, 28240, 28241, 28242, 28243, 28244, 28245, 28246, 28247, 28249, 2825, 28260, 28261, 28262, 28263, 28264, 28268, 28269, 2827, 2828, 2829, 2830, 28310, 28311, 28319, 2832, 2839, 28401, 28409, 28411, 28412, 28419, 2842, 28481, 28489, 2849, 2850, 2851, 28521, 28522, 28529, 2853, 2858, 2859, D500, D501, D508, D509, D510, D511, D512, D513, D518, D519, D520, D521, D528, D529, D530, D531, D532, D538, D539, D550, D551, D552, D553, D558, D559, D560, D561, D562, D563, D564, D565, D568, D569, D5700, D5701, D5702, D571, D5720, D57211, D57212, D57219, D573, D5740, D57411, D57412, D57419, D5780, D57811, D57812, D57819, D580, D581, D582, D588, D589, D590, D591, D592, D593, D594, D595, D596, D598, D599, D600, D601, D608, D609, D6101, D6109, D611, D612, D613, D61810, D61811, D61818, D6182, D6189, D619, D62, D630, D631, D638, D640, D641, D642, D643, D644, D6481, D6489, D649 |
| Anxiety Disorders | 29384, 30000, 30001, 30002, 30009, 30010, 30020, 30021, 30022, 30023, 30029, 3003, 3005, 30089, 3009, 3080, 3081, 3082, 3083, 3084, 3089, 30981, 3130, 3131, 31321, 31322, 3133, 31382, 31383, F064, F4000, F4001, F4002, F4010, F4011, F40210, F40218, F40220, F40228, F40230, F40231, F40232, F40233, F40240, F40241, F40242, F40243, F40248, F40290, F40291, F40298, F408, F409, F410, F411, F413, F418, F419, F42, F422, F423, F424, F428, F429, F430, F4310, F4311, F4312, F449, F458, F488, F489, F938, F99, R452, R455, R456, R457 |
| Asthma | 49300, 49301, 49302, 49310, 49311, 49312, 49320, 49321, 49322, 49381, 49382, 49390, 49391, 49392, J4520, J4521, J4522, J4530, J4531, J4532, J4540, J4541, J4542, J4550, J4551, J4552, J45901, J45902, J45909, J45990, J45991, J45998 |
| Atrial Fibrillation | 42731, I480, I481, I482, I4891 |
| Autism Spectrum Disorders | 2990, 29900, 29901, 2991, 29911, 2998, 29980, 29981, 2999, 29990, 29991, F840, F843, F845, F848, F849 |
| Benign Prostatic Hyperplasia | 60000, 60001, 60010, 60011, 60020, 60021, 6003, 60090, 60091, N400, N401, N402, N403, N4283 |
| Bipolar Disorder | 29600, 29601, 29602, 29603, 29604, 29605, 29606, 29610, 29611, 29612, 29613, 29614, 29615, 29616, 29640, 29641, 29642, 29643, 29644, 29645, 29646, 29650, 29651, 29652, 29653, 29654, 29655, 29656, 29660, 29661, 29662, 29663, 29664, 29665, 29666, 2967, 29680, 29681, 29682, 29689, 29690, 29699, F3010, F3011, F3012, F3013, F302, F303, F304, F308, F309, F310, F3110, F3111, F3112, F3113, F312, F3130, F3131, F3132, F314, F315, F3160, F3161, F3162, F3163, F3164, F3170, F3171, F3172, F3173, F3174, F3175, F3176, F3177, F3178, F3181, F3189, F319, F338, F3481, F3489, F349, F39 |
| Cataract | 36601, 36602, 36603, 36604, 36609, 36610, 36612, 36613, 36614, 36615, 36616, 36617, 36618, 36619, 36620, 36621, 36622, 36623, 36630, 36645, 36646, 36650, 36651, 36652, 36653, 3668, 3669, 37926, 37931, 37939, 74330, 74331, 74332, 74333, H25011, H25012, H25013, H25019, H25031, H25032, H25033, H25039, H25041, H25042, H25043, H25049, H25091, H25092, H25093, H25099, H2510, H2511, H2512, H2513, H2520, H2521, H2522, H2523, H25811, H25812, H25813, H25819, H2589, H259, H26011, H26012, H26013, H26019, H26031, H26032, H26033, H26039, H26041, H26042, H26043, H26049, H26051, H26052, H26053, H26059, H26061, H26062, H26063, H26069, H2609, H26101, H26102, H26103, H26109, H26111, H26112, H26113, H26119, H26121, H26122, H26123, H26129, H26131, H26132, H26133, H26139, H2620, H26211, H26212, H26213, H26219, H2630, H2631, H2632, H2633, H2640, H26411, H26412, H26413, H26419, H26491, H26492, H26493, H26499, H268, H269, Q120, V431, Z961 |

| | |
|---|---|
| Cerebral Palsy | 33371, 343, 3430, 3431, 3432, 3433, 3434, 3438, 3439, G800, G801, G802, G803, G804, G808, G809 |
| Chronic Kidney Disease | 01600, 01601, 01602, 01603, 01604, 01605, 01606, 0954, 1890, 1899, 2230, 23691, 24940, 24941, 25040, 25041, 25042, 25043, 2714, 27410, 28311, 40301, 40311, 40391, 40402, 40403, 40412, 40413, 40492, 40493, 4401, 4421, 5724, 5800, 5804, 58081, 58089, 5809, 5810, 5811, 5812, 5813, 58181, 58189, 5819, 5820, 5821, 5822, 5824, 58281, 58289, 5829, 5830, 5831, 5832, 5834, 5836, 5837, 58381, 58389, 5839, 5845, 5846, 5847, 5848, 5849, 5851, 5852, 5853, 5854, 5855, 5856, 5859, 586, 587, 5880, 5881, 58881, 58889, 5889, 591, 75312, 75313, 75314, 75315, 75316, 75317, 75319, 75320, 75321, 75322, 75323, 75329, 7944, A1811, A5275, B520, C641, C642, C649, C689, D3000, D3001, D3002, D4100, D4101, D4102, D4110, D4111, D4112, D4120, D4121, D4122, D593, E0821, E0822, E0829, E0865, E0921, E0922, E0929, E1021, E1022, E1029, E1065, E1121, E1122, E1129, E1165, E1321, E1322, E1329, E748, I120, I129, I130, I1310, I1311, I132, I701, I722, K767, M1030, M10311, M10312, M10319, M10321, M10322, M10329, M10331, M10332, M10339, M10341, M10342, M10349, M10351, M10352, M10359, M10361, M10362, M10369, M10371, M10372, M10379, M1038, M1039, M3214, M3215, M3504, N000, N001, N002, N003, N004, N005, N006, N007, N008, N009, N010, N011, N012, N013, N014, N015, N016, N017, N018, N019, N020, N021, N022, N023, N024, N025, N026, N027, N028, N029, N030, N031, N032, N033, N034, N035, N036, N037, N038, N039, N040, N041, N042, N043, N044, N045, N046, N047, N048, N049, N050, N051, N052, N053, N054, N055, N056, N057, N058, N059, N060, N061, N062, N063, N064, N065, N066, N067, N068, N069, N070, N071, N072, N073, N074, N075, N076, N077, N078, N079, N08, N131, N132, N1330, N1339, N140, N141, N142, N143, N144, N150, N158, N159, N16, N170, N171, N172, N178, N179, N181, N182, N183, N184, N185, N186, N189, N19, N250, N251, N2581, N2589, N259, N261, N269, Q6102, Q6111, Q6119, Q612, Q613, Q614, Q615, Q618, Q620, Q6210, Q6211, Q6212, Q622, Q6231, Q6232, Q6239, R944 |
| Chronic Obstructive Pulmonary Disease and Bronchiectasis | 490, 4910, 4911, 49120, 49121, 49122, 4918, 4919, 4920, 4928, 4940, 4941, 496, J40, J410, J411, J418, J42, J430, J431, J432, J438, J439, J440, J441, J449, J470, J471, J479 |
| Colorectal Cancer | 1530, 1531, 1532, 1533, 1534, 1535, 1536, 1537, 1538, 1539, 1540, 1541, 2303, 2304, C180, C181, C182, C183, C184, C185, C186, C187, C188, C189, C19, C20, D010, D011, D012, V1005, V1006, Z85038, Z85040, Z85048 |
| Cystic Fibrosis and Other Metabolic Developmental Disorders | 243, 2552, 2692, 2701, 2702, 2703, 2704, 2706, 2707, 2711, 2770, 27700, 27701, 27702, 27703, 27709, 2776, 27781, 27785, D81810, D841, E000, E001, E002, E009, E030, E031, E250, E258, E259, E569, E700, E701, E7020, E7021, E7029, E7030, E70310, E70311, E70318, E70319, E70320, E70321, E70328, E70329, E70330, E70331, E70338, E70339, E7039, E705, E708, E709, E710, E71110, E71111, E71118, E7119, E712, E71310, E71311, E71312, E71313, E71314, E71318, E7132, E7141, E7210, E7211, E7212, E7219, E7220, E7221, E7222, E7223, E7229, E723, E724, E7250, E7251, E7259, E728, E7420, E7421, E7429, E840, E8411, E8419, E848, E849 |
| Depression | 29620, 29621, 29622, 29623, 29624, 29625, 29626, 29630, 29631, 29632, 29633, 29634, 29635, 29636, 29651, 29652, 29653, 29654, 29655, 29656, 29660, 29661, 29662, 29663, 29664, 29665, 29666, 29689, 2980, 3004, 3091, 311, F3130, F3131, F3132, F314, F315, F3160, F3161, F3162, F3163, F3164, F3175, F3176, F3177, F3178, F3181, F320, F321, F322, F323, F324, F325, F329, F330, F331, F332, F333, F3340, F3341, F3342, F338, F339, F341, F4321, F4323 |

| | |
|---|---|
| Depressive Disorders | 29620, 29621, 29622, 29623, 29624, 29625, 29626, 29630, 29631, 29632, 29633, 29634, 29635, 29636, 3004, 311, F320, F321, F322, F323, F324, F325, F3289, F329, F330, F331, F332, F333, F3340, F3341, F3342, F338, F339, F341, V790 |

| | |
|---|---|
| Diabetes | 24900, 24901, 24910, 24911, 24920, 24921, 24930, 24931, 24940, 24941, 24950, 24951, 24960, 24961, 24970, 24971, 24980, 24981, 24990, 24991, 25000, 25001, 25002, 25003, 25010, 25011, 25012, 25013, 25020, 25021, 25022, 25023, 25030, 25031, 25032, 25033, 25040, 25041, 25042, 25043, 25050, 25051, 25052, 25053, 25060, 25061, 25062, 25063, 25070, 25071, 25072, 25073, 25080, 25081, 25082, 25083, 25090, 25091, 25092, 25093, 3572, 36201, 36202, 36203, 36204, 36205, 36206, 36641, E0800, E0801, E0810, E0811, E0821, E0822, E0829, E08311, E08319, E08321, E083211, E083212, E083213, E083219, E08329, E083291, E083292, E083293, E083299, E08331, E083311, E083312, E083313, E083319, E08339, E083391, E083392, E083393, E083399, E08341, E083411, E083412, E083413, E083419, E08349, E083491, E083492, E083493, E083499, E08351, E083511, E083512, E083513, E083519, E083521, E083522, E083523, E083529, E083531, E083532, E083533, E083539, E083541, E083542, E083543, E083549, E083551, E083552, E083553, E083559, E08359, E083591, E083592, E083593, E083599, E0836, E0837X1, E0837X2, E0837X3, E0837X9, E0839, E0840, E0841, E0842, E0843, E0844, E0849, E0851, E0852, E0859, E08610, E08618, E08620, E08621, E08622, E08628, E08630, E08638, E08641, E08649, E0865, E0869, E088, E089, E0900, E0901, E0910, E0911, E0921, E0922, E0929, E09311, E09319, E09321, E093211, E093212, E093213, E093219, E09329, E093291, E093292, E093293, E093299, E09331, E093311, E093312, E093313, E093319, E09339, E093391, E093392, E093393, E093399, E09341, E093411, E093412, E093413, E093419, E09349, E093491, E093492, E093493, E093499, E09351, E093511, E093512, E093513, E093519, E093521, E093522, E093523, E093529, E093531, E093532, E093533, E093539, E093541, E093542, E093543, E093549, E093551, E093552, E093553, E093559, E09359, E093591, E093592, E093593, E093599, E0936, E0937X1, E0937X2, E0937X3, E0937X9, E0939, E0940, E0941, E0942, E0943, E0944, E0949, E0951, E0952, E0959, E09610, E09618, E09620, E09621, E09622, E09628, E09630, E09638, E09641, E09649, E0965, E0969, E098, E099, E1010, E1011, E1021, E1022, E1029, E10311, E10319, E10321, E103211, E103212, E103213, E103219, E10329, E103291, E103292, E103293, E103299, E10331, E103311, E103312, E103313, E103319, E10339, E103391, E103392, E103393, E103399, E10341, E103411, E103412, E103413, E103419, E10349, E103491, E103492, E103493, E103499, E10351, E103511, E103512, E103513, E103519, E10359, E1036, E1037X1, E1037X2, E1037X3, E1037X9, E1039, E1040, E1041, E1042, E1043, E1044, E1049, E1051, E1052, E1059, E10610, E10618, E10620, E10621, E10622, E10628, E10630, E10638, E10641, E10649, E1065, E1069, E108, E109, E1100, E1101, E1110, E1111, E1121, E1122, E1129, E11311, E11319, E11321, E113211, E113212, E113213, E113219, E11329, E113291, E113292, E113293, E113299, E11331, E113311, E113312, E113313, E113319, E11339, E113391, E113392, E113393, E113399, E11341, E113411, E113412, E113413, E113419, E11349, E113491, E113492, E113493, E113499, E11351, E113511, E113512, E113513, E113519, E113521, E113522, E113523, E113529, E113531, E113532, E113533, E113539, E113541, E113542, E113543, E113549, E113551, E113552, E113553, E113559, E11359, E113591, E113592, E113593, E113599, E1136, E1137X1, E1137X2, E1137X3, E1137X9, E1139, E1140, E1141, E1142, E1143, E1144, E1149, E1151, E1152, E1159, E11610, E11618, E11620, E11621, E11622, E11628, E11630, E11638, E11641, E11649, E1165, E1169, E118, E119, E1300, E1301, E1310, E1311, E1321, E1322, E1329, E13311, E13319, E13321, E133211, E133212, E133213, E133219, E13329, E133291, E133292, E133293, E133299, E13331, E133311, E133312, E133313, E133319, E13339, E133391, E133392, E133393, E133399, E13341, E133411, E133412, E133413, E133419, E13349, E133491, E133492, E133493, E133499, E13351, E133511, E133512, E133513, E133519, E133521, E133522, E133523, E133529, E133531, E133532, E133533, E133539, E133541, E133542, E133543, E133549, E133551, E133552, E133553, E133559, E13359, E1336, E1339, E1340, E1341, E1342, E1343, E1344, E1349, E1351, E1352, E1359, E13610, E13618, E13620, E13621, E13622, E13628, E13630, E13638, E13641, E13649, E1365, E1369, E138, E139 |

| | |
|---|---|
| Drug Use Disorders | 2920, 29211, 29212, 2922, 29281, 29282, 29283, 29284, 29285, 29289, 2929, 30400, 30401, 30402, 30410, 30411, 30412, 3042, 30420, 30421, 30422, 3043, 30430, 30431, 30432, 3044, 30440, 30441, 30442, 3045, 30450, 30451, 30452, 3046, 30460, 30461, 30462, 3047, 30470, 30471, 30472, 3048, 30480, 30481, 30482, 3049, 30490, 30491, 30492, 3052, 30520, 30521, 30522, 3053, 30530, 30531, 30532, 3054, 30540, 30541, 30542, 3055, 30550, 30551, 30552, 3056, 30560, 30561, 30562, 3057, 30570, 30571, 30572, 3058, 30580, 30581, 30582, 3059, 30590, 30591, 30592, 6483, 64830, 64831, 64832, 64833, 64834, 6555, 65550, 65551, 65553, 76072, 76073, 76075, 7795, 9650, 96500, 96501, 96502, 96509, E8500, E8501, E8502, E8541, E9350, E9351, F1110, F11120, F11121, F11122, F11129, F1114, F11150, F11151, F11159, F11181, F11182, F11188, F1119, F1120, F11220, F11221, F11222, F11229, F1123, F1124, F11250, F11251, F11259, F11281, F11282, F11288, F1129, F1190, F11920, F11921, F11922, F11929, F1193, F1194, F11950, F11951, F11959, F11981, F11982, F11988, F1199, F1210, F12120, F12121, F12122, F12129, F12150, F12151, F12159, F12180, F12188, F1219, F1220, F12220, F12221, F12222, F12229, F12250, F12251, F12259, F12280, F12288, F1229, F1290, F12920, F12921, F12922, F12929, F12950, F12951, F12959, F12980, F12988, F1299, F1310, F13120, F13121, F13129, F1314, F13150, F13151, F13159, F13180, F13181, F13182, F13188, F1319, F1320, F13220, F13221, F13229, F13230, F13231, F13232, F13239, F1324, F13250, F13251, F13259, F1326, F1327, F13280, F13281, F13282, F13288, F1329, F1390, F13920, F13921, F13929, F13930, F13931, F13932, F13939, F1394, F13950, F13951, F13959, F1396, F1397, F13980, F13981, F13982, F13988, F1399, F1410, F14120, F14121, F14122, F14129, F1414, F14150, F14151, F14159, F14180, F14181, F14182, F14188, F1419, F1420, F14220, F14221, F14222, F14229, F1423, F1424, F14250, F14251, F14259, F14280, F14281, F14282, F14288, F1429, F1490, F14920, F14921, F14922, F14929, F1494, F14950, F14951, F14959, F14980, F14981, F14982, F14988, F1499, F1510, F15120, F15121, F15122, F15129, F1514, F15150, F15151, F15159, F15180, F15181, F15182, F15188, F1519, F1520, F15220, F15221, F15222, F15229, F1523, F1524, F15250, F15251, F15259, F15280, F15281, F15282, F15288, F1529, F1590, F15920, F15921, F15922, F15929, F1593, F1594, F15950, F15951, F15959, F15980, F15981, F15982, F15988, F1599, F1610, F16120, F16121, F16122, F16129, F1614, F16150, F16151, F16159, F16180, F16183, F16188, F1619, F1620, F16220, F16221, F16229, F1624, F16250, F16251, F16259, F16280, F16283, F16288, F1629, F1690, F16920, F16921, F16929, F1694, F16950, F16951, F16959, F16980, F16983, F16988, F1699, F17203, F17208, F17209, F17213, F17218, F17219, F17223, F17228, F17229, F17293, F17298, F17299, F1810, F18120, F18121, F18129, F1814, F18150, F18151, F18159, F1817, F18180, F18188, F1819, F1820, F18220, F18221, F18229, F1824, F18250, F18251, F18259, F1827, F18280, F18288, F1829, F1890, F18920, F18921, F18929, F1894, F18950, F18951, F18959, F1897, F18980, F18988, F1899, F1910, F19120, F19121, F19122, F19129, F1914, F19150, F19151, F19159, F1916, F1917, F19180, F19181, F19182, F19188, F1919, F1920, F19220, F19221, F19222, F19229, F19230, F19231, F19232, F19239, F1924, F19250, F19251, F19259, F1926, F1927, F19280, F19281, F19282, F19288, F1929, F1990, F19920, F19921, F19922, F19929, F19930, F19931, F19932, F19939, F1994, F19950, F19951, F19959, F1996, F1997, F19980, F19981, F19982, F19988, F1999, F550, F551, F552, F553, F554, F558, O355XX0, O355XX1, O355XX2, O355XX3, O355XX4, O355XX5, O355XX9, O99320, O99321, O99322, O99323, O99324, O99325, P0441, P0449, P961, P962, T400X1A, T400X2A, T400X3A, T400X4A, T400X5A, T400X5S, T401X1A, T401X2A, T401X3A, T401X4A, T402X1A, T402X2A, T402X3A, T402X4A, T403X1A, T403X2A, T403X3A, T403X4A, T403X5A, T403X5S, T404X1A, T404X2A, T404X3A, T404X4A, T40601A, T40602A, T40603A, T40604A, T40691A, T40692A, T40693A, T40694A, T407X1A, T408X1A, T40901A, T40991A, V6542, Z7141, Z7142, Z7151, Z7152, Z716 |
| Endometrial Cancer | 1820, 2332, C541, C542, C543, C548, C549, D070, V1042, Z8542 |

| | |
|---|---|
| Epilepsy | 345, 3450, 34500, 34501, 3451, 34510, 34511, 3452, 3453, 3454, 34540, 34541, 3455, 34550, 34551, 3456, 34560, 34561, 3457, 34570, 34571, 3458, 34580, 34581, 3459, 34590, 34591, G40001, G40009, G40011, G40019, G40101, G40109, G40111, G40119, G40201, G40209, G40211, G40219, G40301, G40309, G40311, G40319, G40401, G40409, G40411, G40419, G40501, G40509, G40801, G40802, G40803, G40804, G40811, G40812, G40813, G40814, G40821, G40822, G40823, G40824, G4089, G40901, G40909, G40911, G40919, G40A01, G40A09, G40A11, G40A19, G40B01, G40B09, G40B11, G40B19 |
| Female / Male Breast Cancer | 1740, 1741, 1742, 1743, 1744, 1745, 1746, 1748, 1749, 1750, 1759, 2330, C50011, C50012, C50019, C50021, C50022, C50029, C50111, C50112, C50119, C50121, C50122, C50129, C50211, C50212, C50219, C50221, C50222, C50229, C50311, C50312, C50319, C50321, C50322, C50329, C50411, C50412, C50419, C50421, C50422, C50429, C50511, C50512, C50519, C50521, C50522, C50529, C50611, C50612, C50619, C50621, C50622, C50629, C50811, C50812, C50819, C50821, C50822, C50829, C50911, C50912, C50919, C50921, C50922, C50929, D0500, D0501, D0502, D0510, D0511, D0512, D0580, D0581, D0582, D0590, D0591, D0592, V103, Z853 |
| Fibromyalgia, Chronic Pain and Fatigue | 3382, 33821, 33822, 33823, 33829, 3383, 3384, 7291, 7292, 7807, 78071, G8921, G8922, G8928, G8929, G893, G894, M5410, M5411, M5412, M5413, M5414, M5415, M5416, M5417, M5418, M6080, M60811, M60812, M60819, M60821, M60822, M60829, M60831, M60832, M60839, M60841, M60842, M60849, M60851, M60852, M60859, M60861, M60862, M60869, M60871, M60872, M60879, M6088, M6089, M609, M791, M7910, M7911, M7912, M7918, M792, M797, R5382 |

| | |
|---|---|
| Glaucoma | 36285, 36500, 36501, 36502, 36503, 36504, 36510, 36511, 36512, 36513, 36515, 36520, 36521, 36522, 36523, 36524, 36531, 36532, 36541, 36542, 36543, 36551, 36552, 36559, 36560, 36561, 36562, 36563, 36564, 36565, 36581, 36582, 36583, 36589, 3659, 37714, H40001, H40002, H40003, H40009, H40011, H40012, H40013, H40019, H40031, H40032, H40033, H40039, H40041, H40042, H40043, H40049, H40051, H40052, H40053, H40059, H4010X0, H4010X1, H4010X2, H4010X3, H4010X4, H401110, H401111, H401112, H401113, H401114, H401120, H401121, H401122, H401123, H401124, H401130, H401131, H401132, H401133, H401134, H401190, H401191, H401192, H401193, H401194, H4011X0, H4011X1, H4011X2, H4011X3, H4011X4, H401210, H401211, H401212, H401213, H401214, H401220, H401221, H401222, H401223, H401224, H401230, H401231, H401232, H401233, H401234, H401290, H401291, H401292, H401293, H401294, H401310, H401311, H401312, H401313, H401314, H401320, H401321, H401322, H401323, H401324, H401330, H401331, H401332, H401333, H401334, H401390, H401391, H401392, H401393, H401394, H401410, H401411, H401412, H401413, H401414, H401420, H401421, H401422, H401423, H401424, H401430, H401431, H401432, H401433, H401434, H401490, H401491, H401492, H401493, H401494, H40151, H40152, H40153, H40159, H4020X0, H4020X1, H4020X2, H4020X3, H4020X4, H40211, H40212, H40213, H40219, H402210, H402211, H402212, H402213, H402214, H402220, H402221, H402222, H402223, H402224, H402230, H402231, H402232, H402233, H402234, H402290, H402291, H402292, H402293, H402294, H40231, H40232, H40233, H40239, H40241, H40242, H40243, H40249, H4030X0, H4030X1, H4030X2, H4030X3, H4030X4, H4031X0, H4031X1, H4031X2, H4031X3, H4031X4, H4032X0, H4032X1, H4032X2, H4032X3, H4032X4, H4033X0, H4033X1, H4033X2, H4033X3, H4033X4, H4040X0, H4040X1, H4040X2, H4040X3, H4040X4, H4041X0, H4041X1, H4041X2, H4041X3, H4041X4, H4042X0, H4042X1, H4042X2, H4042X3, H4042X4, H4043X0, H4043X1, H4043X2, H4043X3, H4043X4, H4050X0, H4050X1, H4050X2, H4050X3, H4050X4, H4051X0, H4051X1, H4051X2, H4051X3, H4051X4, H4052X0, H4052X1, H4052X2, H4052X3, H4052X4, H4053X0, H4053X1, H4053X2, H4053X3, H4053X4, H4060X0, H4060X1, H4060X2, H4060X3, H4060X4, H4061X0, H4061X1, H4061X2, H4061X3, H4061X4, H4062X0, H4062X1, H4062X2, H4062X3, H4062X4, H4063X0, H4063X1, H4063X2, H4063X3, H4063X4, H40811, H40812, H40813, H40819, H40821, H40822, H40823, H40829, H40831, H40832, H40833, H40839, H4089, H409, H42, H44511, H44512, H44513, H44519, H47231, H47232, H47233, H47239,<br>Q150<br>, |
| Heart Failure | 39891, 40201, 40211, 40291, 40401, 40403, 40411, 40413, 40491, 40493, 4280, 4281, 42820, 42821, 42822, 42823, 42830, 42831, 42832, 42833, 42840, 42841, 42842, 42843, 4289, I0981, I110, I130, I132, I501, I5020, I5021, I5022, I5023, I5030, I5031, I5032, I5033, I5040, I5041, I5042, I5043, I50810, I50811, I50812, I50813, I50814, I5082, I5083, I5084, I5089, I509 |

| | |
|---|---|
| Hip/Pelvic Fracture | 73314, 73315, 73396, 73397, 73398, 8080, 8081, 8082, 8083, 80841, 80842, 80843, 80844, 80849, 80851, 80852, 80853, 80854, 80859, 8088, 8089, 82000, 82001, 82002, 82003, 82009, 82010, 82011, 82012, 82013, 82019, 82020, 82021, 82022, 82030, 82031, 82032, 8208, 8209, M80051A, M80052A, M80059A, M80851A, M80852A, M80859A, M84350A, M84351A, M84352A, M84353A, M84359A, M84451A, M84452A, M84453A, M84459A, M84550A, M84551A, M84552A, M84553A, M84559A, M84650A, M84651A, M84652A, M84653A, M84659A, S32301A, S32301B, S32302A, S32302B, S32309A, S32309B, S32311A, S32311B, S32312A, S32312B, S32313A, S32313B, S32314A, S32314B, S32315A, S32315B, S32316A, S32316B, S32391A, S32391B, S32392A, S32392B, S32399A, S32399B, S32401A, S32401B, S32402A, S32402B, S32409A, S32409B, S32411A, S32411B, S32412A, S32412B, S32413A, S32413B, S32414A, S32414B, S32415A, S32415B, S32416A, S32416B, S32421A, S32421B, S32422A, S32422B, S32423A, S32423B, S32424A, S32424B, S32425A, S32425B, S32426A, S32426B, S32431A, S32431B, S32432A, S32432B, S32433A, S32433B, S32434A, S32434B, S32435A, S32435B, S32436A, S32436B, S32441A, S32441B, S32442A, S32442B, S32443A, S32443B, S32444A, S32444B, S32445A, S32445B, S32446A, S32446B, S32451A, S32451B, S32452A, S32452B, S32453A, S32453B, S32454A, S32454B, S32455A, S32455B, S32456A, S32456B, S32461A, S32461B, S32462A, S32462B, S32463A, S32463B, S32464A, S32464B, S32465A, S32465B, S32466A, S32466B, S32471A, S32471B, S32472A, S32472B, S32473A, S32473B, S32474A, S32474B, S32475A, S32475B, S32476A, S32476B, S32481A, S32481B, S32482A, S32482B, S32483A, S32483B, S32484A, S32484B, S32485A, S32485B, S32486A, S32486B, S32491A, S32491B, S32492A, S32492B, S32499A, S32499B, S32501A, S32501B, S32502A, S32502B, S32509A, S32509B, S32511A, S32511B, S32512A, S32512B, S32519A, S32519B, S32591A, S32591B, S32592A, S32592B, S32599A, S32599B, S32601A, S32601B, S32602A, S32602B, S32609A, S32609B, S32611A, S32611B, S32612A, S32612B, S32613A, S32613B, S32614A, S32614B, S32615A, S32615B, S32616A, S32616B, S32691A, S32691B, S32692A, S32692B, S32699A, S32699B, S32810A, S32810B, S32811A, S32811B, S3282XA, S3282XB, S3289XA, S3289XB, S329XXA, S329XXB, S72001A, S72001B, S72001C, S72002A, S72002B, S72002C, S72009A, S72009B, S72009C, S72011A, S72011B, S72011C, S72012A, S72012B, S72012C, S72019A, S72019B, S72019C, S72021A, S72021B, S72021C, S72022A, S72022B, S72022C, S72023A, S72023B, S72023C, S72024A, S72024B, S72024C, S72025A, S72025B, S72025C, S72026A, S72026B, S72026C, S72031A, S72031B, S72031C, S72032A, S72032B, S72032C, S72033A, S72033B, S72033C, S72034A, S72034B, S72034C, S72035A, S72035B, S72035C, S72036A, S72036B, S72036C, S72041A, S72041B, S72041C, S72042A, S72042B, S72042C, S72043A, S72043B, S72043C, S72044A, S72044B, S72044C, S72045A, S72045B, S72045C, S72046A, S72046B, S72046C, S72051A, S72051B, S72051C, S72052A, S72052B, S72052C, S72059A, S72059B, S72059C, S72061A, S72061B, S72061C, S72062A, S72062B, S72062C, S72063A, S72063B, S72063C, S72064A, S72064B, S72064C, S72065A, S72065B, S72065C, S72066A, S72066B, S72066C, S72091A, S72091B, S72091C, S72092A, S72092B, S72092C, S72099A, S72099B, S72099C, S72101A, S72101B, S72101C, S72102A, S72102B, S72102C, S72109A, S72109B, S72109C, S72111A, S72111B, S72111C, S72112A, S72112B, S72112C, S72113A, S72113B, S72113C, S72114A, S72114B, S72114C, S72115A, S72115B, S72115C, S72116A, S72116B, S72116C, S72121A, S72121B, S72121C, S72122A, S72122B, S72122C, S72123A, S72123B, S72123C, S72124A, S72124B, S72124C, S72125A, S72125B, S72125C, S72126A, S72126B, S72126C, S72131A, S72131B, S72131C, S72132A, S72132B, S72132C, S72133A, S72133B, S72133C, S72134A, S72134B, S72134C, S72135A, S72135B, S72135C, S72136A, S72136B, S72136C, S72141A, S72141B, S72141C, S72142A, S72142B, S72142C, S72143A, S72143B, S72143C, S72144A, S72144B, S72144C, S72145A, S72145B, S72145C, S72146A, S72146B, S72146C, S7221XA, S7221XB, S7221XC, S7222XA, S7222XB, S7222XC, S7223XA, S7223XB, S7223XC, S7224XA, S7224XB, S7224XC, S7225XA, S7225XB, S7225XC, S7226XA, S7226XB, S7226XC, S79001A, S79002A, S79009A, S79011A, S79012A, S79019A, S79091A, S79092A, S79099A |
| Human Immunodeficiency Virus and/or Acquired Immunodeficiency Syndrome | 042, 0420, 0421, 0422, 0429, 043, 0431, 0432, 0433, 0439, 044, 0440, 0449, 07953, 79571, B20, B9735, R75, V08, Z21 |

| | |
|---|---|
| (HIV/AIDS) | |
| Hyperlipidemia | 2720, 2721, 2722, 2723, 2724, E780, E7800, E7801, E781, E782, E783, E784, E7841, E7849, E785 |
| Hypertension | 36211, 4010, 4011, 4019, 40200, 40201, 40210, 40211, 40290, 40291, 40300, 40301, 40310, 40311, 40390, 40391, 40400, 40401, 40402, 40403, 40410, 40411, 40412, 40413, 40490, 40491, 40492, 40493, 40501, 40509, 40511, 40519, 40591, 40599, 4372, H35031, H35032, H35033, H35039, I10, I110, I119, I120, I129, I130, I1310, I1311, I132, I150, I151, I152, I158, I159, I674, N262 |
| Intellectual Disabilities and Related Conditions | 317, 318, 3180, 3181, 3182, 319, 758, 7580, 7581, 7582, 7583, 75831, 75832, 75833, 75839, 7585, 7597, 75981, 75983, 75989, 76071, E7871, E7872, F70, F71, F72, F73, F78, F79, P043, Q860, Q871, Q872, Q873, Q875, Q8781, Q8789, Q897, Q898, Q900, Q901, Q902, Q909, Q910, Q911, Q912, Q913, Q914, Q915, Q916, Q917, Q920, Q921, Q922, Q925, Q9261, Q9262, Q927, Q928, Q929, Q930, Q931, Q932, Q933, Q934, Q935, Q9351, Q93529, Q937, Q9381, Q9388, Q9389, Q939, Q952, Q953, Q992 |
| Ischemic Heart Disease | 41000, 41001, 41002, 41010, 41011, 41012, 41020, 41021, 41022, 41030, 41031, 41032, 41040, 41041, 41042, 41050, 41051, 41052, 41060, 41061, 41062, 41070, 41071, 41072, 41080, 41081, 41082, 41090, 41091, 41092, 4110, 4111, 41181, 41189, 412, 4130, 4131, 4139, 41400, 41401, 41402, 41403, 41404, 41405, 41406, 41407, 41412, 4142, 4143, 4144, 4148, 4149, I200, I201, I208, I209, I2101, I2102, I2109, I2111, I2119, I2121, I2129, I213, I214, I21A1, I21A9, I220, I221, I222, I228, I229, I230, I231, I232, I233, I234, I235, I236, I237, I238, I240, I241, I248, I249, I2510, I25110, I25111, I25118, I25119, I252, I253, I2541, I2542, I255, I256, I25700, I25701, I25708, I25709, I25710, I25711, I25718, I25719, I25720, I25721, I25728, I25729, I25730, I25731, I25738, I25739, I25750, I25751, I25758, I25759, I25760, I25761, I25768, I25769, I25790, I25791, I25798, I25799, I25810, I25811, I25812, I2582, I2583, I2584, I2589, I259 |
| Learning Disabilities | 315, 31501, 31502, 31509, 3151, 3152, 31531, 31532, 31534, 31535, 31539, 3154, F800, F801, F802, F804, F8081, F8082, F8089, F809, F810, F812, F8181, F8189, F819, F82, H9325, R480 |
| Leukemias and Lymphomas | 2000, 20000, 20001, 20002, 20003, 20004, 20005, 20006, 20007, 20008, 2001, 20010, 20011, 20012, 20013, 20014, 20015, 20016, 20017, 20018, 2002, 20020, 20021, 20022, 20023, 20024, 20025, 20026, 20027, 20028, 2003, 20030, 20031, 20032, 20033, 20034, 20035, 20036, 20037, 20038, 2004, 20040, 20041, 20042, 20043, 20044, 20045, 20046, 20047, 20048, 2005, 20050, 20051, 20052, 20053, 20054, 20055, 20056, 20057, 20058, 2006, 20060, 20061, 20062, 20063, 20064, 20065, 20066, 20067, 20068, 2007, 20070, 20071, 20072, 20073, 20074, 20075, 20076, 20077, 20078, 2008, 20080, 20081, 20082, 20083, 20084, 20085, 20086, 20087, 20088, 2010, 20100, 20101, 20102, 20103, 20104, 20105, 20106, 20107, 20108, 2011, 20110, 20111, 20112, 20113, 20114, 20115, 20116, 20117, 20118, 2012, 20120, 20121, 20122, 20123, 20124, 20125, 20126, 20127, 20128, 2014, 20140, 20141, 20142, 20143, 20144, 20145, 20146, 20147, 20148, 2015, 20150, 20151, 20152, 20153, 20154, 20155, 20156, 20157, 20158, 2016, 20160, 20161, 20162, 20163, 20164, 20165, 20166, 20167, 20168, 2017, 20170, 20171, 20172, 20173, 20174, 20175, 20176, 20177, 20178, 2019, 20190, 20191, 20192, 20193, 20194, 20195, 20196, 20197, 20198, 2020, 20200, 20201, 20202, 20203, 20204, 20205, 20206, 20207, 20208, 2021, 20210, 20211, 20212, 20213, 20214, 20215, 20216, 20217, 20218, 2022, 20220, 20221, 20222, 20223, 20224, 20225, 20226, 20227, 20228, 2024, 20240, 20241, 20242, 20243, 20244, 20245, 20246, 20247, 20248, 2027, 20270, 20271, 20272, 20273, 20274, 20275, 20276, 20277, 20278, 2028, 20280, 20281, 20282, 20283, 20284, 20285, 20286, 20287, 20288, 2029, 20290, 20291, 20292, 20293, 20294, 20295, 20296, 20297, 20298, 2031, 20310, 20311, 20312, 2040, 20400, |

| | |
|---|---|
| | 20401, 20402, 2041, 20410, 20411, 20412, 2042, 20420, 20421, 20422, 2048, 20480, 20481, 20482, 2049, 20490, 20491, 20492, 2050, 20500, 20501, 20502, 2051, 20510, 20511, 20512, 2052, 20520, 20521, 20522, 2053, 20530, 20531, 20532, 2058, 20580, 20581, 20582, 2059, 20590, 20591, 20592, 2060, 20600, 20601, 20602, 2061, 20610, 20611, 20612, 2062, 20620, 20621, 20622, 2068, 20680, 20681, 20682, 2069, 20690, 20691, 20692, 2070, 20700, 20701, 20702, 2071, 20710, 20711, 20712, 2072, 20720, 20721, 20722, 2078, 20780, 20781, 20782, 2080, 20800, 20801, 20802, 2081, 20810, 20811, 20812, 2082, 20820, 20821, 20822, 2088, 20880, 20881, 20882, 2089, 20890, 20891, 20892, C8100, C8101, C8102, C8103, C8104, C8105, C8106, C8107, C8108, C8109, C8110, C8111, C8112, C8113, C8114, C8115, C8116, C8117, C8118, C8119, C8120, C8121, C8122, C8123, C8124, C8125, C8126, C8127, C8128, C8129, C8130, C8131, C8132, C8133, C8134, C8135, C8136, C8137, C8138, C8139, C8140, C8141, C8142, C8143, C8144, C8145, C8146, C8147, C8148, C8149, C8170, C8171, C8172, C8173, C8174, C8175, C8176, C8177, C8178, C8179, C8190, C8191, C8192, C8193, C8194, C8195, C8196, C8197, C8198, C8199, C8200, C8201, C8202, C8203, C8204, C8205, C8206, C8207, C8208, C8209, C8210, C8211, C8212, C8213, C8214, C8215, C8216, C8217, C8218, C8219, C8220, C8221, C8222, C8223, C8224, C8225, C8226, C8227, C8228, C8229, C8230, C8231, C8232, C8233, C8234, C8235, C8236, C8237, C8238, C8239, C8240, C8241, C8242, C8243, C8244, C8245, C8246, C8247, C8248, C8249, C8250, C8251, C8252, C8253, C8254, C8255, C8256, C8257, C8258, C8259, C8260, C8261, C8262, C8263, C8264, C8265, C8266, C8267, C8268, C8269, C8280, C8281, C8282, C8283, C8284, C8285, C8286, C8287, C8288, C8289, C8290, C8291, C8292, C8293, C8294, C8295, C8296, C8297, C8298, C8299, C8300, C8301, C8302, C8303, C8304, C8305, C8306, C8307, C8308, C8309, C8310, C8311, C8312, C8313, C8314, C8315, C8316, C8317, C8318, C8319, C8330, C8331, C8332, C8333, C8334, C8335, C8336, C8337, C8338, C8339, C8350, C8351, C8352, C8353, C8354, C8355, C8356, C8357, C8358, C8359, C8370, C8371, C8372, C8373, C8374, C8375, C8376, C8377, C8378, C8379, C8380, C8381, C8382, C8383, C8384, C8385, C8386, C8387, C8388, C8389, C8390, C8391, C8392, C8393, C8394, C8395, C8396, C8397, C8398, C8399, C8400, C8401, C8402, C8403, C8404, C8405, C8406, C8407, C8408, C8409, C8410, C8411, C8412, C8413, C8414, C8415, C8416, C8417, C8418, C8419, C8440, C8441, C8442, C8443, C8444, C8445, C8446, C8447, C8448, C8449, C8460, C8461, C8462, C8463, C8464, C8465, C8466, C8467, C8468, C8469, C8470, C8471, C8472, C8473, C8474, C8475, C8476, C8477, C8478, C8479, C8490, C8491, C8492, C8493, C8494, C8495, C8496, C8497, C8498, C8499, C84A0, C84A1, C84A2, C84A3, C84A4, C84A5, C84A6, C84A7, C84A8, C84A9, C84Z0, C84Z1, C84Z2, C84Z3, C84Z4, C84Z5, C84Z6, C84Z7, C84Z8, C84Z9, C8510, C8511, C8512, C8513, C8514, C8515, C8516, C8517, C8518, C8519, C8520, C8521, C8522, C8523, C8524, C8525, C8526, C8527, C8528, C8529, C8580, C8581, C8582, C8583, C8584, C8585, C8586, C8587, C8588, C8589, C8590, C8591, C8592, C8593, C8594, C8595, C8596, C8597, C8598, C8599, C860, C861, C862, C863, C864, C865, C866, C884, C9010, C9011, C9012, C9100, C9101, C9102, C9110, C9111, C9112, C9130, C9131, C9132, C9140, C9141, C9142, C9150, C9151, C9152, C9160, C9161, C9162, C9190, C9191, C9192, C91A0, C91A1, C91A2, C91Z0, C91Z1, C91Z2, C9200, C9201, C9202, C9210, C9211, C9212, C9220, C9221, C9222, C9230, C9231, C9232, C9240, C9241, C9242, C9250, C9251, C9252, C9260, C9261, C9262, C9290, C9291, C9292, C92A0, C92A1, C92A2, C92Z0, C92Z1, C92Z2, C9300, C9301, C9302, C9310, C9311, C9312, C9330, C9331, C9332, C9390, C9391, C9392, C93Z0, C93Z1, C93Z2, C9400, C9401, C9402, C9420, C9421, C9422, C9430, C9431, C9432, C9480, C9481, C9482, C9500, C9501, C9502, C9510, C9511, C9512, C9590, C9591, C9592, C964, C969, C96Z, D45, V 1063, V106, V1060, V1061, V1062, V1069, V107, V1071, V1072, V1079, Z85231, Z856, Z8571, Z8579 |
| Liver Disease, Cirrhosis and Other Liver Conditions (except Viral Hepatitis) | 570, 571, 5710, 5711, 5712, 5713, 5715, 5716, 5718, 5719, 572, 5720, 5721, 5722, 5723, 5724, 5728, 573, 5730, 5734, 5735, 5738, 5739, 5761, 7891, K700, K7010, K7011, K702, K7030, K7031, K7040, K7041, K709, K710, K7111, K717, K718, K719, K7200, K7201, K7210, K7211, K7290, K7291, K740, K741, K742, K743, K744, K745, K7460, K7469, K750, K751, K7581, K7589, K759, K760, K761, K762, K763, K765, K766, K767, K7681, K7689, K769, K77, K8030, K8031, K8032, K8033, K8034, K8035, K8036, K8037, K830, R160, R162, V427, Z4823, Z944 |

| | |
|---|---|
| Lung Cancer | 1622, 1623, 1624, 1625, 1628, 1629, 2312, C3400, C3401, C3402, C3410, C3411, C3412, C342, C3430, C3431, C3432, C3480, C3481, C3482, C3490, C3491, C3492, D0220, D0221, D0222, V1011, Z85110, Z85118 |
| Migraine and Chronic Headache | 339, 3390, 33900, 33901, 33902, 33903, 33904, 33905, 33909, 3391, 33910, 33911, 33912, 3392, 33920, 33921, 33922, 3393, 3394, 33941, 33942, 33943, 33944, 3398, 33981, 33982, 33983, 33984, 33985, 33989, 346, 3460, 34600, 34601, 34602, 34603, 3461, 34610, 34611, 34612, 34613, 3462, 34620, 34621, 34622, 34623, 3463, 34630, 34631, 34632, 34633, 3464, 34640, 34641, 34642, 34643, 3465, 34650, 34651, 34652, 34653, 3466, 34660, 34661, 34662, 34663, 3467, 34670, 34671, 34672, 34673, 3468, 34680, 34681, 34682, 34683, 3469, 34690, 34691, 34692, 34693, G43001, G43009, G43011, G43019, G43101, G43109, G43111, G43119, G43401, G43409, G43411, G43419, G43501, G43509, G43511, G43519, G43601, G43609, G43611, G43619, G43701, G43709, G43711, G43719, G43801, G43809, G43811, G43819, G43821, G43829, G43831, G43839, G43901, G43909, G43911, G43919, G43A0, G43A1, G43B0, G43B1, G43C0, G43C1, G43D0, G43D1, G44001, G44009, G44011, G44019, G44021, G44029, G44031, G44039, G44041, G44049, G44051, G44059, G44091, G44099, G441, G44201, G44209, G44211, G44219, G44221, G44229, G44301, G44309, G44311, G44319, G44321, G44329, G4440, G4441, G4451, G4452, G4453, G4459, G4481, G4482, G4483, G4484, G4485, G4489 |
| Mobility Impairments | 3341, 34200, 34201, 34202, 34210, 34211, 34212, 34280, 34281, 34282, 34290, 34291, 34292, 344, 3440, 34400, 34401, 34402, 34403, 34404, 34409, 3441, 3442, 3443, 34430, 34431, 34432, 3444, 34440, 34441, 34442, 3445, 3446, 34460, 34461, 3448, 34481, 34489, 3449, 43820, 43821, 43822, 43830, 43831, 43832, 43840, 43841, 43842, 43850, 43851, 43852, 43853, G041, G114, G8100, G8101, G8102, G8103, G8104, G8110, G8111, G8112, G8113, G8114, G8190, G8191, G8192, G8193, G8194, G8220, G8221, G8222, G8250, G8251, G8252, G8253, G8254, G830, G8310, G8311, G8312, G8313, G8314, G8320, G8321, G8322, G8323, G8324, G8330, G8331, G8332, G8333, G8334, G834, G835, G8381, G8382, G8383, G8384, G8389, G839, I69031, I69032, I69033, I69034, I69039, I69041, I69042, I69043, I69044, I69049, I69051, I69052, I69053, I69054, I69059, I69061, I69062, I69063, I69064, I69065, I69069, I69131, I69132, I69133, I69134, I69139, I69141, I69142, I69143, I69144, I69149, I69151, I69152, I69153, I69154, I69159, I69161, I69162, I69163, I69164, I69165, I69169, I69231, I69232, I69233, I69234, I69239, I69241, I69242, I69243, I69244, I69249, I69251, I69252, I69253, I69254, I69259, I69261, I69262, I69263, I69264, I69265, I69269, I69331, I69332, I69333, I69334, I69339, I69341, I69342, I69343, I69344, I69349, I69351, I69352, I69353, I69354, I69359, I69361, I69362, I69363, I69364, I69365, I69369, I69831, I69832, I69833, I69834, I69839, I69841, I69842, I69843, I69844, I69849, I69851, I69852, I69853, I69854, I69859, I69861, I69862, I69863, I69864, I69865, I69869, I69931, I69932, I69933, I69934, I69939, I69941, I69942, I69943, I69944, I69949, I69951, I69952, I69953, I69954, I69959, I69961, I69962, I69963, I69964, I69965,<br>I69969<br>, |
| Multiple Sclerosis and Transverse Myelitis | 340, 341, 3410, 3412, 34120, 34121, 34122, 3418, 3419, G35, G360, G361, G368, G369, G371, G372, G373, G374, G378, G379 |
| Muscular Dystrophy | 359, 3590, 3591, G710, G7100, G7101, G7102, G7109, G7111, G712 |
| Obesity | 2780, 27800, 27801, 27803, E6601, E6609, E661, E662, E668, E669, V853, V8530, V8531, V8532, V8533, V8534, V8535, V8536, V8537, V8538, V8539, V854, V8541, V8542, V8543, V8544, V8545, Z6830, Z6831, Z6832, Z6833, Z6834, Z6835, Z6836, Z6837, Z6838, Z6839, Z6841, Z6842, Z6843, Z6844, Z6845 |

| | |
|---|---|
| Opioid Use Disorder | 11288, 3040, 30400, 30401, 30402, 3047, 30470, 30471, 30472, 3055, 30550, 30551, 30552, 76072, 9650, 96500, 96501, 96502, 96509, 9701, E8500, E8501, E8502, E9350, E9351, E9352, E9401, F1110, F11120, F11121, F11122, F11129, F1114, F11150, F11151, F11159, F11181, F11182, F11188, F1119, F1120, F11220, F11221, F11222, F11229, F1123, F1124, F11250, F11251, F11259, F11281, F11282, F11288, F1129, F1190, F11920, F11921, F11922, F11929, F1193, F1194, F11950, F11951, F11959, F11981, F11982, F11988, F1199, J0571, J0572, J0573, J0574, J0575, J1230, J2315, S0109, T400X1A, T400X1D, T400X1S, T400X2A, T400X2D, T400X2S, T400X3A, T400X3D, T400X3S, T400X4A, T400X4D, T400X4S, T400X5A, T400X5D, T400X5S, T401X1A, T401X1D, T401X1S, T401X2A, T401X2D, T401X2S, T401X3A, T401X3D, T401X3S, T401X4A, T401X4D, T401X4S, T402X1A, T402X1D, T402X1S, T402X2A, T402X2D, T402X2S, T402X3A, T402X3D, T402X3S, T402X4A, T402X4D, T402X4S, T402X5A, T402X5D, T402X5S, T403X1A, T403X1D, T403X1S, T403X2A, T403X2D, T403X2S, T403X3A, T403X3D, T403X3S, T403X4A, T403X4D, T403X4S, T403X5A, T403X5D, T403X5S, T404X1A, T404X1D, T404X1S, T404X2A, T404X2D, T404X2S, T404X3A, T404X3D, T404X3S, T404X4A, T404X4D, T404X4S, T404X5A, T404X5D, T404X5S, T40601A, T40601D, T40601S, T40602A, T40602D, T40602S, T40603A, T40603D, T40603S, T40604A, T40604D, T40604S, T40605A, T40605D, T40605S, T40691A, T40691D, T40691S, T40692A, T40692D, T40692S, T40693A, T40693D, T40693S, T40694A, T40694D, T40694S, T40695A, T40695D, T40695S |
| Osteoporosis | 73300, 73301, 73302, 73303, 73309, M810, M816, M818 |
| Other Developmental Delays | 3155, 3158, 3159, F819, F82, F88, F89 |
| Peripheral Vascular Disease (PVD) | 4400, 4401, 4402, 44020, 44021, 44022, 44023, 44029, 4404, 4438, 44381, 44382, 44389, 4439, E0851, E0852, E0951, E0952, E1051, E1052, E1151, E1152, E1351, E1352, I700, I701, I70201, I70202, I70203, I70208, I70209, I70211, I70212, I70213, I70218, I70219, I70221, I70222, I70223, I70228, I70229, I70231, I70232, I70233, I70234, I70235, I70238, I70239, I70241, I70242, I70243, I70244, I70245, I70248, I70249, I7025, I70291, I70292, I70293, I70298, I70299, I7092, I7381, I7389, I739, I791, I798 |
| Personality Disorders | 3010, 30110, 30111, 30112, 30113, 30120, 30121, 30122, 3013, 3014, 30150, 30151, 30159, 3016, 3017, 30181, 30182, 30183, 30184, 30189, 3019, F21, F340, F341, F600, F601, F602, F603, F604, F605, F606, F607, F6081, F6089, F609, F6810, F6811, F6812, F6813, F69 |
| Post-Traumatic Stress Disorder (PTSD) | 30981, F4310, F4311, F4312 |

| | |
|---|---|
| Pressure and Chronic Ulcers | 7070, 70700, 70701, 70702, 70703, 70704, 70705, 70706, 70707, 70709, 7071, 70710, 70711, 70712, 70713, 70714, 70715, 70719, 7072, 70722, 70723, 70724, 70725, 7078, 7079, I70231, I70232, I70233, I70234, I70235, I70238, I70239, I70241, I70242, I70243, I70244, I70245, I70248, I70249, I7025, I70331, I70332, I70333, I70334, I70335, I70338, I70339, I70341, I70342, I70343, I70344, I70345, I70348, I70349, I7035, I70431, I70432, I70433, I70434, I70435, I70438, I70439, I70441, I70442, I70443, I70444, I70445, I70448, I70449, I7045, I70531, I70532, I70533, I70534, I70535, I70538, I70539, I70541, I70542, I70543, I70544, I70545, I70548, I70549, I7055, I70631, I70632, I70633, I70634, I70635, I70638, I70639, I70641, I70642, I70643, I70644, I70645, I70648, I70649, I7065, I70731, I70732, I70733, I70734, I70735, I70738, I70739, I70741, I70742, I70743, I70744, I70745, I70748, I70749, I7075, L89000, L89001, L89002, L89003, L89004, L89009, L89010, L89011, L89012, L89013, L89014, L89019, L89020, L89021, L89022, L89023, L89024, L89029, L89100, L89101, L89102, L89103, L89104, L89109, L89110, L89111, L89112, L89113, L89114, L89119, L89120, L89121, L89122, L89123, L89124, L89129, L89130, L89131, L89132, L89133, L89134, L89139, L89140, L89141, L89142, L89143, L89144, L89149, L89150, L89151, L89152, L89153, L89154, L89159, L89200, L89201, L89202, L89203, L89204, L89209, L89210, L89211, L89212, L89213, L89214, L89219, L89220, L89221, L89222, L89223, L89224, L89229, L89300, L89301, L89302, L89303, L89304, L89309, L89310, L89311, L89312, L89313, L89314, L89319, L89320, L89321, L89322, L89323, L89324, L89329, L8940, L8941, L8942, L8943, L8944, L8945, L89500, L89501, L89502, L89503, L89504, L89509, L89510, L89511, L89512, L89513, L89514, L89519, L89520, L89521, L89522, L89523, L89524, L89529, L89600, L89601, L89602, L89603, L89604, L89609, L89610, L89611, L89612, L89613, L89614, L89619, L89620, L89621, L89622, L89623, L89624, L89629, L89810, L89811, L89812, L89813, L89814, L89819, L89890, L89891, L89892, L89893, L89894, L89899, L8990, L8991, L8992, L8993, L8994, L8995, L97101, L97102, L97103, L97104, L97105, L97106, L97108, L97109, L97111, L97112, L97113, L97114, L97115, L97116, L97118, L97119, L97121, L97122, L97123, L97124, L97125, L97126, L97128, L97129, L97201, L97202, L97203, L97204, L97205, L97206, L97208, L97209, L97211, L97212, L97213, L97214, L97215, L97216, L97218, L97219, L97221, L97222, L97223, L97224, L97225, L97226, L97228, L97229, L97301, L97302, L97303, L97304, L97305, L97306, L97308, L97309, L97311, L97312, L97313, L97314, L97315, L97316, L97318, L97319, L97321, L97322, L97323, L97324, L97325, L97326, L97328, L97329, L97401, L97402, L97403, L97404, L97405, L97406, L97408, L97409, L97411, L97412, L97413, L97414, L97415, L97416, L97418, L97419, L97421, L97422, L97423, L97424, L97425, L97426, L97428, L97429, L97501, L97502, L97503, L97504, L97505, L97506, L97508, L97509, L97511, L97512, L97513, L97514, L97515, L97516, L97518, L97519, L97521, L97522, L97523, L97524, L97525, L97526, L97528, L97529, L97801, L97802, L97803, L97804, L97805, L97806, L97808, L97809, L97811, L97812, L97813, L97814, L97815, L97816, L97818, L97819, L97821, L97822, L97823, L97824, L97825, L97826, L97828, L97829, L97901, L97902, L97903, L97904, L97905, L97906, L97908, L97909, L97911, L97912, L97913, L97914, L97915, L97916, L97918, L97919, L97921, L97922, L97923, L97924, L97925, L97926, L97928, L97929, L98411, L98412, L98413, L98414, L98415, L98416, L98418, L98419, L98421, L98422, L98423, L98424, L98425, L98426, L98428, L98429, L98491, L98492, L98493, L98494, L98495, L98496, L98498, L98499 , |
| Prostate Cancer | 185, 2334, C61, D075, V1046, Z8546 |
| RA/OA (Rheumatoid Arthritis/ Osteoarthritis) | 7140, 7141, 7142, 71430, 71431, 71432, 71433, 71500, 71504, 71509, 71510, 71511, 71512, 71513, 71514, 71515, 71516, 71517, 71518, 71520, 71521, 71522, 71523, 71524, 71525, 71526, 71527, 71528, 71530, 71531, 71532, 71533, 71534, 71535, 71536, 71537, 71538, 71580, 71589, 71590, 71591, 71592, 71593, 71594, 71595, 71596, 71597, 71598, 7200, 7210, 7211, 7212, 7213, 72190, 72191, M0500, M05011, M05012, M05019, M05021, M05022, M05029, M05031, M05032, M05039, M05041, M05042, M05049, M05051, M05052, M05059, M05061, M05062, M05069, M05071, M05072, M05079, M0509, M0520, M05211, M05212, M05219, M05221, M05222, M05229, M05231, M05232, M05239, M05241, M05242, M05249, M05251, M05252, M05259, M05261, M05262, M05269, M05271, M05272, M05279, M0529, M0530, M05311, M05312, M05319, M05321, M05322, M05329, M05331, M05332, M05339, |

| | |
|---|---|
| | M05341, M05342, M05349, M05351, M05352, M05359, M05361, M05362, M05369, M05371, M05372, M05379, M0539, M0540, M05411, M05412, M05419, M05421, M05422, M05429, M05431, M05432, M05439, M05441, M05442, M05449, M05451, M05452, M05459, M05461, M05462, M05469, M05471, M05472, M05479, M0549, M0550, M05511, M05512, M05519, M05521, M05522, M05529, M05531, M05532, M05539, M05541, M05542, M05549, M05551, M05552, M05559, M05561, M05562, M05569, M05571, M05572, M05579, M0559, M0560, M05611, M05612, M05619, M05621, M05622, M05629, M05631, M05632, M05639, M05641, M05642, M05649, M05651, M05652, M05659, M05661, M05662, M05669, M05671, M05672, M05679, M0569, M0570, M05711, M05712, M05719, M05721, M05722, M05729, M05731, M05732, M05739, M05741, M05742, M05749, M05751, M05752, M05759, M05761, M05762, M05769, M05771, M05772, M05779, M0579, M0580, M05811, M05812, M05819, M05821, M05822, M05829, M05831, M05832, M05839, M05841, M05842, M05849, M05851, M05852, M05859, M05861, M05862, M05869, M05871, M05872, M05879, M0589, M059, M0600, M06011, M06012, M06019, M06021, M06022, M06029, M06031, M06032, M06039, M06041, M06042, M06049, M06051, M06052, M06059, M06061, M06062, M06069, M06071, M06072, M06079, M0608, M0609, M061, M0620, M06211, M06212, M06219, M06221, M06222, M06229, M06231, M06232, M06239, M06241, M06242, M06249, M06251, M06252, M06259, M06261, M06262, M06269, M06271, M06272, M06279, M0628, M0629, M0630, M06311, M06312, M06319, M06321, M06322, M06329, M06331, M06332, M06339, M06341, M06342, M06349, M06351, M06352, M06359, M06361, M06362, M06369, M06371, M06372, M06379, M0638, M0639, M0680, M06811, M06812, M06819, M06821, M06822, M06829, M06831, M06832, M06839, M06841, M06842, M06849, M06851, M06852, M06859, M06861, M06862, M06869, M06871, M06872, M06879, M0688, M0689, M069, M0800, M08011, M08012, M08019, M08021, M08022, M08029, M08031, M08032, M08039, M08041, M08042, M08049, M08051, M08052, M08059, M08061, M08062, M08069, M08071, M08072, M08079, M0808, M0809, M081, M0820, M08211, M08212, M08219, M08221, M08222, M08229, M08231, M08232, M08239, M08241, M08242, M08249, M08251, M08252, M08259, M08261, M08262, M08269, M08271, M08272, M08279, M0828, M0829, M083, M0840, M08411, M08412, M08419, M08421, M08422, M08429, M08431, M08432, M08439, M08441, M08442, M08449, M08451, M08452, M08459, M08461, M08462, M08469, M08471, M08472, M08479, M0848, M0880, M08811, M08812, M08819, M08821, M08822, M08829, M08831, M08832, M08839, M08841, M08842, M08849, M08851, M08852, M08859, M08861, M08862, M08869, M08871, M08872, M08879, M0888, M0889, M0890, M08911, M08912, M08919, M08921, M08922, M08929, M08931, M08932, M08939, M08941, M08942, M08949, M08951, M08952, M08959, M08961, M08962, M08969, M08971, M08972, M08979, M0898, M0899, M150, M151, M152, M153, M154, M158, M159, M160, M1610, M1611, M1612, M162, M1630, M1631, M1632, M164, M1650, M1651, M1652, M166, M167, M169, M170, M1710, M1711, M1712, M172, M1730, M1731, M1732, M174, M175, M179, M180, M1810, M1811, M1812, M182, M1830, M1831, M1832, M184, M1850, M1851, M1852, M189, M19011, M19012, M19019, M19021, M19022, M19029, M19031, M19032, M19039, M19041, M19042, M19049, M19071, M19072, M19079, M19111, M19112, M19119, M19121, M19122, M19129, M19131, M19132, M19139, M19141, M19142, M19149, M19171, M19172, M19179, M19211, M19212, M19219, M19221, M19222, M19229, M19231, M19232, M19239, M19241, M19242, M19249, M19271, M19272, M19279, M1990, M1991, M1992, M1993, M450, M451, M452, M453, M454, M455, M456, M457, M458, M459, M47011, M47012, M47013, M47014, M47015, M47016, M47019, M47021, M47022, M47029, M4710, M4711, M4712, M4713, M4720, M4721, M4722, M4723, M4724, M4725, M4726, M4727, M4728, M47811, M47812, M47813, M47814, M47815, M47816, M47817, M47818, M47819, M47891, M47892, M47893, M47894, M47895, M47896, M47897, M47898, M47899, M479, M488X1, M488X2, M488X3, M488X4, M488X5, M488X6, M488X7, M488X8, M488X9 , |
| Schizophrenia | 29500, 29501, 29502, 29503, 29504, 29505, 29510, 29511, 29512, 29513, 29514, 29515, 29520, 29521, 29522, 29523, 29524, 29525, 29530, 29531, 29532, 29533, 29534, 29535, 29540, 29541, 29542, 29543, 29544, 29545, 29550, 29551, 29552, 29553, 29554, 29555, 29560, 29561, 29562, 29563, 29564, 29565, 29570, 29571, 29572, 29573, 29574, 29575, 29580, 29581, 29582, 29583, 29584, 29585, 29590, 29591, 29592, 29593, 29594, 29595, F200, F201, F202, F203, F205, F2081, F2089, F209, F250, F251, F258, F259 |

| | |
|---|---|
| Schizophrenia and Other Psychotic Disorders | 29381, 29382, 29500, 29501, 29502, 29503, 29504, 29505, 29510, 29511, 29512, 29513, 29514, 29515, 29520, 29521, 29522, 29523, 29524, 29525, 29530, 29531, 29532, 29533, 29534, 29535, 29540, 29541, 29542, 29543, 29544, 29545, 29550, 29551, 29552, 29553, 29554, 29555, 29560, 29561, 29562, 29563, 29564, 29565, 29570, 29571, 29572, 29573, 29574, 29575, 29580, 29581, 29582, 29583, 29584, 29585, 29590, 29591, 29592, 29593, 29594, 29595, 2970, 2971, 2972, 2973, 2978, 2979, 2980, 2981, 2982, 2983, 2984, 2988, 2989, F060, F062, F200, F201, F202, F203, F205, F2081, F2089, F209, F21, F22, F23, F24, F250, F251, F258, F259, F28, F29, F323, F333, F4489 |
| Sensory - Blindness and Visual Impairment | 369, 3690, 36900, 36901, 36902, 36903, 36904, 36905, 36906, 36907, 36908, 3691, 36910, 36911, 36912, 36913, 36914, 36915, 36916, 36917, 36918, 3692, 36920, 36921, 36922, 36923, 36924, 36925, 3693, 3694, H540, H540X33, H540X34, H540X35, H540X43, H540X44, H540X45, H540X53, H540X54, H540X55, H5410, H5411, H541131, H541132, H541141, H541142, H541151, H541152, H5412, H541213, H541214, H541215, H541223, H541224, H541225, H542, H542X11, H542X12, H542X21, H542X22, H543, H548 |
| Sensory – Deafness and Hearing Impairment | 389, 3891, 38910, 38911, 38912, 38913, 38914, 38915, 38916, 38917, 38918, 3892, 38920, 38921, 38922, 3897, 3898, 3899, H903, H9041, H9042, H905, H906, H9071, H9072, H908, H90A21, H90A22, H90A31, H90A32, H9101, H9102, H9103, H9109, H913, H918X1, H918X2, H918X3, H918X9, H9190, H9191, H9192, H9193 |
| Sickle Cell Disease | 28241, 28242, 28260, 28261, 28262, 28263, 28264, 28268, 28269, D5700, D5701, D5702, D571, D5720, D57211, D57212, D57219, D5740, D57411, D57412, D57419, D5780, D57811, D57812, D57819 |
| Spina Bifida and Other Congenital Anomalies of the Nervous System | 7400, 7401, 7402, 741, 7410, 74100 74101, 74102, 74103, 7419, 74190, 74191 74192, 74193, 7420, 7421, 7422, 7423, 7424, 7425, 74251, 74253, 74259, 7428, 7429, G901, Q000, Q001, Q002, Q010, Q011, Q012, Q018, Q019, Q02, Q030, Q031, Q038, Q039, Q040, Q041, Q042, Q043, Q044, Q045, Q046, Q048, Q049, Q050, Q051, Q052, Q053, Q054, Q055, Q056, Q057, Q058, Q059, Q060, Q061, Q062, Q063, Q064, Q068, Q069, Q0700, Q0701, Q0702, Q0703, Q078, Q079 |

| | |
|---|---|
| Spinal Cord Injury | 15, 34939, 80600 80601, 80602, 80603, 80604, 80605, 80606, 80607, 80608, 80609, 80610, 80611, 80612, 80613, 80614, 80615, 80616, 80617, 80618, 80619, 80620, 80621, 80622, 80623, 80624, 80625, 80626, 80627, 80628, 80629, 80630, 80631, 80632, 80633, 80634, 80635, 80636, 80637, 80638, 80639, 8064, 8065, 80660, 80661, 80662, 80669, 80670, 80671, 80672, 80679, 8068, 8069, 9072, 952, 95200, 95201, 95202, 95203, 95204, 95205, 95206, 95207, 95208, 95209, 95210, 95211, 95212, 95213, 95214, 95216, 95217, 95218, 95219, 9522, 9523, 9524, 9528, 9529, G9611, S12000A, S12000B, S12001A, S12001B, S12100A, S12100B, S12101A, S12101B, S12200A, S12200B, S12201A, S12201B, S12300A, S12300B, S12301A, S12301B, S12400A, S12400B, S12401A, S12401B, S12500A, S12500B, S12501A, S12501B, S12600A, S12600B, S12601A, S12601B, S129XXA, S13113A, S140XXA, S140XXS, S14101A, S14101S, S14102A, S14102S, S14103A, S14103S, S14104A, S14104S, S14105A, S14105S, S14106A, S14106S, S14107A, S14107S, S14108A, S14108S, S14109A, S14109S, S14111A, S14111S, S14112A, S14112S, S14113S, S14114A, S14114S, S14115A, S14115S, S14116A, S14116S, S14117A, S14117S, S14118A, S14118S, S14119A, S14119S, S14121A, S14121S, S14122A, S14122S, S14123A, S14123S, S14124A, S14124S, S14125A, S14125S, S14126A, S14126S, S14127A, S14127S, S14128A, S14128S, S14129A, S14129S, S14131A, S14131S, S14132A, S14132S, S14133A, S14133S, S14134A, S14134S, S14135A, S14135S, S14136A, S14136S, S14137A, S14137S, S14138A, S14138S, S14139A, S14139S, S14141A, S14141S, S14142A, S14142S, S14143A, S14143S, S14144A, S14144S, S14145A, S14145S, S14146A, S14146S, S14147A, S14147S, S14148A, S14148S, S14149A, S14149S, S14151A, S14151S, S14152A, S14152S, S14153A, S14153S, S14154A, S14154S, S14155A, S14155S, S14156A, S14156S, S14157A, S14157S, S14158A, S14158S, S14159A, S14159S, S22009A, S22009B, S22019A, S22019B, S22029A, S22029B, S22039A, S22039B, S22049A, S22049B, S22059A, S22059B, S22069A, S22069B, S22079A, S22079B, S22089A, S22089B, S240XXA, S240XXS, S24101A, S24101S, S24102A, S24102S, S24103A, S24103S, S24104A, S24104S, S24109A, S24109S, S24111A, S24111S, S24112A, S24112S, S24113A, S24113S, S24114A, S24114S, S24119A, S24119S, S24131A, S24131S, S24132A, S24132S, S24133A, S24133S, S24134A, S24134S, S24139A, S24139S, S24141A, S24141S, S24142A, S24142S, S24143A, S24143S, S24144A, S24144S, S24149A, S24149S, S24151A, S24151S, S24152A, S24152S, S24153A, S24153S, S24154A, S24154S, S24159A, S24159S, S32009A, S32009B, S32019A, S32019B, S32029A, S32029B, S32039A, S32039B, S32049A, S32049B, S32059A, S32059B, S3210XA, S3210XB, S322XXA, S322XXB, S3401XA, S3401XS, S3402XA, S3402XS, S34101A, S34101S, S34102A, S34102S, S34103A, S34103S, S34104A, S34104S, S34105A, S34105S, S34109A, S34109S, S34111A, S34111S, S34112A, S34112S, S34113A, S34113S, S34114A, S34114S, S34115A, S34115S, S34119A, S34119S, S34121A, S34121S, S34122A, S34122S, S34123A, S34123S, S34124A, S34124S, S34125A, S34125S, S34129A, S34129S, S34131A, S34131S, S34132A, S34132S, S34139A, S34139S, S343XXA |
| Stroke / Transient Ischemic Attack | 430, 431, 43301, 43311, 43321, 43331, 43381, 43391, 43400, 43401, 43410, 43411, 43490, 43491, 4350, 4351, 4353, 4358, 4359, 436, 99702, G450, G451, G452, G458, G459, G460, G461, G462, G463, G464, G465, G466, G467, G468, G9731, G9732, I6000, I6001, I6002, I6010, I6011, I6012, I6020, I6021, I6022, I6030, I6031, I6032, I604, I6050, I6051, I6052, I606, I607, I608, I609, I610, I611, I612, I613, I614, I615, I616, I618, I619, I6300, I63011, I63012, I63013, I63019, I6302, I63031, I63032, I63039, I6309, I6310, I63111, I63112, I63119, I6312, I63131, I63132, I63139, I6319, I6320, I63211, I63212, I63213, I63219, I6322, I63231, I63232, I63233, I63239, I6329, I6330, I63311, I63312, I63313, I63319, I63321, I63322, I63323, I63329, I63331, I63332, I63333, I63339, I63341, I63342, I63343, I63349, I6339, I6340, I63411, I63412, I63413, I63419, I63421, I63422, I63423, I63429, I63431, I63432, I63433, I63439, I63441, I63442, I63443, I63449, I6349, I6350, I63511, I63512, I63513, I63519, I63521, I63522, I63523, I63529, I63531, I63532, I63533, I63539, I63541, I63542, I63543, I63549, I6359, I636, I638, I639, I6601, I6602, I6603, I6609, I6611, I6612, I6613, I6619, I6621, I6622, I6623, I6629, I663, I668, I669, I67841, I67848, I6789, I97810, I97811, I97820, I97821 |

| | |
|---|---|
| Tobacco Use | 3051, 64900, 64901, 64902, 64903, 64904, 98984, F17200, F17201, F17203, F17208, F17209, F17210, F17211, F17213, F17218, F17219, F17220, F17221, F17223, F17228, F17229, F17290, F17291, F17293, F17298, F17299, O99330, O99331, O99332, O99333, O99334, O99335, T65211A, T65212A, T65213A, T65214A, T65221A, T65222A, T65223A, T65224A, T65291A, T65292A, T65293A, T65294A, Z720 |
| Traumatic Brain Injury and Nonpsychotic Mental Disorders due to Brain Damage | 310, 3100, 3101, 3102, 3108, 31081, 31089, 907, 9070, 9071, F070, F0781, F0789, F482, S04011S, S04012S, S04019S, S0402XS, S04031S, S04032S, S04039S, S04041S, S04042S, S04049S, S0410XS, S0411XS, S0412XS, S0420XS, S0421XS, S0422XS, S0430XS, S0431XS, S0432XS, S0440XS, S0441XS, S0442XS, S0450XS, S0451XS, S0452XS, S0460XS, S0461XS, S0462XS, S0470XS, S0471XS, S0472XS, S04811S, S04812S, S04819S, S04891S, S04892S, S04899S, S049XXS, S060X0S, S060X1S, S060X2S, S060X3S, S060X4S, S060X5S, S060X6S, S060X7S, S060X8S, S060X9S, S061X0S, S061X1S, S061X2S, S061X3S, S061X4S, S061X5S, S061X6S, S061X7S, S061X8S, S061X9S, S062X0S, S062X1S, S062X2S, S062X3S, S062X4S, S062X5S, S062X6S, S062X7S, S062X8S, S062X9S, S06300S, S06301S, S06302S, S06303S, S06304S, S06305S, S06306S, S06307S, S06308S, S06309S, S06310S, S06311S, S06312S, S06313S, S06314S, S06315S, S06316S, S06317S, S06318S, S06319S, S06320S, S06321S, S06322S, S06323S, S06324S, S06325S, S06326S, S06327S, S06328S, S06329S, S06330S, S06331S, S06332S, S06333S, S06334S, S06335S, S06336S, S06337S, S06338S, S06339S, S06340S, S06341S, S06342S, S06343S, S06344S, S06345S, S06346S, S06347S, S06348S, S06349S, S06350S, S06351S, S06352S, S06353S, S06354S, S06355S, S06356S, S06357S, S06358S, S06359S, S06360S, S06361S, S06362S, S06363S, S06364S, S06365S, S06366S, S06367S, S06368S, S06369S, S06370S, S06371S, S06372S, S06373S, S06374S, S06375S, S06376S, S06377S, S06378S, S06379S, S06380S, S06381S, S06382S, S06383S, S06384S, S06385S, S06386S, S06387S, S06388S, S06389S, S064X0S, S064X1S, S064X2S, S064X3S, S064X4S, S064X5S, S064X6S, S064X7S, S064X8S, S064X9S, S065X0S, S065X1S, S065X2S, S065X3S, S065X4S, S065X5S, S065X6S, S065X7S, S065X8S, S065X9S, S066X0S, S066X1S, S066X2S, S066X3S, S066X4S, S066X5S, S066X6S, S066X7S, S066X8S, S066X9S, S06810S, S06811S, S06812S, S06813S, S06814S, S06815S, S06816S, S06817S, S06818S, S06819S, S06820S, S06821S, S06822S, S06823S, S06824S, S06825S, S06826S, S06827S, S06828S, S06829S, S06890S, S06891S, S06892S, S06893S, S06894S, S06895S, S06896S, S06897S, S06898S, S06899S, S069X0S, S069X1S, S069X2S, S069X3S, S069X4S, S069X5S, S069X6S, S069X7S, S069X8S, S069X9S |
| Viral Hepatitis (General) | 0700, 0701, 0702, 07020, 07021, 07022, 07023, 0703, 07030, 07031, 07032, 07033, 0704, 07041, 07042, 07043, 07049, 0705, 07051, 07052, 07053, 07054, 07059, 0706, 0707, 07070, 07071, 0709, B150, B159, B160, B161, B162, B169, B170, B1710, B1711, B172, B178, B179, B180, B181, B182, B188, B189, B190, B1910, B1911, B1920, B1921, B199, V026, V0260, V0261, V0262, V0269, Z2250, Z2251, Z2252, Z2259 |
| Sleep disorders | F13282, 78058, Z73811, F13288, G4722, 78051, F13982, 32726, P283, G4733, G47411, 30744, 34701, 34710, 32737, G47429, F515, G4711, 32720, 30746, 4672, 32734, 32743, 32730, 34711, F514, 32712, F19282, F14182, G4752, 78055, 29285, G4709, G4734, 30749, G4720, G4737, 32727, G47421, F11182, 32729, F14282, F5103, 32736, G4723, G4753, F13182, 30745, 30741, G4712, Z73819, G4730, G4701, F5112, F5105, G4726, 29182, G4739, Z73812, 32724, 30743, 78054, G4762, G478, 30742, F10282, F15182, F19982, V694, 32701, G4763, 32721, F513, 78056, 78057, 32700, F5104, 32725, G4724, 32722, 32732, 32739, G4769, 32740, 32731, 32711, 32709, 32723, Z72820, 78052, A8183, 32702, 32735, 32742, F10982, F5101, 30740, F11282, F519, Z73810, G4725, F14982, G4727, F11982, 30747, G4729, 78053, F5109, F518, 32733, G4721, G4700, F15982, 32759, G47419, G4731, F10182, 78059, G4736, F15282, 3278, F5102, 30748, 34700, G479, Z72821 |

| | |
|---|---|
| Periodontitis | K05319, 52342, K056, 52341, K08121, K08124, K05212, K05223, K08122, K0530, K054, 5239, 52331, K044, K08421, K05222, K05313, K05312, K05323, K08429, K05321, K05329, 52330, K05211, K05221, 5238, K08424, K05311, K08129, 5226, K055, K05229, K08422, K05213, 5235, K05219, K0520, 52340, 52333, K045, K05322, 5224, 52332, K08123, K08423 |
| Menopause | E28319, 6274, 6270, E28310, N924, 25631 |

### Supplementary Figures

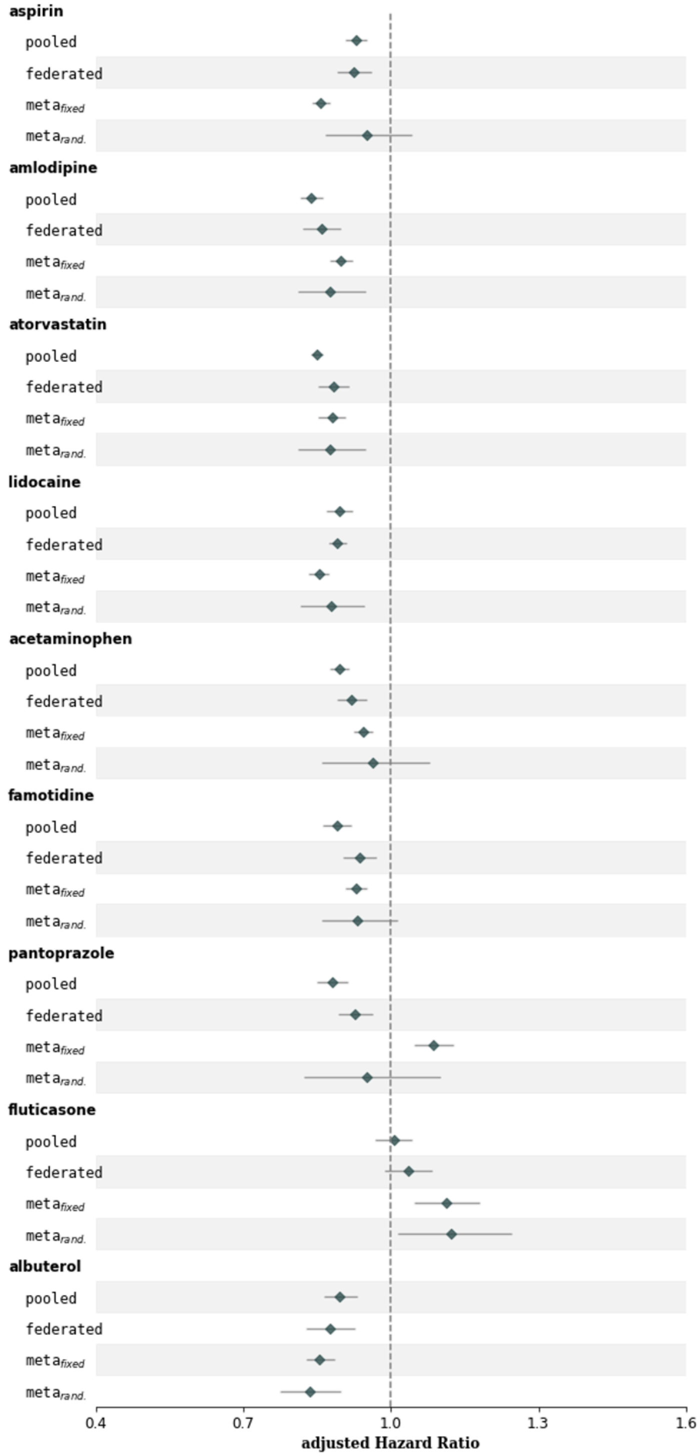


| Drug / Method | aHR | 95% CI | P-value | Z-Test statistic (compared with pooled-analysis) | P-value (vs. pooled) |
|---|---|---|---|---|---|
| **aspirin** | | | | | |
| pooled | 0.93 | (0.91, 0.95) | 9.94e-10$^{***}$ | 0.00 | 1.000$^{ns}$ |
| federated | 0.93 | (0.89, 0.96) | 5.04e-05$^{***}$ | 0.18 | 0.860$^{ns}$ |
| $meta_{fixed}$ | 0.86 | (0.84, 0.88) | 1.94e-45$^{***}$ | 4.97 | 6.73e-07$^{***}$ |
| $meta_{rand.}$ | 0.95 | (0.87, 1.04) | 0.290$^{ns}$ | 0.46 | 0.643$^{ns}$ |
| **amlodipine** | | | | | |
| pooled | 0.84 | (0.81, 0.86) | 1.19e-35$^{***}$ | 0.00 | 1.000$^{ns}$ |
| federated | 0.86 | (0.82, 0.90) | 1.25e-11$^{***}$ | 0.91 | 0.362$^{ns}$ |
| $meta_{fixed}$ | 0.90 | (0.88, 0.92) | 2.15e-15$^{***}$ | 3.62 | 2.96e-04$^{***}$ |
| $meta_{rand.}$ | 0.88 | (0.81, 0.95) | 9.15e-04$^{***}$ | 1.08 | 0.282$^{ns}$ |
| **atorvastatin** | | | | | |
| pooled | 0.85 | (0.84, 0.86) | 7.77e-123$^{***}$ | 0.00 | 1.000$^{ns}$ |
| federated | 0.88 | (0.85, 0.92) | 2.15e-11$^{***}$ | 1.97 | 4.94e-02$^{*}$ |
| $meta_{fixed}$ | 0.88 | (0.85, 0.91) | 3.63e-15$^{***}$ | 2.06 | 3.90e-02$^{*}$ |
| $meta_{rand.}$ | 0.88 | (0.81, 0.95) | 1.00e-03$^{**}$ | 0.81 | 0.419$^{ns}$ |
| **lidocaine** | | | | | |
| pooled | 0.90 | (0.87, 0.92) | 8.70e-13$^{***}$ | 0.00 | 1.000$^{ns}$ |
| federated | 0.89 | (0.87, 0.91) | 5.04e-30$^{***}$ | 0.25 | 0.805$^{ns}$ |
| $meta_{fixed}$ | 0.85 | (0.83, 0.87) | 1.14e-39$^{***}$ | 2.41 | 1.60e-02$^{*}$ |
| $meta_{rand.}$ | 0.88 | (0.82, 0.95) | 6.89e-04$^{***}$ | 0.45 | 0.652$^{ns}$ |
| **acetaminophen** | | | | | |
| pooled | 0.89 | (0.88, 0.91) | 1.83e-24$^{***}$ | 0.00 | 1.000$^{ns}$ |
| federated | 0.92 | (0.89, 0.95) | 3.75e-07$^{***}$ | 1.43 | 0.154$^{ns}$ |
| $meta_{fixed}$ | 0.94 | (0.93, 0.96) | 1.71e-08$^{***}$ | 3.57 | 3.53e-04$^{***}$ |
| $meta_{rand.}$ | 0.96 | (0.86, 1.08) | 0.519$^{ns}$ | 1.23 | 0.217$^{ns}$ |
| **famotidine** | | | | | |
| pooled | 0.89 | (0.86, 0.92) | 1.74e-12$^{***}$ | 0.00 | 1.000$^{ns}$ |
| federated | 0.94 | (0.90, 0.97) | 3.97e-04$^{***}$ | 1.98 | 4.78e-02$^{*}$ |
| $meta_{fixed}$ | 0.93 | (0.91, 0.95) | 1.16e-09$^{***}$ | 2.03 | 4.19e-02$^{*}$ |
| $meta_{rand.}$ | 0.93 | (0.86, 1.01) | 0.098$^{ns}$ | 1.02 | 0.306$^{ns}$ |
| **pantoprazole** | | | | | |
| pooled | 0.88 | (0.85, 0.91) | 2.95e-12$^{***}$ | 0.00 | 1.000$^{ns}$ |
| federated | 0.93 | (0.89, 0.96) | 8.15e-05$^{***}$ | 1.98 | 4.72e-02$^{*}$ |
| $meta_{fixed}$ | 1.09 | (1.05, 1.13) | 6.63e-06$^{***}$ | 8.11 | 5.14e-16$^{***}$ |
| $meta_{rand.}$ | 0.95 | (0.82, 1.10) | 0.505$^{ns}$ | 1.03 | 0.304$^{ns}$ |
| **fluticasone** | | | | | |
| pooled | 1.01 | (0.97, 1.04) | 0.779$^{ns}$ | 0.00 | 1.000$^{ns}$ |
| federated | 1.03 | (0.99, 1.08) | 0.150$^{ns}$ | 0.94 | 0.348$^{ns}$ |
| $meta_{fixed}$ | 1.11 | (1.05, 1.18) | 4.10e-04$^{***}$ | 2.83 | 4.66e-03$^{**}$ |
| $meta_{rand.}$ | 1.12 | (1.01, 1.25) | 2.73e-02$^{*}$ | 1.98 | 4.79e-02$^{*}$ |
| **albuterol** | | | | | |
| pooled | 0.90 | (0.86, 0.93) | 1.16e-08$^{***}$ | 0.00 | 1.000$^{ns}$ |
| federated | 0.88 | (0.83, 0.93) | 5.38e-06$^{***}$ | 0.65 | 0.513$^{ns}$ |
| $meta_{fixed}$ | 0.86 | (0.83, 0.89) | 2.83e-19$^{***}$ | 1.81 | 0.070$^{ns}$ |
| $meta_{rand.}$ | 0.83 | (0.78, 0.90) | 9.01e-07$^{***}$ | 1.74 | 0.082$^{ns}$ |

**Supplementary Figure 1: The estimated aHR and 95% CI on INSIGHT under differential privacy**. The compared methods consist of pooled-analysis (labeled as "pooled" in the figure), our FL-TTE (labeled as "federated"), and meta-analysis with fixed effects and random effects (labeled as "$meta_{fixed}$" and "$meta_{rand.}$"). The third column in the right side is p-value and significance level of the Z-test on whether the estimated aHR is significantly different with 1.0 (reference value indicating the treatment does not alter the risk compared to no treatment). The fourth and fifth columns denote the test statistic and p-value of the Z-test on whether the estimated aHR is significantly different with the results of pooled analysis. Our FL-TTE still achieved less-biased treatment effect estimates than two typical meta-analysis methods when compared to the estimates from the pooled data even using differential privacy techniques. *p < 0.05; **p < 0.01; ***p <

0.001; not significant (“ns”) with p ≥ 0.05.

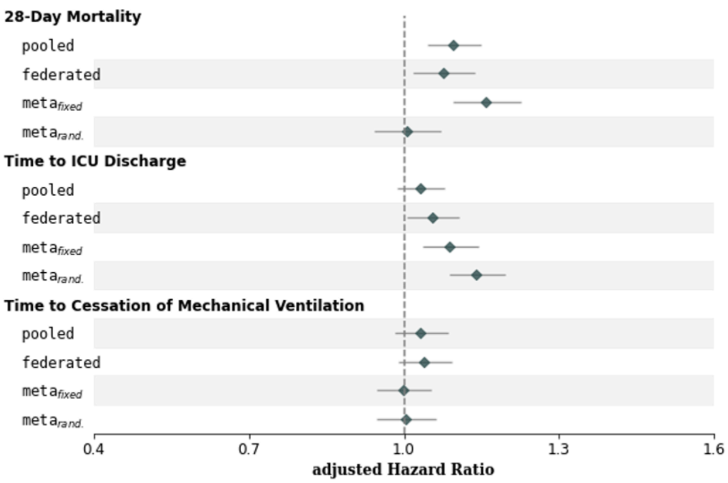


| | Estimates | | | Compared with pooled-analysis | |
|---|---|---|---|---|---|
| | aHR | 95% CI | P-value | Z-Test statistic | P-value (vs. pooled) |
| **28-Day Mortality** | | | | | |
| pooled | 1.10 | (1.05, 1.15) | 1.17e-04$^{***}$ | 0.00 | 1.000$^{ns}$ |
| federated | 1.08 | (1.02, 1.14) | 9.02e-03$^{**}$ | 0.50 | 0.617$^{ns}$ |
| $meta_{fixed}$ | 1.16 | (1.09, 1.23) | 4.33e-07$^{***}$ | 1.47 | 0.141$^{ns}$ |
| $meta_{rand.}$ | 1.01 | (0.94, 1.07) | 0.877$^{ns}$ | 2.14 | 3.27e-02$^{*}$ |
| **Time to ICU Discharge** | | | | | |
| pooled | 1.03 | (0.99, 1.08) | 0.176$^{ns}$ | 0.00 | 1.000$^{ns}$ |
| federated | 1.05 | (1.01, 1.11) | 3.01e-02$^{*}$ | 0.67 | 0.501$^{ns}$ |
| $meta_{fixed}$ | 1.09 | (1.04, 1.14) | 6.88e-04$^{***}$ | 1.61 | 0.107$^{ns}$ |
| $meta_{rand.}$ | 1.14 | (1.09, 1.20) | 6.32e-08$^{***}$ | 3.02 | 2.53e-03$^{**}$ |
| **Time to Cessation of Mechanical Ventilation** | | | | | |
| pooled | 1.03 | (0.98, 1.08) | 0.214$^{ns}$ | 0.00 | 1.000$^{ns}$ |
| federated | 1.04 | (0.99, 1.09) | 0.133$^{ns}$ | 0.20 | 0.840$^{ns}$ |
| $meta_{fixed}$ | 1.00 | (0.95, 1.05) | 0.958$^{ns}$ | 0.89 | 0.374$^{ns}$ |
| $meta_{rand.}$ | 1.00 | (0.95, 1.06) | 0.913$^{ns}$ | 0.74 | 0.459$^{ns}$ |

**Supplementary Figure 2: The estimated aHR and 95% CI on eICU-MIMIC under differential privacy**. The compared methods consist of pooled-analysis (labeled as “pooled” in the figure), our FL-TTE (labeled as “federated”), and meta-analysis with fixed effects and random effects (labeled as “$meta_{fixed}$” and “$meta_{rand.}$”). The third column in the right side is p-value and significance level of the Z-test on whether the estimated aHR is significantly different with 1.0 (reference value indicating the treatment does not alter the risk compared to no treatment). The fourth and fifth columns denote the test statistic and p-value of the Z-test on whether the estimated aHR is significantly different with the results of pooled analysis. Our FL-TTE still achieved less-biased treatment effect estimates than two typical meta-analysis methods when compared to the estimates from the pooled data even using differential privacy techniques. *p < 0.05; **p < 0.01; ***p < 0.001; not significant (“ns”) with p ≥ 0.05.

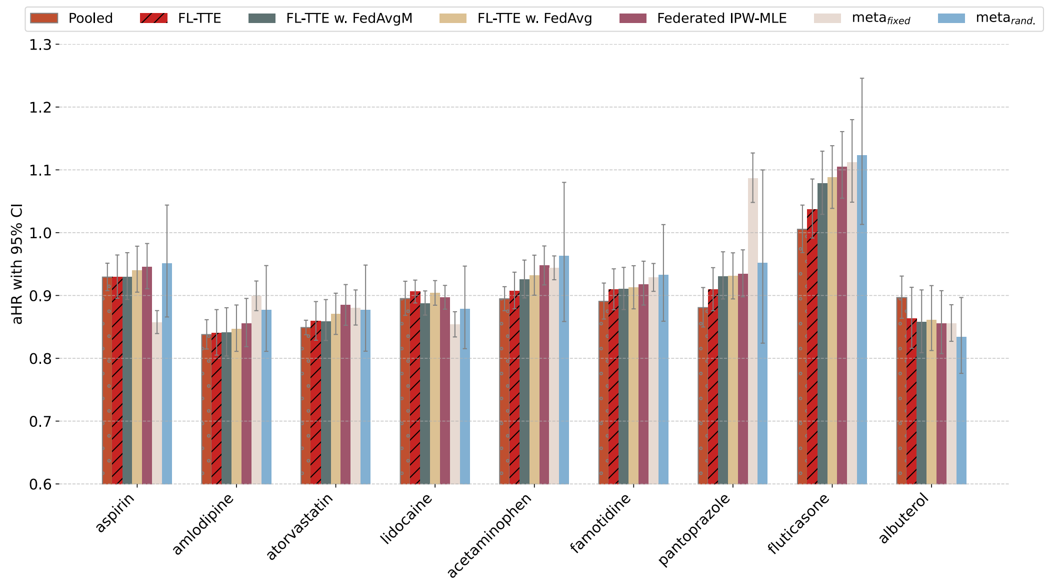


**Supplementary Figure 3: Sensitivity analysis on different federated learning algorithms on INSIGHT dataset.** The FL-TTE with the proximal term achieved less-biased estimates, slightly better than using the other FL algorithms, including FedAvgM that introduces the momentum to address heterogeneity, FedAvg that there are no explicit designs for handling heterogeneity and Federated IPW-MLE.

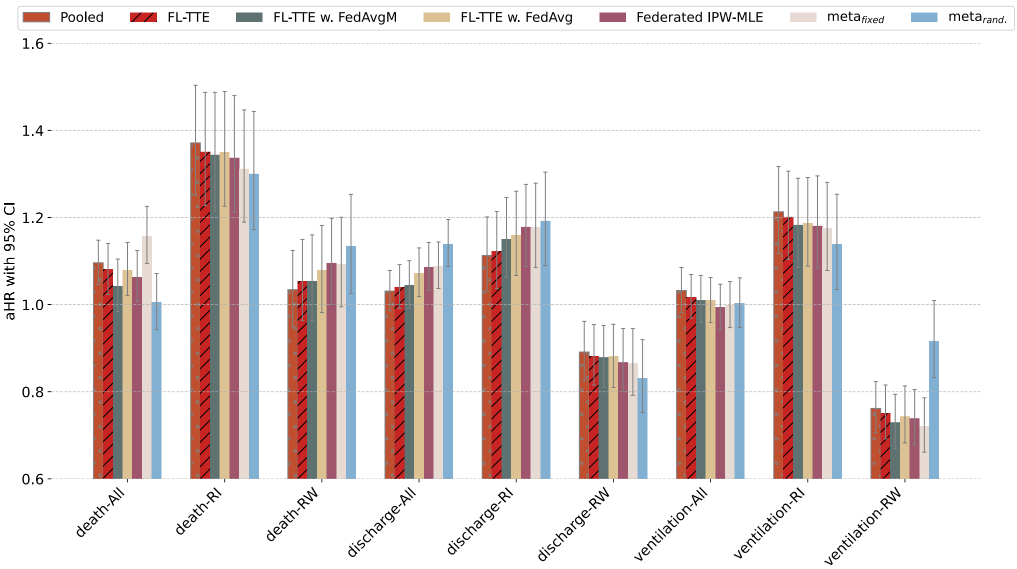


**Supplementary Figure 4: Sensitivity analysis on different federated learning algorithms on eICU-MIMIC dataset.** The FL-TTE with the proximal term achieved less-biased estimates, slightly better than using the other FL algorithms, including FedAvgM that introduces the momentum to address heterogeneity, FedAvg that there are no explicit designs for handling heterogeneity and Federated IPW-MLE.

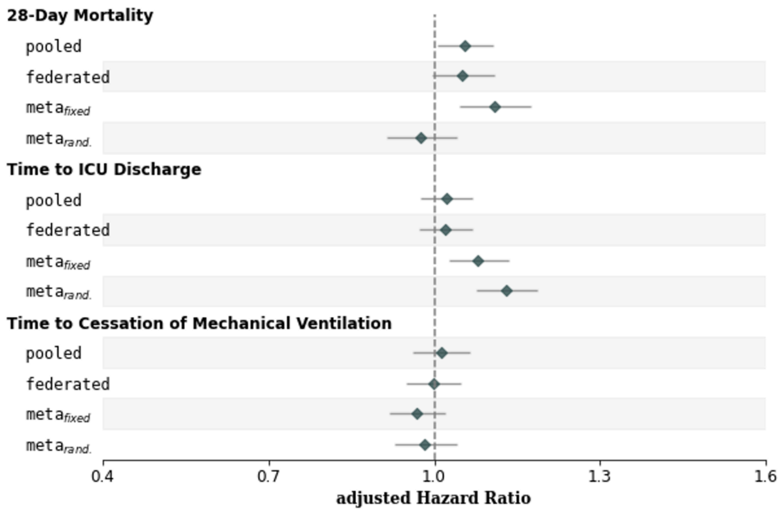


| | Estimates | | | Compared with pooled-analysis | |
|---|---|---|---|---|---|
| | aHR | 95% CI | P-value | Z-Test statistic | P-value (vs. pooled) |
| **28-Day Mortality** | | | | | |
| pooled | 1.06 | (1.01, 1.11) | 2.50e-02$^{*}$ | 0.00 | 1.000$^{ns}$ |
| federated | 1.05 | (1.00, 1.11) | 0.073$^{ns}$ | 0.15 | 0.885$^{ns}$ |
| $meta_{fixed}$ | 1.11 | (1.05, 1.17) | 4.91e-04$^{***}$ | 1.27 | 0.206$^{ns}$ |
| $meta_{rand.}$ | 0.98 | (0.91, 1.04) | 0.451$^{ns}$ | 1.93 | 0.054$^{ns}$ |
| **Time to ICU Discharge** | | | | | |
| pooled | 1.02 | (0.98, 1.07) | 0.360$^{ns}$ | 0.00 | 1.000$^{ns}$ |
| federated | 1.02 | (0.97, 1.07) | 0.412$^{ns}$ | 0.03 | 0.974$^{ns}$ |
| $meta_{fixed}$ | 1.08 | (1.03, 1.13) | 3.01e-03$^{**}$ | 1.60 | 0.110$^{ns}$ |
| $meta_{rand.}$ | 1.13 | (1.08, 1.19) | 8.81e-07$^{***}$ | 2.99 | 2.77e-03$^{**}$ |
| **Time to Cessation of Mechanical Ventilation** | | | | | |
| pooled | 1.01 | (0.96, 1.07) | 0.646$^{ns}$ | 0.00 | 1.000$^{ns}$ |
| federated | 1.00 | (0.95, 1.05) | 0.937$^{ns}$ | 0.39 | 0.700$^{ns}$ |
| $meta_{fixed}$ | 0.97 | (0.92, 1.02) | 0.227$^{ns}$ | 1.18 | 0.237$^{ns}$ |
| $meta_{rand.}$ | 0.98 | (0.93, 1.04) | 0.564$^{ns}$ | 0.74 | 0.462$^{ns}$ |

**Supplementary Figure 5: The estimated aHR and 95% CI on eICU-MIMIC using clone-censor-weight approach to limit immortal time bias.** The compared methods consist of pooled-analysis (labeled as "pooled" in the figure), our FL-TTE (labeled as "federated"), and meta-analysis with fixed effects and random effects (labeled as "$meta_{fixed}$" and "$meta_{rand.}$"). The third column in the right side is p-value and significance level of the Z-test on whether the estimated aHR is significantly different with 1.0 (reference value indicating the treatment does not alter the risk compared to no treatment). The fourth and fifth columns denote the test statistic and p-value of the Z-test on whether the estimated aHR is significantly different with the results of pooled analysis. Our FL-TTE still achieved less-biased treatment effect estimates than two typical meta-analysis methods when compared to the estimates from the pooled data. *$p < 0.05$; **$p < 0.01$; ***$p < 0.001$; not significant ("ns") with $p \geq 0.05$.
